# Phenotype-Specific Recalibration of MAVE Data Enables Repurposing of *BAP1* Functional Assays for Küry-Isidor Syndrome

**DOI:** 10.64898/2026.05.15.26352805

**Authors:** Pankhuri Gupta, Elsa V. Balton, Mavika Tejura, Runjun D. Kumar, Matthew W. Snyder, Jeremy Stone, Rehan M. Villani, Peter H. Byers, Sirisak Chanprasert, Ian A. Glass, Martha Horike-Pyne, Danny E. Miller, Jane Ranchalis, Elisabeth A. Rosenthal, Andrew K. Solomon, Mark Wener, Undiagnosed Diseases Network (UDN), Kathleen A. Leppig, Matthew D. Benson, Melissa J. MacPherson, Gail P. Jarvik, Katrina M. Dipple, Elizabeth E. Blue, Douglas M. Fowler, Lea M. Starita, Abbye E. McEwen, Andrew B. Stergachis

## Abstract

**Purpose:** Multiplexed assays of variant effect (MAVEs) are transforming clinical variant interpretation. However, many genes are associated with more than one disease, making it unclear whether functional data generated in one disease context may be directly applicable to another. For example, germline *BAP1* missense variants are associated with both *BAP1* tumor predisposition syndrome (*BAP1*-TPDS) and Küry-Isidor syndrome (KURIS), a rare neurodevelopmental disorder. Here, we demonstrate how phenotype-specific calibration of *BAP1* MAVE data enables disease-specific variant classification.

**Methods:** Saturation genome editing (SGE) data for *BAP1* were recalibrated using either *BAP1*-TPDS- or KURIS-associated missense variants as pathogenic controls. Functional evidence strength was quantified using the Odds of Pathogenicity (OddsPath) framework and mapped to ACMG/AMP PS3/BS3 criteria. Recalibrated functional evidence was integrated with standard clinical criteria for variant classification. A workshop was developed to teach phenotype-specific MAVE recalibration to clinicians and variant curators and evaluated for educational impact.

**Results:** Phenotype-specific recalibration using *BAP1*-TPDS and KURIS controls yielded OddsPath values consistent with PS3_Strong evidence in both contexts. Application of KURIS-specific recalibration enabled the diagnosis of KURIS in an individual with a previously uncertain *BAP1* missense variant. The educational workshop enabled quantitatively improved understanding in applying functional evidence.

**Conclusion:** Phenotype-specific recalibration enables appropriately calibrated reuse of MAVE datasets across distinct disease contexts, increasing the clinical utility of MAVE datasets and the interpretability of variants in pleiotropic genes. This framework expands the diagnostic utility of existing functional datasets without requiring new experimental assays.

## Introduction

Multiplexed functional approaches such as variant effect maps have the potential to transform clinical variant interpretation by enabling systematic functional assessment of large numbers of genetic variants in parallel^1^. To date, however, most variant effect maps have been generated and applied in the context of relatively common Mendelian conditions, and are typically interpreted within a single gene-disease framework. However, many disease-associated genes are pleiotropic, giving rise to multiple, phenotypically distinct disorders through variants that might affect protein function in different ways^2^. MAVEs often measure downstream functional impacts, which may capture multiple molecular mechanisms of disruption.

This motivates a phenotype-specific calibration-based approach to MAVE data, as existing variant effect maps may contain underutilized information that could be repurposed to inform interpretation in rarer disease contexts, once appropriately recalibrated.

Germline pathogenic variants in *BAP1* (*BRCA1*-associated deubiquitinase 1, HGNC:950) are classically associated with *BAP1* tumor predisposition syndrome (*BAP1*-TPDS, MIM: 614327), an autosomal dominant condition characterized by increased risk for malignancies including uveal and cutaneous melanoma, mesothelioma, and renal cell carcinoma^3,4^. However, recent work has established a distinct *BAP1*-associated syndromic neurodevelopmental disorder, Küry-Isidor syndrome (KURIS, MIM: 619762)^5^. KURIS presents with developmental delay, hypotonia, autism, seizures, and non-specific dysmorphic features^6^. In contrast to *BAP1*-TPDS, which is usually caused by heterozygous null variants, KURIS has only been reported in the context of heterozygous missense variants^7^.

Here, we present how phenotype-specific recalibration can be used to enable a MAVE for *BAP1* to be used to separately provide evidence for the pathogenicity of *BAP1*-TPDS or KURIS-associated *BAP1* variants. Furthermore, we demonstrate the application of this recalibrated functional evidence towards solving the genetic basis of a participant within the Undiagnosed Diseases Network (UDN) with a *BAP1* missense variant and present a tested educational workshop for teaching clinicians on phenotype-specific recalibration of MAVE data. Our findings provide a practical framework for appropriately adapting functional data across disease contexts and illustrate how phenotype-specific calibration can strengthen clinical interpretation of variants for rare diseases.

## Materials and Methods

### Subject evaluation

This study was approved by the National Institutes of Health Institutional Review Board (IRB) (IRB protocol #15HG0130), and written informed consent was obtained from all participants in the study. Detailed medical records were reviewed by Undiagnosed Diseases Network (UDN) investigators, and the participant was evaluated by UDN investigators. Clinical trio genome sequencing was performed on buccal cells at Baylor Genetics.

### Functional assay data source

Functional scores for *BAP1* variants were obtained from a published saturation genome editing (SGE) assay by Waters et al^7^. In this assay, all possible single nucleotide variants were introduced into a HAP1 LIG4-knockout cell line dependent on *BAP1* for survival. Each variant was assigned a functional score representing its change in frequency over time relative to wild-type (centered at zero).

### Control variant ascertainment

P/LP and B/LB *BAP1* variants were obtained from ClinVar (accessed February 2026). For the initial evidence strength calculation, all variants with review status of at least one star and a classification of pathogenic, likely pathogenic, benign, likely benign, or benign/likely benign were included, yielding 186 P/LP and 1,171 B/LB variants.

The 17 KURIS-associated *de novo* missense variants used as phenotype-specific pathogenic controls were drawn from the *BAP1*-associated neurodevelopmental disorder literature and public databases. Sources included all 11 families reported in Küry et al. 2022^5^ (comprising 9 unique variants), additional variants from DECIPHER^8^ with *de novo* inheritance and KURIS-consistent neurodevelopmental phenotypes (n=3), and variants classified as Pathogenic or Likely Pathogenic in ClinVar for a *BAP1*-related neurodevelopmental condition (n=8). Variants that occurred across multiple sources were only counted once. Benign controls were restricted to the 28 B/LB missense variants in ClinVar to match variant type with the pathogenic KURIS controls. Supplemental Table 1 contains all variants used as calibration controls.

### Evidence strength assignment

Translating MAVE data results into clinical evidence requires systematic calibration against variants of known pathogenicity. Here we used the established Odds of Pathogenicity (OddsPath) framework^9^. This method uses variants with established clinical classifications (pathogenic/likely pathogenic [P/LP] and benign/likely benign [B/LB]) as presumed controls to calculate likelihood ratios reflecting how strongly a given functional result predicts pathogenicity or benignity. These likelihood ratios are then mapped to ACMG/AMP evidence strength levels.

LR+ = OddsPath_Abnormal = [P(abnormal | P/LP)] / [P(abnormal | B/LB)]

LR- = OddsPath_Normal = [P(normal | P/LP)] / [P(normal | B/LB)]

Zero counts were handled by a Haldane-Anscombe correction, which added 0.5 to all counts^10,11^. OddsPath values were mapped to ACMG/AMP evidence strength levels using established thresholds^9^.

### Threshold determination

A two-component Gaussian mixture model was fitted to the *BAP1* functional scores using scikit-learn (version 1.7.1). Initial means for the two components were set using the mean of the middle 95% of scores from synonymous variants expected to be functionally neutral and the mean score of nonsense variants expected to be functionally deleterious. Each variant was then assigned a probability of belonging to each fitted component. Variants with greater than 0.95 probability of membership in a given component were classified as “functionally abnormal” or “functionally normal,” corresponding to the components with the lesser and greater means, respectively. Single nucleotide variants (SNVs) not meeting this threshold were classified as “indeterminate”. Approximate score boundaries for functional class assignment were determined by identifying the variant scores closest to each 0.95 probability threshold.

### Variant Classification

gnomAD v4.0 (gnomad.broadinstitute.org) was manually queried for population frequency. *De novo* occurrence in the proband was confirmed through trio-genome sequencing at Baylor Genetics. Phenotypic information for the proband was obtained through consultation reports by the proband’s clinicians at the Undiagnosed Diseases Network (UDN) and external institutions. For variant classification, functional data was applied in conjunction with existing population frequency, phenotypic and additional published *de novo* occurrences. ACMG/AMP points based thresholds were used for the classification^12^.

## Results

### *BAP1* Genetic Variants causing *BAP1*-TPDS versus KURIS

The molecular landscape of *BAP1* variants underlying *BAP1* tumor predisposition syndrome (*BAP1*-TPDS) is dominated by loss-of-function alleles, including nonsense, frameshift, and canonical splice site variants, with only a minority of definitively classified *BAP1*-TPDS variants being missense changes in *BAP1* (**Fig. 1**). *BAP1*-TPDS missense variants are primarily located in the ubiquitin C-terminal hydrolase (UCH) catalytic domain (amino acids 4-235) and have been shown to disrupt *BAP1* function through multiple molecular mechanisms including protein aggregation, impaired nuclear localization, disrupted ubiquitin binding, reduced deubiquitinase activity, and defective protein-protein interactions^13,14,15,16^. In contrast, KURIS is a phenotypically distinct *BAP1*-associated neurodevelopmental disorder that has been exclusively associated with heterozygous *de novo* missense variants^5^. These variants are also primarily located in the UCH catalytic domain (**Fig. 1a**) but have been shown to impair H2A deubiquitination and induce genome-wide chromatin state alterations affecting developmental genes^5^.

**Figure 1:**
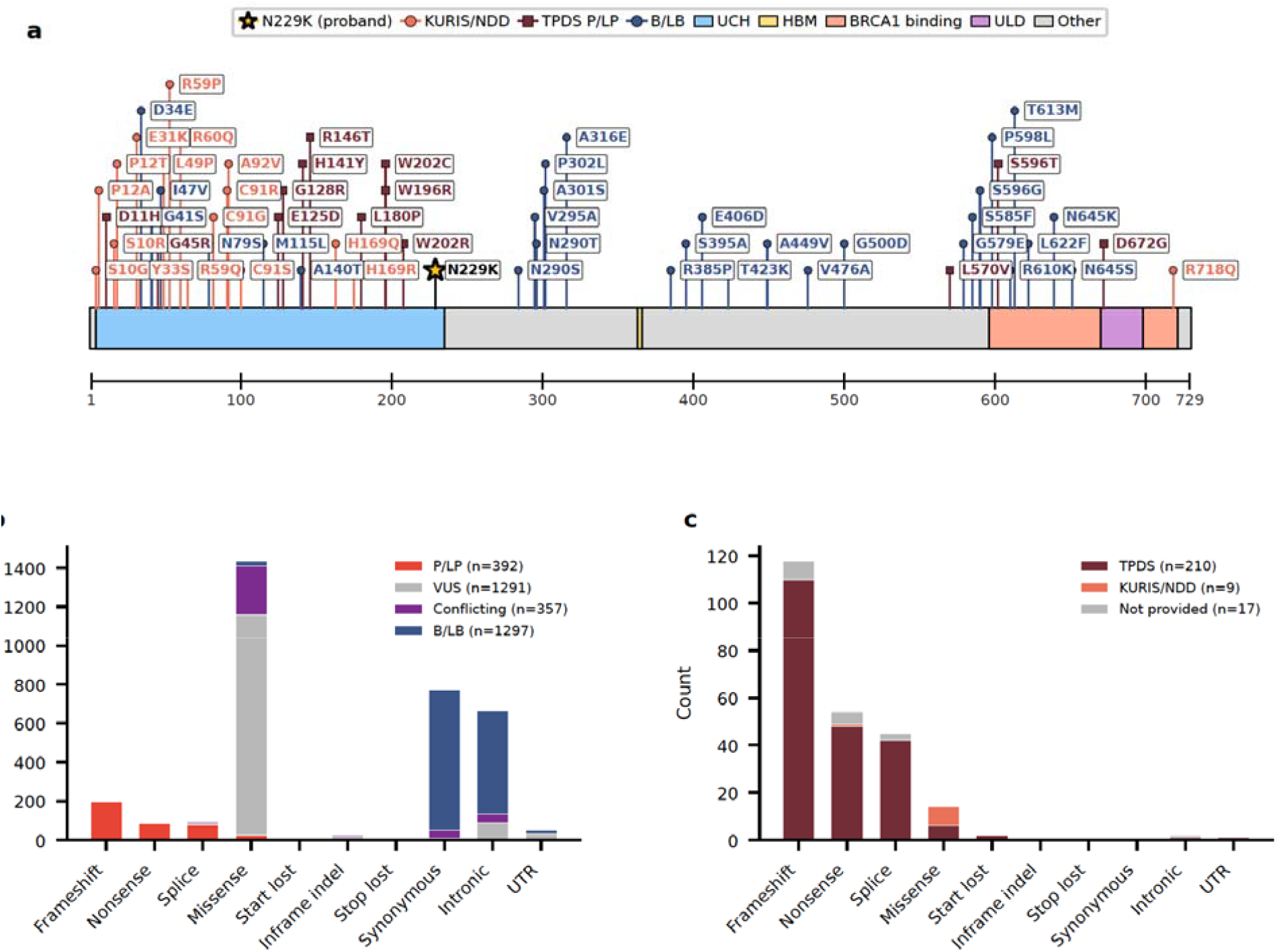
The molecular landscape of *BAP1* variants shows that most ClinVar P/LP variants are truncating variants associated with TPDS and that KURIS associated variants are primarily missense. **a**. Lollipop plot showing missense variants associated with KURIS or TPDS. **b**,**c**. Histograms of ClinVar variants showing (b) all variants colored by classification and (c) pathogenic/likely pathogenic variants colored by disease association.

### *BAP1* Functional Data Calibration for *BAP1*-TPDS

Although many assays of *BAP1* function exist, most published data sets only include a handful of variants, usually after a variant has been identified in an affected individual ^17,18,19,20,5,21,22,23,24,15,25,26,27^. Recently, Waters *et al*. measured the effects of ∼18,000 distinct *BAP1* variants using a large-scale saturation genome editing (SGE) functional assay^7^ (**Fig. 2a**). Here, libraries of variants were introduced into a haploid cell line dependent on *BAP1* for survival. Cell fitness is a surrogate for *BAP1* function, with each variant assigned a functional score representing its change in frequency over time relative to wild-type. This readout captures whether a variant disrupts *BAP1* function but does not distinguish between the specific molecular mechanisms of disruption. The assay accurately identified ClinVar P/LP variants as loss-of-function and B/LB variants as unchanged with >99% sensitivity and >98% specificity.

**Figure 2:**
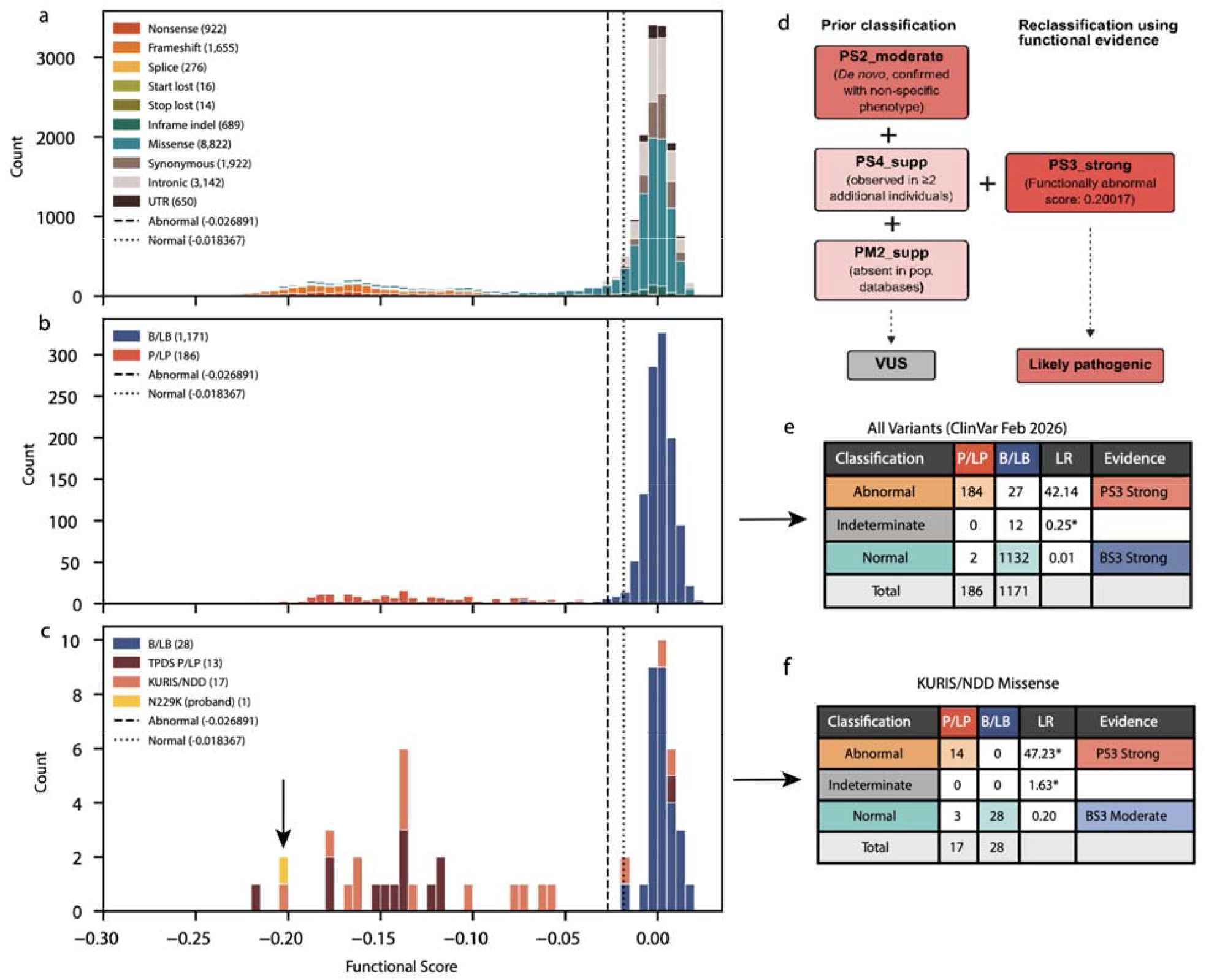
KURIS specific calibration of *BAP1* SGE assay reveals strong evidence for pathogenicity. Histograms of functional scores colored by (A) molecular consequence and (B) ClinVar classification. (C) Histogram of functional scores for missense variants only colored by disease association, with the arrow indicating variant p.N229K. (D) Reclassification of p.N229K from VUS to likely pathogenic. Prior evidence (PS2_moderate, PS4_supporting, PM2_supporting) combined with SGE data (PS3_strong) supported LP classification per ACMG/AMP guidelines. Contingency tables showing evidence strength calculated for (E) all ClinVar variants and (F) missense variants known to be associated with KURIS and benign missense variants from ClinVar.

Waters *et al*. originally calibrated their SGE assay using 2,423 truncating variants before the final exon as presumed pathogenic controls and 138 high population frequency variants as presumed benign controls, yielding OddsPath_Abnormal 27.6 (PS3_strong) and OddsPath_Normal = 0.002 (BS3_VeryStrong). This calibration uses a presumed loss-of-function truth set rather than clinically classified variants. Tejura et al.^28^ subsequently recalibrated this dataset using ClinVar P/LP and B/LB variants, which are predominantly related to cancer predisposition, and obtained OddsPath_Abnormal = 15.18 (PS3_Moderate) and OddsPath_Normal = 0.0116 (BS3_Strong). Applying our GMM-derived functional thresholds to a truth set with only definitively classified variants predominately related to cancer predisposition (ClinVar February 2026; n=186 P/LP, n=1,171 B/LB) yielded OddsPath_Abnormal = 27.6 (PS3_Strong) and OddsPath_Normal = 0.01 (BS3_Strong) (**Fig. 2b,e**). Regardless of calibration method, all three approaches demonstrate that the assay provides informative evidence for *BAP1*-TPDS. However, none of these calibrations address whether the assay is appropriately calibrated for KURIS.

### *BAP1* Functional Data Calibration for KURIS

Prior calibrations of the *BAP1* SGE dataset have relied on ClinVar pathogenic controls dominated by loss-of-function truncating variants associated with *BAP1*-TPDS, without distinguishing whether pathogenic variants were classified in the context of cancer predisposition or neurodevelopmental disease. This is a critical limitation. *BAP1*-TPDS is clearly related to overall loss-of-function, as premature stop codons and splice site alterations are the dominant variant class for *BAP1*-TPDS pathogenic variants. The *BAP1* SGE cellular fitness assay correctly measures these variants as loss-of-function. However, only missense variants are linked to KURIS. Consequently, the pathogenic mechanism for KURIS may be distinct from *BAP1*-TPDS, a difference that may not be captured by the *BAP1* SGE cellular fitness assay.

Although Waters *et al*. previously showed that *BAP1* variants reported in individuals with neurodevelopmental phenotypes were depleted in the SGE assay at magnitudes comparable to cancer-associated variants, prior calibrations have not formally established the evidence strength this assay provides in the KURIS context. We therefore asked whether the assay’s functional readout, while mechanism-agnostic, could nonetheless provide appropriately calibrated evidence for KURIS variants when benchmarked against a KURIS-specific truth set.

To calibrate the *BAP1* KURIS-specific cutoffs, we utilized a set of 17 *de novo* missense variants with well-characterized phenotypic information as pathogenic controls and 28 B/LB missense variants in ClinVar. Of the 17 KURIS controls, 14 were classified as functionally abnormal and 3 as functionally normal. Among the 28 B/LB variants, 27 were normal, and 1 was abnormal. This yielded OddsPath_Abnormal = 47.23 (PS3_Strong) and OddsPath_Normal = 0.20 (BS3_Moderate), demonstrating that the SGE assay has strong predictive value for pathogenicity when appropriately benchmarked to a KURIS-specific truth set (**Fig. 2c,f**). These phenotype-specific calibrations have been integrated into MaveMD^29^ to enable the broader use of these datasets.

While the KURIS-specific calibration demonstrates that the SGE assay provided strong evidence of pathogenicity in the context of neurodevelopmental disorders, evidence for benignity was reduced to moderate. The reduced BS3 evidence strength reflects the observation that three KURIS-associated variants (p.Arg59Gln, p.Arg60Gln, p.Arg718Gln) scored as functionally normal in the SGE assay, suggesting that some KURIS-associated variants may act through mechanisms not captured by the cell-fitness phenotype. Of note, p.Arg718Gln is outside of the UCH domain and was shown by Küry *et al*. to preserve deubiquitinase activity in a complementation assay. Moreover, it is present in three heterozygotes in gnomADv4, suggesting that this variant may not be related to neurodevelopmental disease.

### Application of KURIS-specific *BAP1* Calibration for Variant Reclassification

We integrated functional evidence from the KURIS-specific calibration to classify a germline *de novo* missense variant within the UCH catalytic domain of *BAP1*, NM_004656.4:c.687C>A (p.Asn229Lys) identified in a pediatric proband. This individual was referred to the UDN due to a history of neurodevelopmental and ocular symptoms. Although the *BAP1* variant was classified as a VUS, the individual’s clinical phenotype overlapped with KURIS, including global developmental delay, hypotonia, dysmorphic features, limb anomalies, and sensory involvement (**Supplemental Note 1**). Following ACMG/AMP and ClinGen guidance, we confirmed the original VUS classification, applying criteria; PS2_moderate (confirmed *de novo* in the current proband with a consistent but non-specific phenotype), PS4_supporting (observed in at least two additional unrelated individuals with a neurodevelopmental phenotype^30,31^, ClinVar SCV003799185.3), and PM2_supporting (absent from large population databases). This *BAP1* VUS was classified as functionally abnormal in the *BAP1* SGE assay (**Fig. 2c**), therefore providing PS3_Strong evidence according to the KURIS-specific calibration. Applying this additional strong evidence to existing supporting and moderate pathogenic evidence reclassified this variant as likely pathogenic (**Fig 2d**).

### Broader Implementation of Disease-Specific Recalibration of Functional Data

Disease-specific recalibration of functional data has the potential to be broadly useful in clinical genomics, and emerging gene and variant-specific calibration approaches can further refine clinical variant classification^32,33^. Even though MAVE datasets are being generated at an accelerating pace^28,34,35^, translation of this data into clinical practice remains a bottleneck. A recent survey reported that 36% of respondents had low confidence in using high-throughput functional data, while 83% indicated that they would benefit from targeted training opportunities at professional meetings^36^. Given this, we generated and evaluated an educational workshop that demonstrates how MAVE functional data can be translated into disease-specific functional evidence for clinical variant classification. This workshop is framed around the *BAP1* VUS and the *BAP1* disease-specific reclassifications presented above, and guides participants through different assay types, the ClinGen SVI framework for evaluating assay validity, calculating likelihood ratios using appropriate reference sets, and applying calibrated functional data as evidence for variant classification (**Supplemental Note 2**). We deployed this workshop at the 2026 American College of Medical Genetics and Genomics (ACMG) annual meeting, and a pre-workshop survey of participants (n=114) showed that 82% attendees (n=93) were initially only slightly or not at all familiar with MAVEs. Notably, in the post-workshop survey (n=65), the percentage of participants reporting “comfortable or very comfortable” rose from 23% to 43%, and those reporting “not at all comfortable” fell to 0% (**Supplemental Fig. 2**).

## Discussion

Multiplexed assays of variant effect (MAVEs) continue to expand across disease-associated genes^35^, and SGE has emerged as a particularly powerful approach for generating variant effect maps at single-nucleotide resolution^37,38, 39^. However, most SGE assays, including the Waters *et al. BAP1* assay applied here, measure variant effects on cell fitness, a readout that largely captures loss-of-function, but is nonetheless agnostic to the underlying molecular mechanism by which loss-of-function is achieved^40^. This limitation becomes consequential when a gene causes multiple disorders through potentially distinct mechanisms, raising the question of how functional data generated in one clinical context can be appropriately applied to another.

The *BAP1* SGE assay was originally developed and calibrated in the context of cancer predisposition, where known pathogenic variants are overwhelmingly truncating variants. Against ClinVar controls, composed almost entirely of *BAP1*-TPDS variants, the assay provided PS3_Strong evidence for pathogenicity. However, the *BAP1* VUS in our study was identified in the context of a neurodevelopmental phenotype (KURIS), which has only been described with *de novo* missense variants. Recalibrating the assay with a truth set of 17 KURIS-associated missense variants and 28 ClinVar B/LB missense controls independently yielded PS3_Strong evidence for the neurodevelopmental context. This enabled the reclassification of the proband’s variant from VUS to likely pathogenic.

Currently, 12-44% of human disease genes are associated with multiple phenotypes^2,41^. A recent phenome-wide association study of 23 hereditary cancer genes identified 19 previously unrecognized disease associations, of which 12 were non-neoplastic^42^, highlighting that the clinical spectrum of many genes extends well beyond their standard disease associations. Our findings underscore a practical gap in how functional assays are currently calibrated for pleiotropic genes: Brnich *et al*.^*9*^ state “For genes associated with multiple disorders through different mechanisms, an assay validated for one disorder may not necessarily be applied universally to analyze the variant effect in other disorders if the mechanisms of the disease are different.” In practice, however, functional assays are usually calibrated against all ClinVar P/LP and B/LB controls, which reflects whichever disorder dominates clinical testing and ClinVar submissions. For *BAP1*, a calibration based on a ClinVar truth set is effectively a calibration for *BAP1*-TPDS. We show that the typical lumped approach correctly yields PS3_Strong evidence for *BAP1*-TPDS, but over-weights benign evidence when applied to KURIS (BS3_Strong rather than BS3_Moderate). This more conservative BS3 strength may reflect the underlying biology of the KURIS truth set. Three variants (p.Arg59Gln, p.Arg60Gln, p.Arg718Gln) were classified as functionally normal in the SGE assay. Some KURIS-associated variants may therefore act through mechanisms not captured by a cell-fitness readout, or may represent a partial loss of function not sufficient to cause a growth deficit in HAP1 cells.

Phenotype-specific calibration can extend the utility of existing high-quality MAVE datasets across multiple clinical contexts, avoiding the need to generate a new assay for every gene-disease association. Not all genes, however, are suitable for cross-context reuse. A similar scope-of-calibration caveat has recently been made explicit for *SCN5A* (HGNC:10593), where a calibrated automated patch-clamp assay was restricted to Brugada syndrome 1 (OMIM:601144) despite it also causing other cardiac diseases including Long QT Type 3 (OMIM:603830) and dilated cardiomyopathy 1 E^43^ (OMIM:601154). The same concern applies to *PTEN* (HGNC:9588), where neurodevelopmental and autism-associated (OMIM: 605309) variants may act through mechanisms distinct from those seen in individuals with *PTEN* hamartoma-tumor syndrome (OMIM:158350)^44,45,46,47,48^. When underlying molecular mechanisms diverge between diseases, the assay’s validity for each context should be independently established.

In conclusion, our study provides a practical framework for applying functional evidence to pleiotropic genes with multiple clinical presentations, and presents an approach to extend the diagnostic utility of existing MAVE datasets to additional clinical contexts. Furthermore, we provide a validated clinical teaching workshop that can be adapted to curricula for variant review scientists, genetic counselors, and medical geneticists to enable uptake and understanding of MAVE data and their calibrations, a pressing need in the field^36^. As MAVE data accumulate and disease-specific clinical datasets expand, this approach offers a generalizable path to resolving variants of uncertain significance across the diverse phenotypic spectrum of pleiotropic genes.

## Data Availability

Source data for calibration figures and a standalone Python script for figure generation are available at https://github.com/amcewenmdphd/BAP1_KURIS_figures.

Our calibrations of the Waters *et al. BAP1* dataset (urn:mavedb:00000662-0-1) using the GMM-derived thresholds on all ClinVar variants from February 2026 and KURIS/NDD associated variants have been added as an additional calibration that can be referenced.

## Acknowledgments

The study team is deeply grateful to the participant and her family for their participation in the Undiagnosed Diseases Network.

## Author Contributions

Conceptualization: P.G., G.P.J., K.M.D., E.E.B., D.M.F., L.M.S., A.B.S., A.E.M.; Methodology: P.G., D.M.F., L.M.S., A.B.S., A.E.M.; Software: J.S., E.A.R.; Validation: E.E.B.; Formal Analysis: P.G., M.T., M.S., A.E.M.; Investigation: P.G., E.V.B., R.D.K., R.M.V., K.A.L., M.B., M.M., A.B.S., A.E.M.; Resources: P.G., E.V.B., J.S., M.B., M.M., G.P.J., K.M.D., E.E.B., A.B.S., A.E.M.; Data Curation: P.G., M.T., M.S., A.E.M.; Writing-original draft: P.G., R.D.K., M.S., R.M.V., A.E.M.; Writing-review & editing: P.G., E.V.B., G.P.J., K.M.D., E.E.B., D.M.F., L.M.S., A.B.S., A.E.M.; Visualization: P.G., M.S., A.E.M.; Supervision: P.H.B., S.C., I.A.G., D.E.M., A.K.S., V.P.S., M.W., D.M.F., L.M.S., A.B.S., A.E.M.; Project Administration: M.H.-P., J.R., G.P.J., K.M.D., E.E.B.; Funding Acquisition: UDN, G.P.J., K.M.D., E.E.B., D.M.F., L.M.S., A.B.S.

## Ethics Declaration

The participant was enrolled in the Undiagnosed Diseases Network (UDN) research study, approved by the National Institutes of Health Institutional Review Board (IRB# 15HG0130). The legal guardians provided written consent for research participation.

## Supplementary information

**Supplemental Figure 1:**
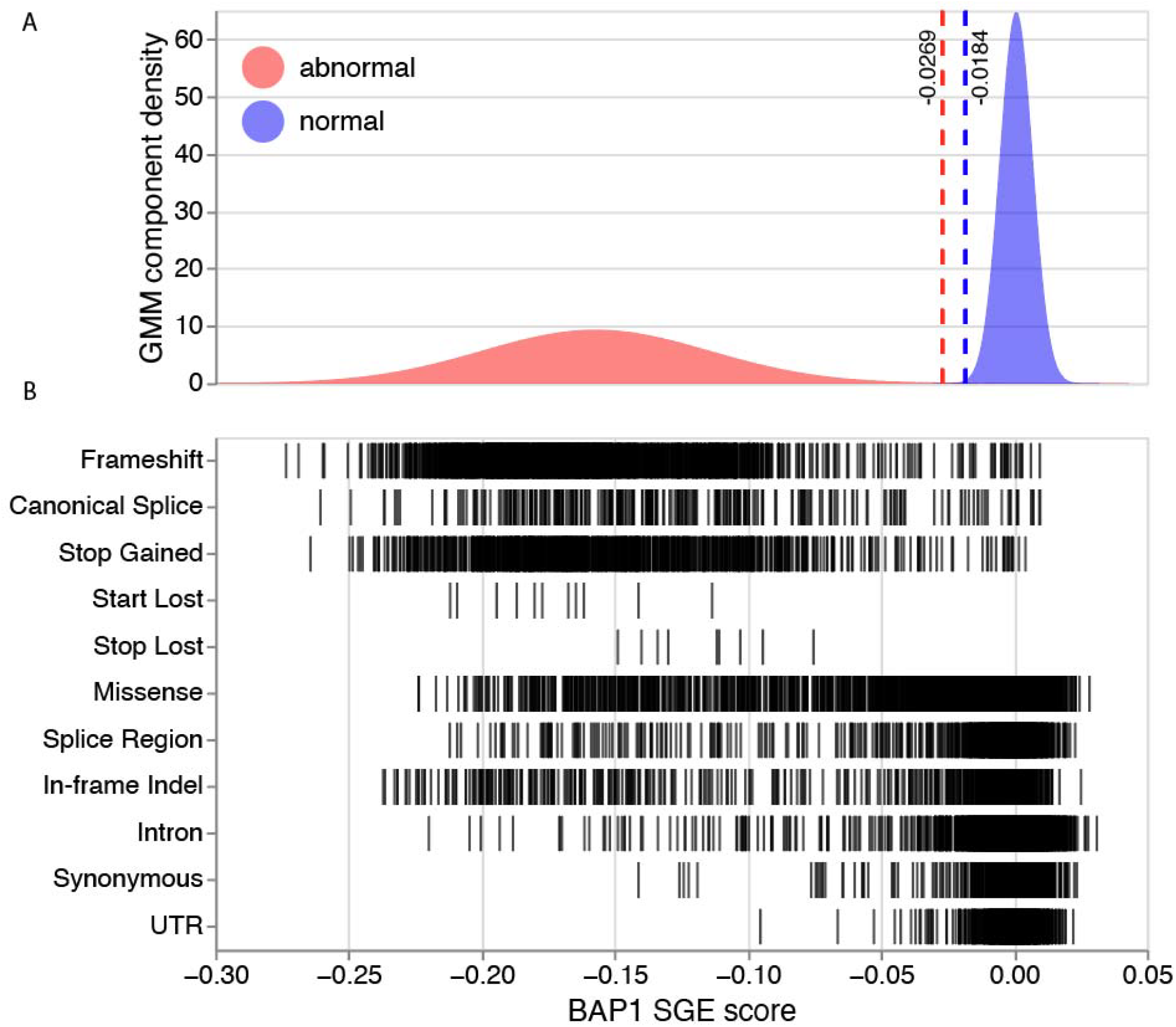
Fitted gaussians for variant functional classification. (A) “Abnormal” (red) and “Normal” (blue) gaussians used to draw thresholds for classifying variants as functionally abnormal or functionally normal. Estimated density from GMM-modeling is on the Y-axis, and SGE score is on the X-axis. Vertical red dashed line at X = -0.0269 represents upper threshold for functionally abnormal variants. Vertical blue dashed line at X = -0.0184 represents lower threshold for functionally normal variants. (B) Strip plot of functional class (Y-axis) vs. SGE score (X-axis)

**Supplemental Figure 2:**
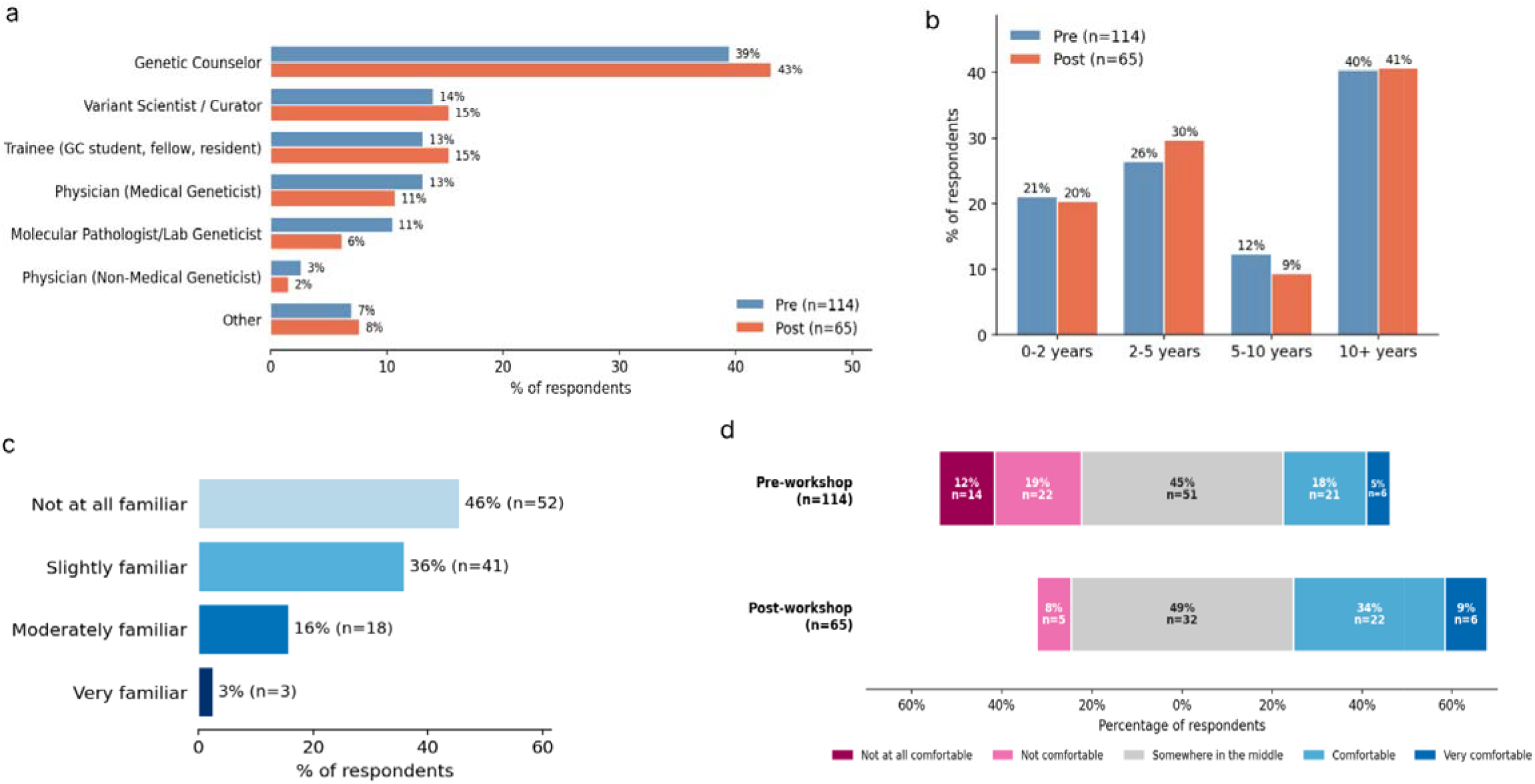
Pre- and post-workshop survey results on utilizing functional data for variant classification. Breakdown of (A) professional roles and (B) duration of professional experience of workshop attendees (C) Respondents’ familiarity with MAVEs (D) Workshop attendees’ confidence in using functional evidence for the purpose of variant classification.

**Supplementary Table 1: Calibration Controls**

**Supplementary Table 2.**
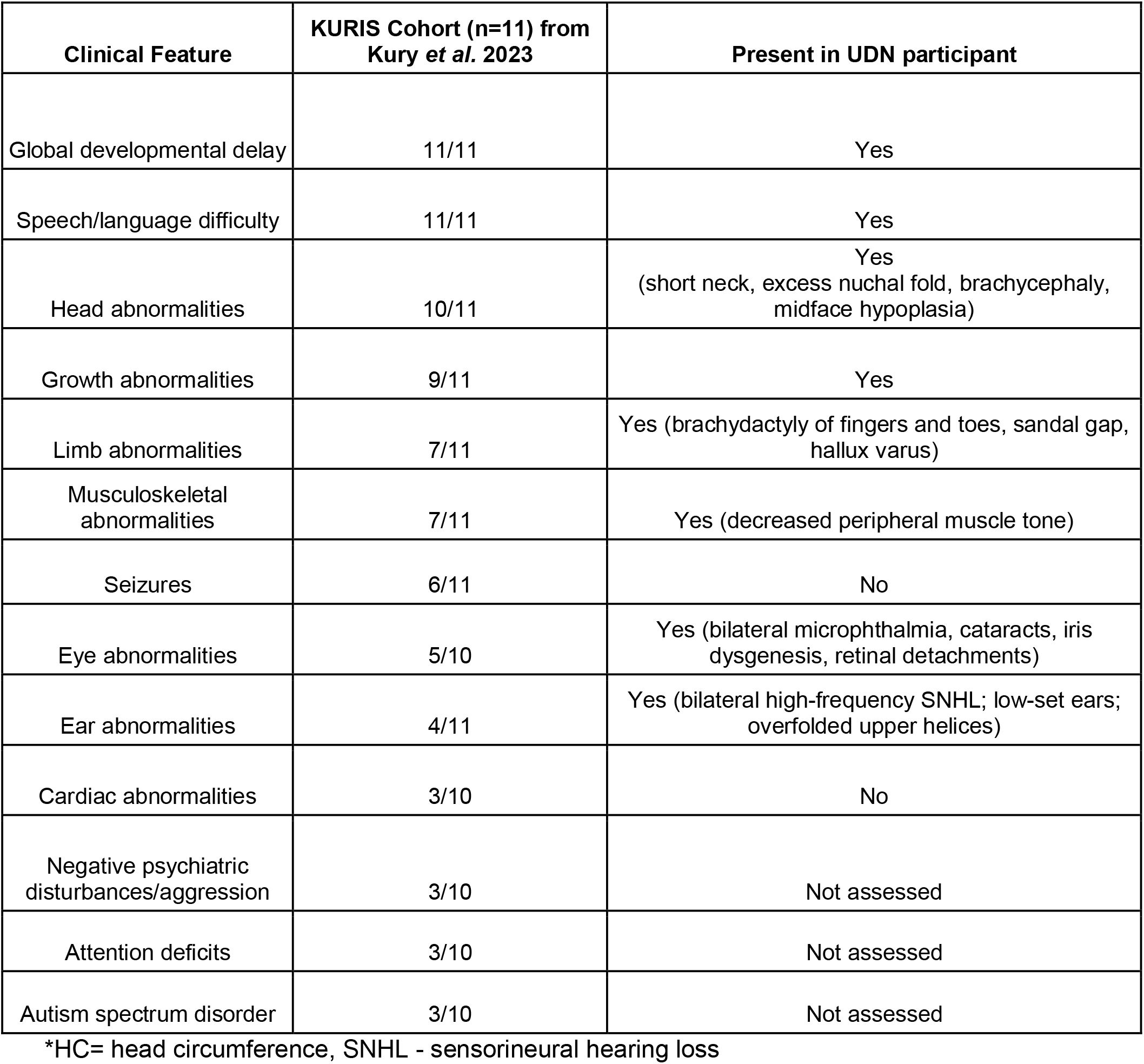
Phenotypic overlap with proband and reported KURIS features.

**Supplemental note 1:** Case Presentation

### Clinical Presentation

A pediatric individual was referred to the Undiagnosed Diseases Network (UDN) for evaluation of congenital ocular anomalies, global developmental delay and dysmorphic features. The individual was born via spontaneous vaginal delivery with no complications other than a nuchal umbilical cord requiring brief resuscitation.

In infancy, the individual was noted to have bilateral leukocoria, bilateral microphthalmia, iris dysgenesis, congenital cataracts, and bilateral funnel retinal detachments. Audiologic evaluation confirmed mild bilateral high-frequency sensorineural hearing loss. Dysmorphic features were also noted including a short neck with excess nuchal fold, brachydactyly of the hands and feet, sandal gap, and bilateral hallux varus. The individual exhibited mild to moderate global developmental delay with hypotonia.

### Prior Workup

Initial diagnostic workup included a normal chromosomal microarray arr(1-22,X)x2 and negative screen for congenital TORCH infection. Trio exome sequencing with mitochondrial genome analysis identified a *de novo* variant of uncertain significance (VUS) in *AGAP1* (HGNC:16922), NM_014914.5: c.1208T>C (p.Val403Ala). This variant was deprioritized because it was observed twice in healthy individuals (gnomADv4 AF = 2.85-06) and is not known to be associated with human disease.

### Workup as part of the UDN

As part of the UDN evaluation, trio genome sequencing was performed on buccal cells which identified a germline *de novo* missense VUS in *BAP1*, NM_004656.4:c.687C>A (p.Asn229Lys). This variant lies within the UCH catalytic domain of *BAP1*, is absent from population databases, has been reported in ClinVar (Variation ID: 2429331), and has been observed in two other individuals with neurodevelopmental disease (PMID 31785789, ClinVar SCV003799185.3).

### Phenotypic Concordance with KURIS

This individual’s clinical features substantially overlap with KURIS. Core features present in all reported KURIS individuals, global developmental delay and speech/language difficulty, were evident in this individual (Supplemental Table 2). The ocular phenotype in this individual was notably more severe than previously reported in KURIS. While eye abnormalities have been observed in approximately half of reported individuals, prior descriptions included refractive errors or mild abnormalities. This individual presented with congenital bilateral microphthalmia, cataracts, iris dysgenesis, and bilateral funnel retinal detachments, features not previously described in this syndrome.

Features reported in KURIS but absent in this individual include seizures, and cardiac abnormalities. Behavioral and psychiatric features reported in subsets of KURIS individuals, including autism spectrum disorder, attention deficits, and aggression, have not been formally assessed.

**Supplemental note 2:** Education workshop slides

### Declaration of AI and AI-assisted technologies in the writing process

During the preparation of this work the authors used Claude (Anthropic) in order to proofread and edit the text for clarity and grammar. After using this tool/service, the authors reviewed and edited the content as needed and take full responsibility for the content of the publication.

