## Supplemental Table 2 for "Phenotype-Specific Recalibration of MAVE Data Enables Repurposing of *BAP1* Functional Assays for Küry-Isidor Syndrome"

**Supplementary Table 2.** Phenotypic overlap with proband and reported KURIS features

| Clinical Feature | KURIS Cohort (n=11) from Kury *et al.* 2023 | Present in UDN participant |
|---|---|---|
| Global developmental delay | 11/11 | Yes |
| Speech/language difficulty | 11/11 | Yes |
| Head abnormalities | 10/11 | Yes (short neck, excess nuchal fold, brachycephaly, midface hypoplasia) |
| Growth abnormalities | 9/11 | Yes |
| Limb abnormalities | 7/11 | Yes (brachydactyly of fingers and toes, sandal gap, hallux varus) |
| Musculoskeletal abnormalities | 7/11 | Yes (decreased peripheral muscle tone) |
| Seizures | 6/11 | No |
| Eye abnormalities | 5/10 | Yes (bilateral microphthalmia, cataracts, iris dysgenesis, retinal detachments) |
| Ear abnormalities | 4/11 | Yes (bilateral high-frequency SNHL; low-set ears; overfolded upper helices) |
| Cardiac abnormalities | 3/10 | No |
| Negative psychiatric disturbances/aggression | 3/10 | Not assessed |
| Attention deficits | 3/10 | Not assessed |
| Autism spectrum disorder | 3/10 | Not assessed |

*HC= head circumference, SNHL - sensorineural hearing loss
