## Supplemental Note 2 for "Phenotype-Specific Recalibration of MAVE Data Enables Repurposing of *BAP1* Functional Assays for Küry-Isidor Syndrome"

### BEYOND UNCERTAINTY: USING FUNCTIONAL ASSAYS FOR CLINICAL VARIANT CLASSIFICATION

### CLINGEN GUIDANCE ON TRANSLATING FUNCTIONAL DATA INTO FUNCTIONAL EVIDENCE

ClinGen
Clinical Genome Resource

Atlas of Variant Effects

### VARIANT **CLASSIFICATION**

#### PROBABILITY THAT A VARIANT *CAN* CAUSE DISEASE

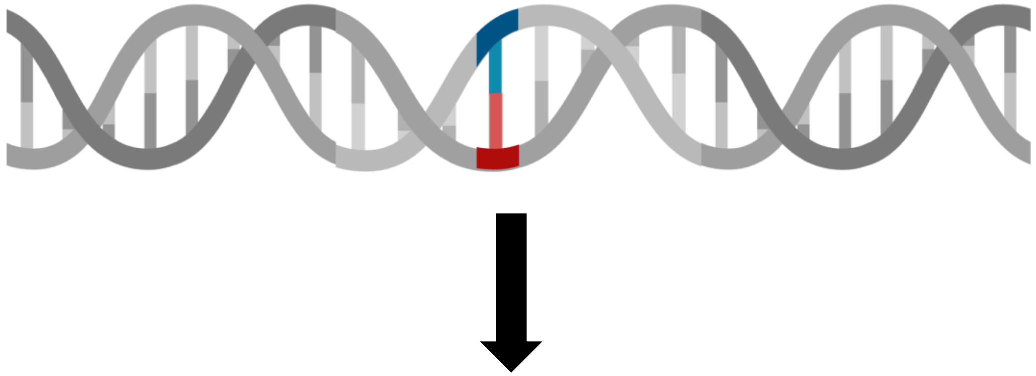

| Benign | Likely Benign | Uncertain | Likely Pathogenic | Pathogenic |
|---|---|---|---|---|
| $P < 0.001$ | $0.001 \leq P < 0.10$ | $0.10 \leq P \leq 0.90$ | $0.90 < P \leq 0.99$ | $P > 0.99$ |

PMID: 31479589 (Richards *et al.* 2015)

### VUS ARE INCREASING EXPONENTIALLY

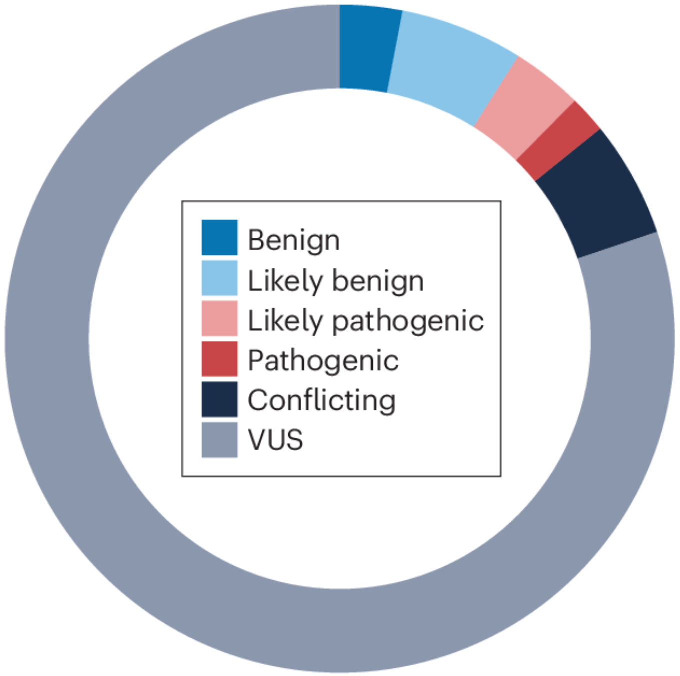

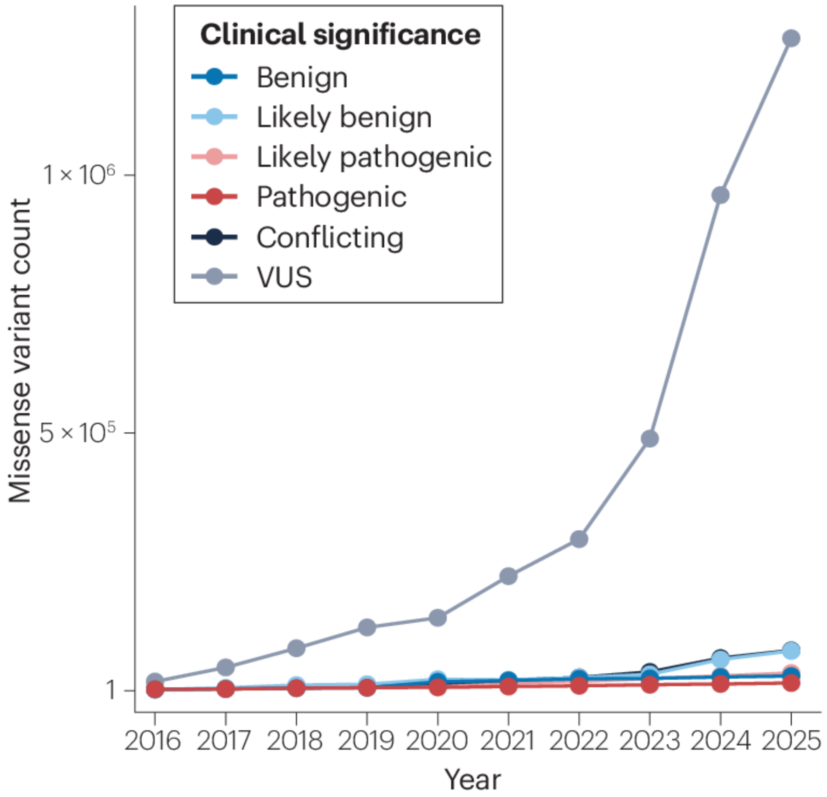

McEwen et al. 2025 (PMID: 40691352)

### VARIANT CLASSIFICATION GUIDELINES
### AKA "RICHARDS RULES"

© American College of Medical Genetics and Genomics

ACMG STANDARDS AND GUIDELINES | Genetics inMedicine

#### Standards and guidelines for the interpretation of sequence variants: a joint consensus recommendation of the American College of Medical Genetics and Genomics and the Association for Molecular Pathology

Sue Richards, PhD[1], Nazneen Aziz, PhD[2,16], Sherri Bale, PhD[3], David Bick, MD[4], Soma Das, PhD[5], Julie Gastier-Foster, PhD[6,7,8], Wayne W. Grody, MD, PhD[9,10,11], Madhuri Hegde, PhD[12], Elaine Lyon, PhD[13], Elaine Spector, PhD[14], Karl Voelkerding, MD[13] and Heidi L. Rehm, PhD[15]; on behalf of the ACMG Laboratory Quality Assurance Committee

### Data Sources for Variant Classification

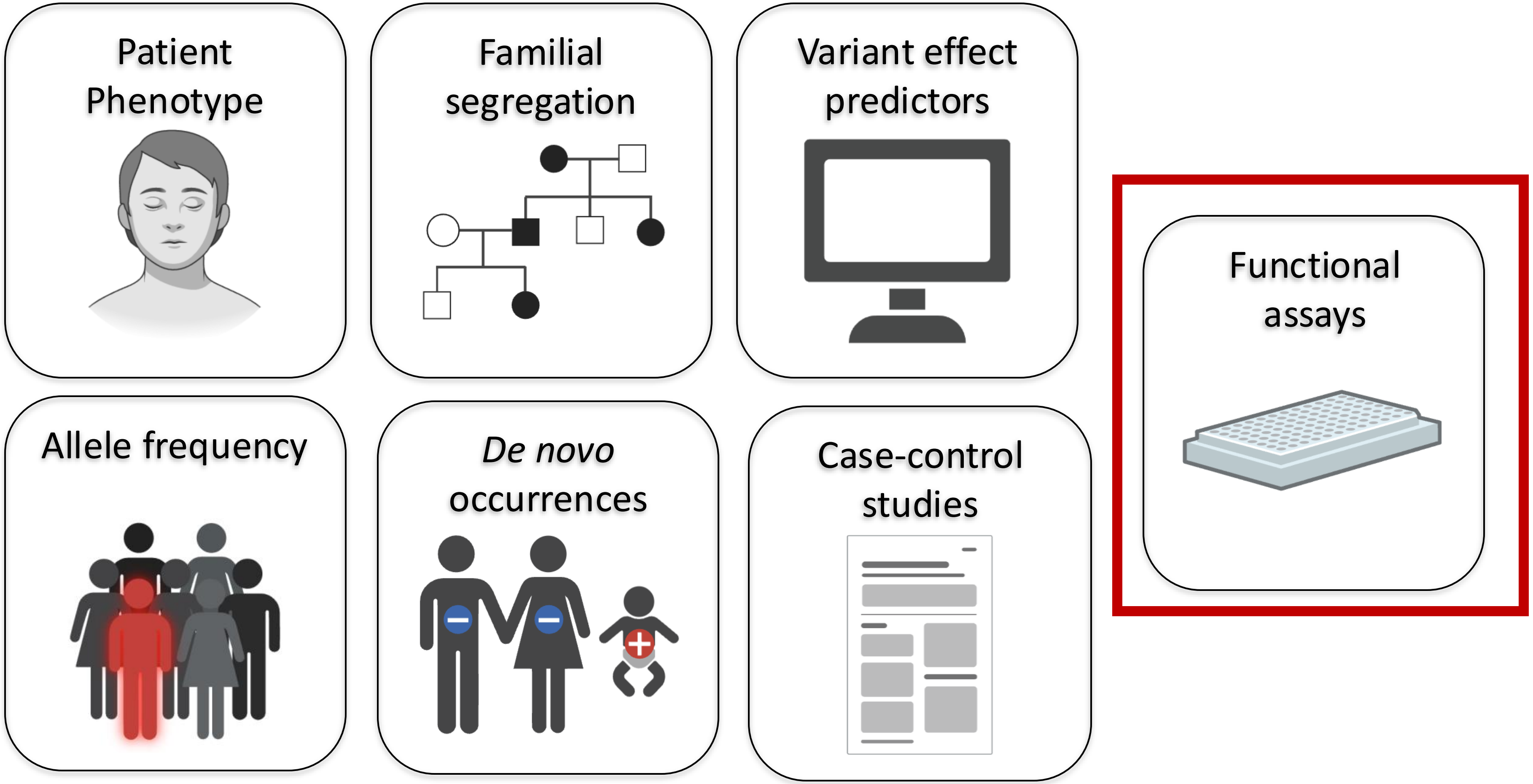

| Evidence Strength | | | |
|---|---|---|---|
| | Benign Strong | Benign Moderate | Benign Supporting |
| | Pathogenic Strong | Pathogenic Moderate | Pathogenic Supporting |

### VARIANT CLASSIFICATION DETERMINED BY COMBINING EVIDENCE

| | | | | | | |
|---|---|---|---|---|---|---|
| Pathogenic | PVS | PS | | | | |
| | PVS | PM | PM | | | |
| | PVS | PM | PP | | | |
| | PS | PS | | | | |
| | PS | PM | PM | PM | | |
| | PS | PM | PP | PP | | |
| | PS | PM | PP | PP | PP | PS |
| Likely Pathogenic | PVS | PM | | | | |
| | PS | PM | PM | | | |
| | PS | PM | | | | |
| | PS | PP | PP | | | |
| | PM | PM | PM | | | |
| | PM | PM | PP | PP | | |
| | PM | PP | PP | PP | PP | |
| Uncertain Significance | Insufficient or conflicting evidence | | | | | |
| Likely Benign | BS | BP | | | | |
| | BP | BP | | | | |
| Benign | BA | | | | | |
| | BS | BS | | | | |

Modified from Brnich 2018 (PMID: 30095857)

### WHAT IS FUNCTIONAL DATA?

Results from a ***laboratory experiment*** to determine how a variant affects protein or gene function

### Patient Specimens

a-gal assay for Fabry disease

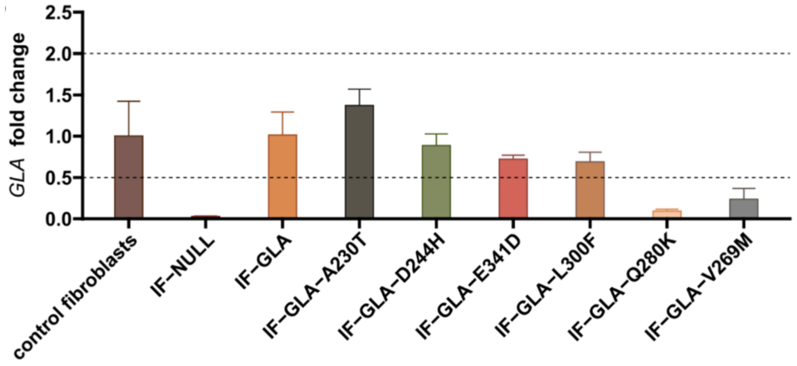

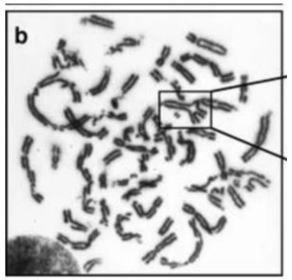

Mitomycin C assay to induce radial chromosomes for Fanconi anemia

Patient Phenotype

### Model Systems

Human tyrosyl-tRNA synthetase assessed in yeast

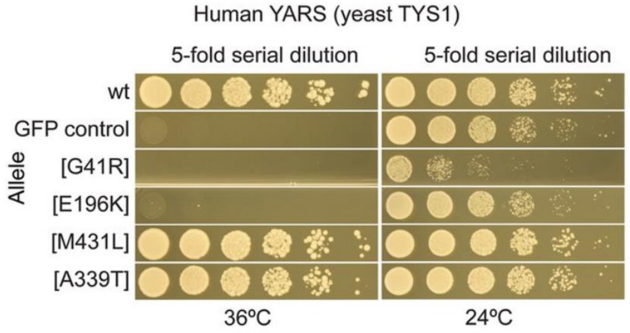

Sun et al. Genome Research 2016

Mouse enhancer deletion recapitulates human phenotype

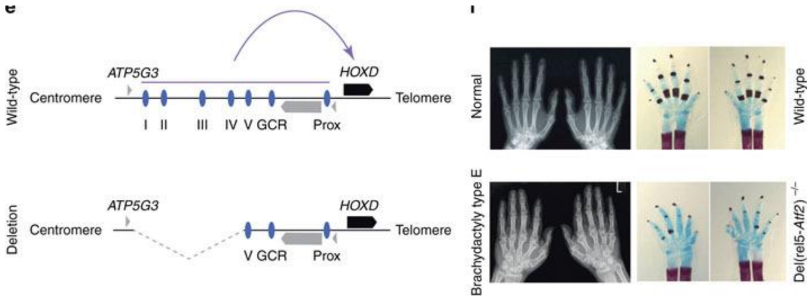

Functional Data

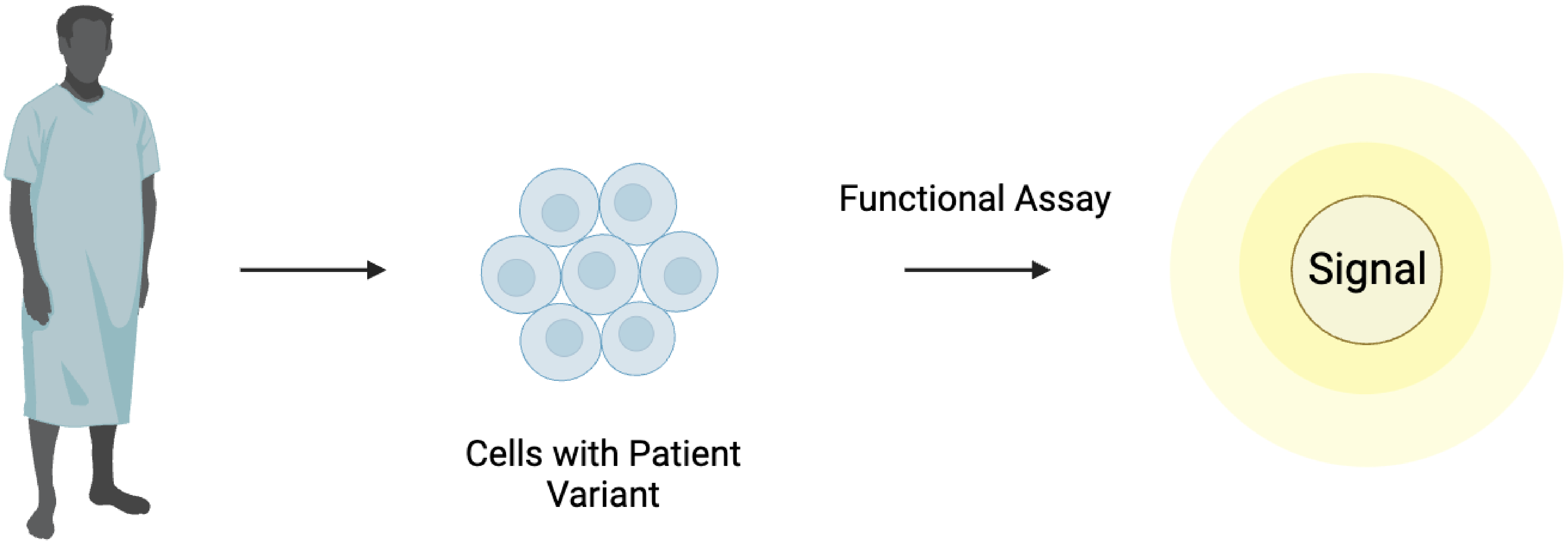

#### TRADITIONAL *IN VITRO* ASSAYS OF VARIANT PROTEIN FUNCTION: SLOW, LABORIOUS, EXPENSIVE

### TRADITIONAL ASSAYS DO NOT SCALE

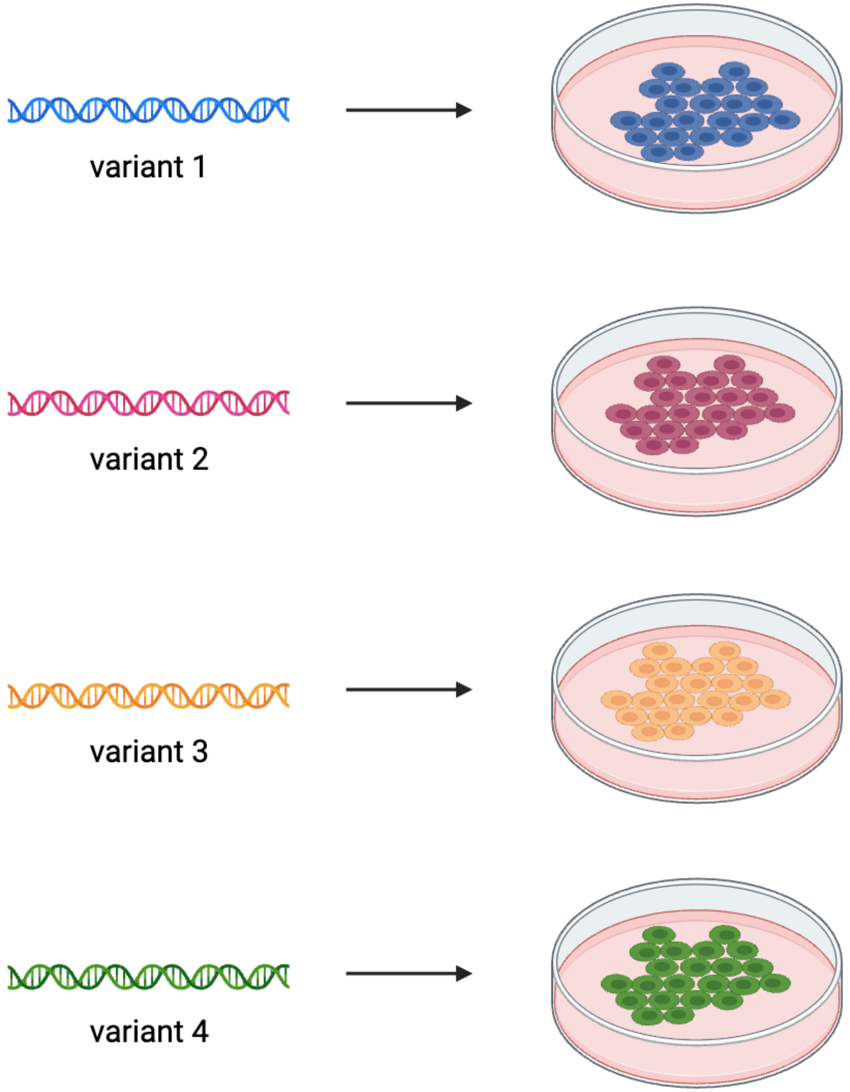

### WHAT IF THOUSANDS OF VARIANTS COULD BE STUDIED IN A SINGLE EXPERIMENT?

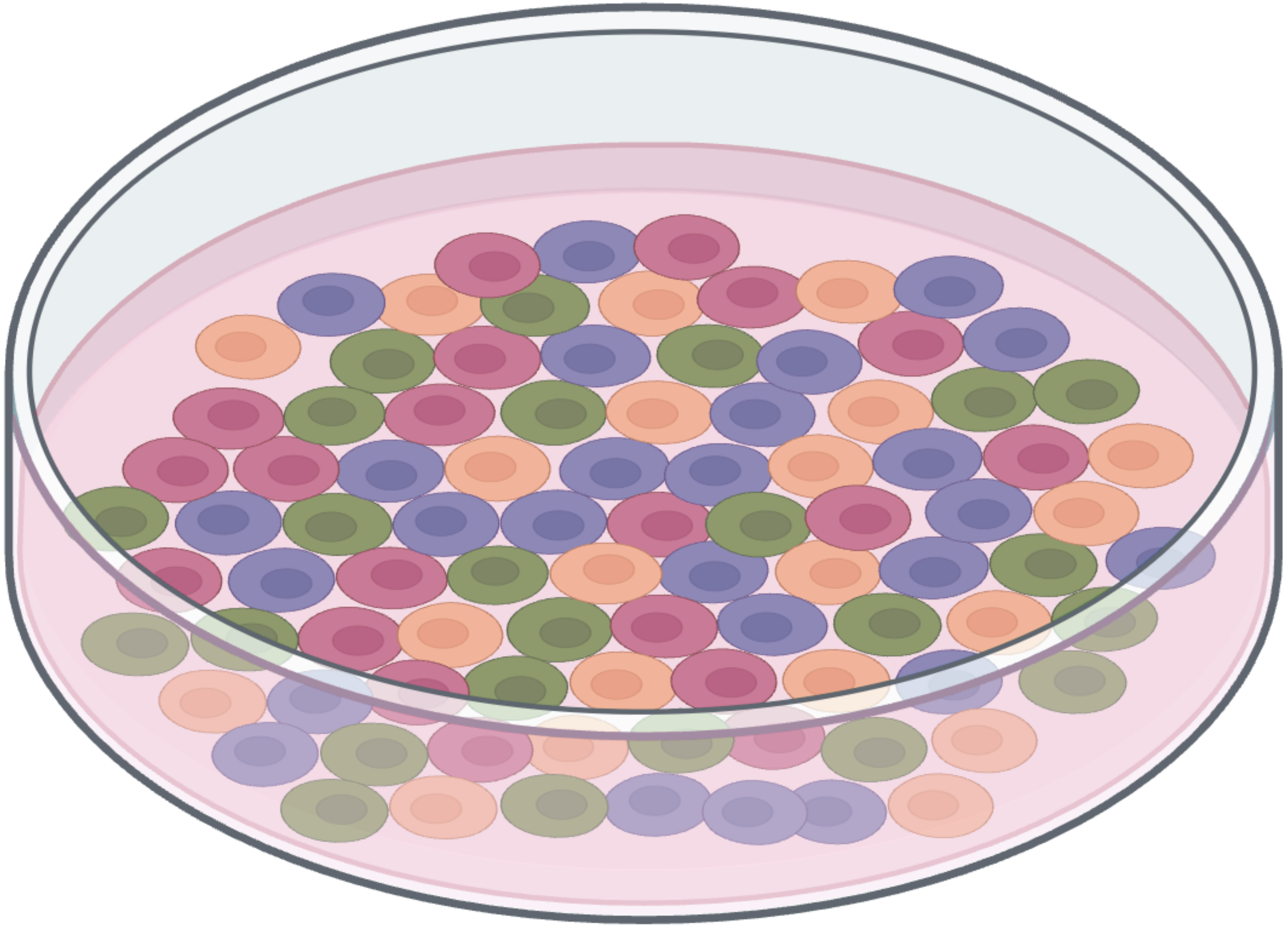

### MULTIPLEXED ASSAYS OF VARIANT EFFECT (MAVES) INVESTIGATE VARIANT FUNCTION AT SCALE

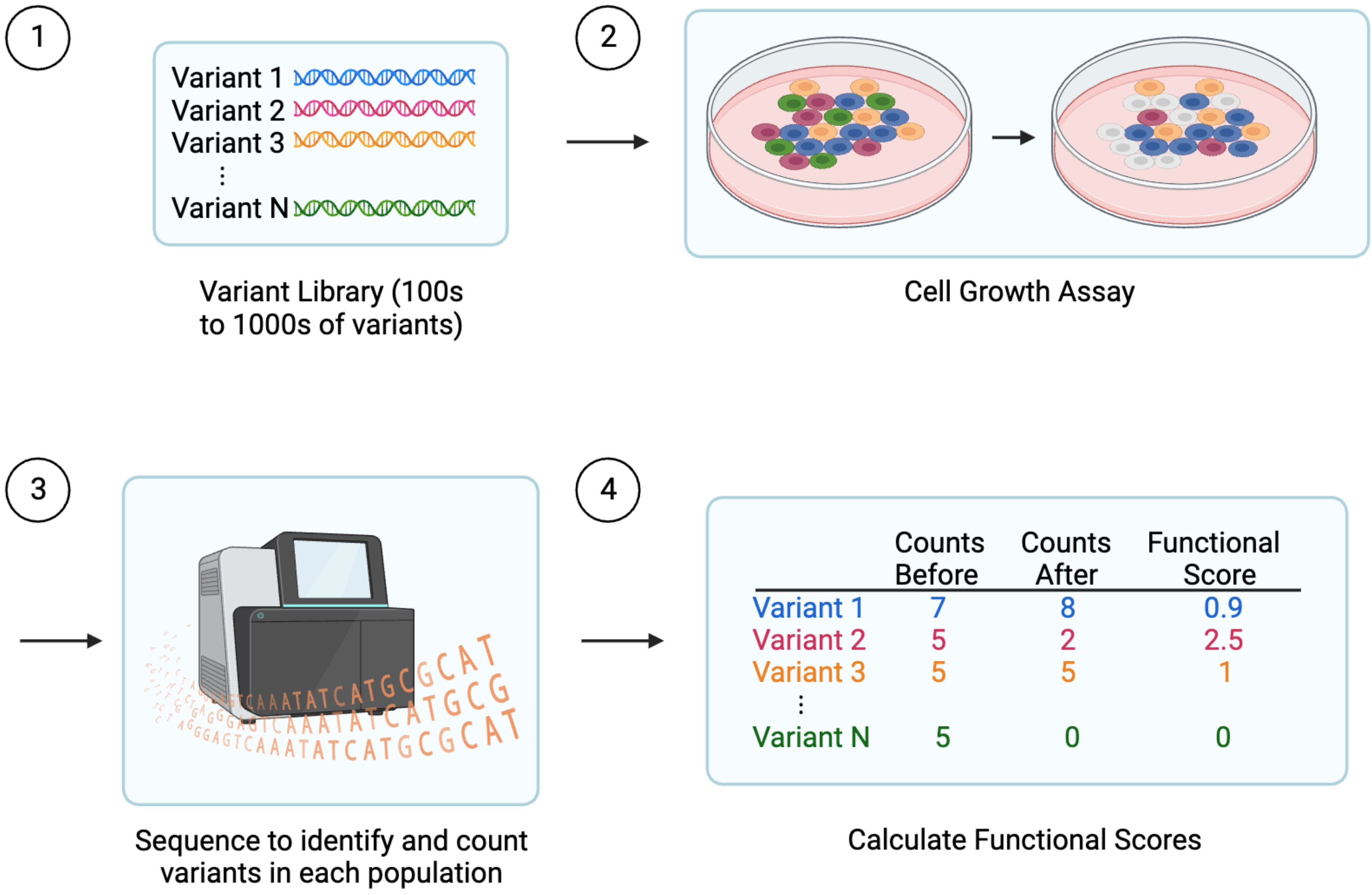

### MAVE DATA CAN BE USED TO GENERATE A VARIANT EFFECT MAP

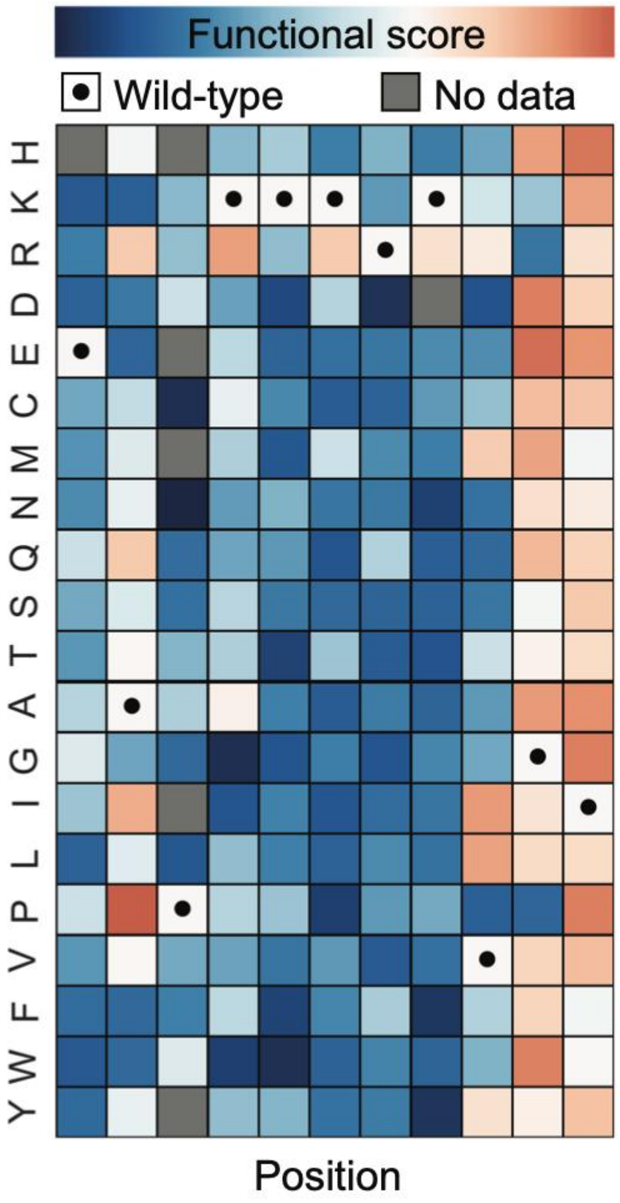

### CURRENT CLINGEN SVI GUIDELINES (2019)

Brnich *et al. Genome Medicine* (2020) 12:3
https://doi.org/10.1186/s13073-019-0690-2

Genome Medicine

GUIDELINE Open Access

#### Recommendations for application of the functional evidence PS3/BS3 criterion using the ACMG/AMP sequence variant interpretation framework

Sarah E. Brnich[1], Ahmad N. Abou Tayoun[2], Fergus J. Couch[3], Garry R. Cutting[4], Marc S. Greenblatt[5], Christopher D. Heinen[6], Dona M. Kanavy[1], Xi Luo[7], Shannon M. McNulty[1], Lea M. Starita[8,9], Sean V. Tavtigian[10], Matt W. Wright[11], Steven M. Harrison[12], Leslie G. Biesecker[13], Jonathan S. Berg[1*] and On behalf of the Clinical Genome Resource Sequence Variant Interpretation Working Group

### RECOMMENDATIONS FOR TRANSLATING FUNCTIONAL *DATA* INTO *EVIDENCE*

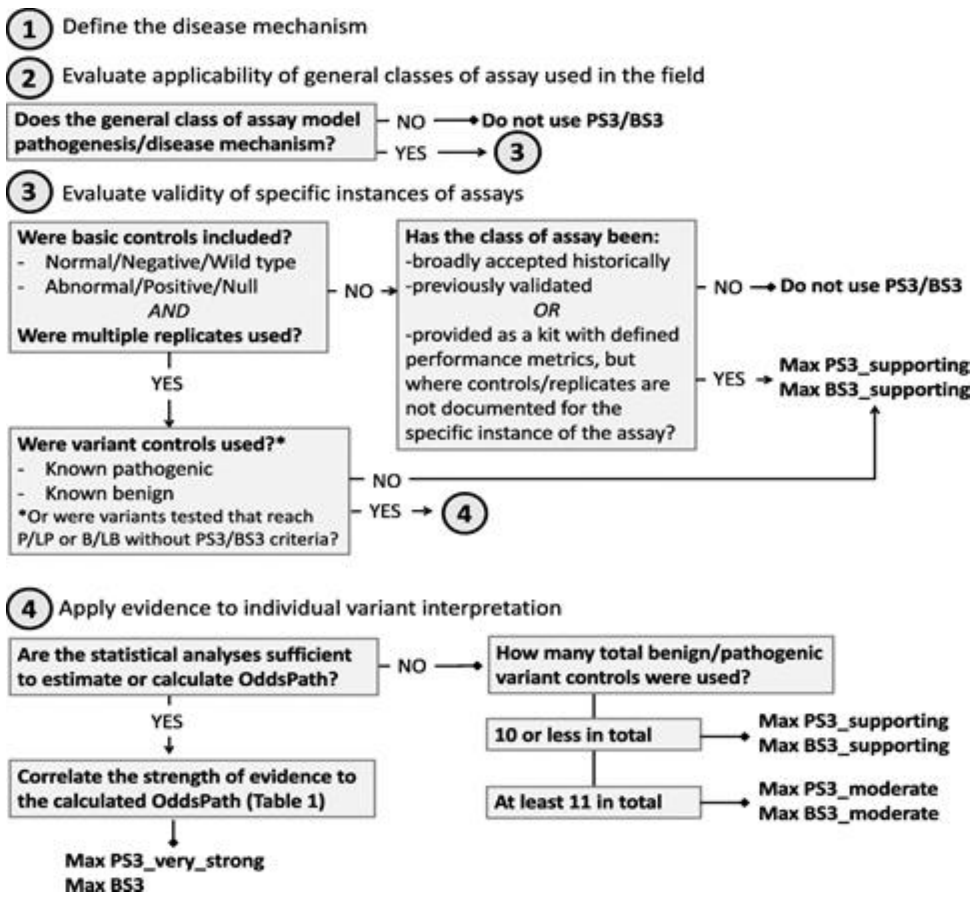

Does the assay measure the correct function? (loss of function, gain of function, dominant negative)

Is the assay predictive of pathogenicity? (how well does it differentiate between known pathogenic and benign variants)

What evidence strength can be applied to PS3/BS3?

Brnich et al. 2019 (PMID:31892348)

### DOES THE ASSAY MEASURE THE CORRECT FUNCTION?

What is the disease mechanism?

- **Loss of function**
  - Reduces or eliminates the function of the encoded protein, resulting in the partial or complete absence of the protein's activity
  - Examples include *BRCA1/2, MSH2*, countless Mendelian pathogenic variant
- **Gain of function**
  - The variant protein gains more activity than normal
  - Examples include classical oncogenic variants e.g. *FGFR3, KRAS, STAT1*
- **Dominant negative**
  - The variant protein interferes with the function of the wild-type copy
  - Examples include *TP53, FBN1, SOD1*

### SOME PROTEINS ONLY HAVE ONE DISEASE MECHANISM

**BRCA1**

**Loss of function:** loss of protein activity and defects in double strand break DNA repair by homologous recombination → hereditary breast and ovarian cancer

### SOME PROTEINS CAN HAVE VARIANTS THAT REPRESENT ALL 3 TYPES OF DISEASE MECHANISMS!

**CARD11**

**Loss of function:**
severe combined immunodeficiency (SCID)

**Gain of function:**
lymphoproliferative disease (BENTA) and high-risk of lymphoma

**Dominant negative:**
combined immune deficiency, which also variably presents with autoimmune features (CADINS)

Meitlis et al. 2020, AJHG

### IS THE ASSAY PREDICTIVE OF PATHOGENICITY?

How well does the assay separate known pathogenic and benign variants?

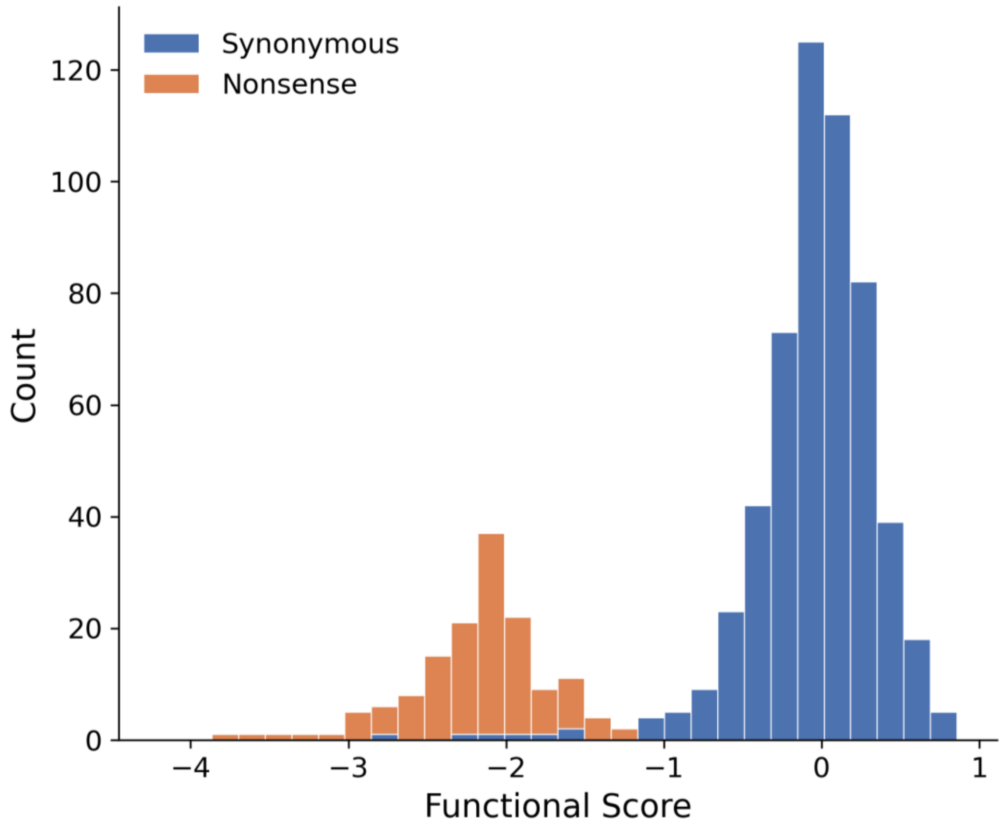

#### Analytical Performance:

Is normal separated from abnormal?

- Example: Synonymous vs nonsense
- Other options:
  - Variants with known function in other assays

### IS THE ASSAY PREDICTIVE OF PATHOGENICITY?

How well does the assay separate known pathogenic and benign variants?

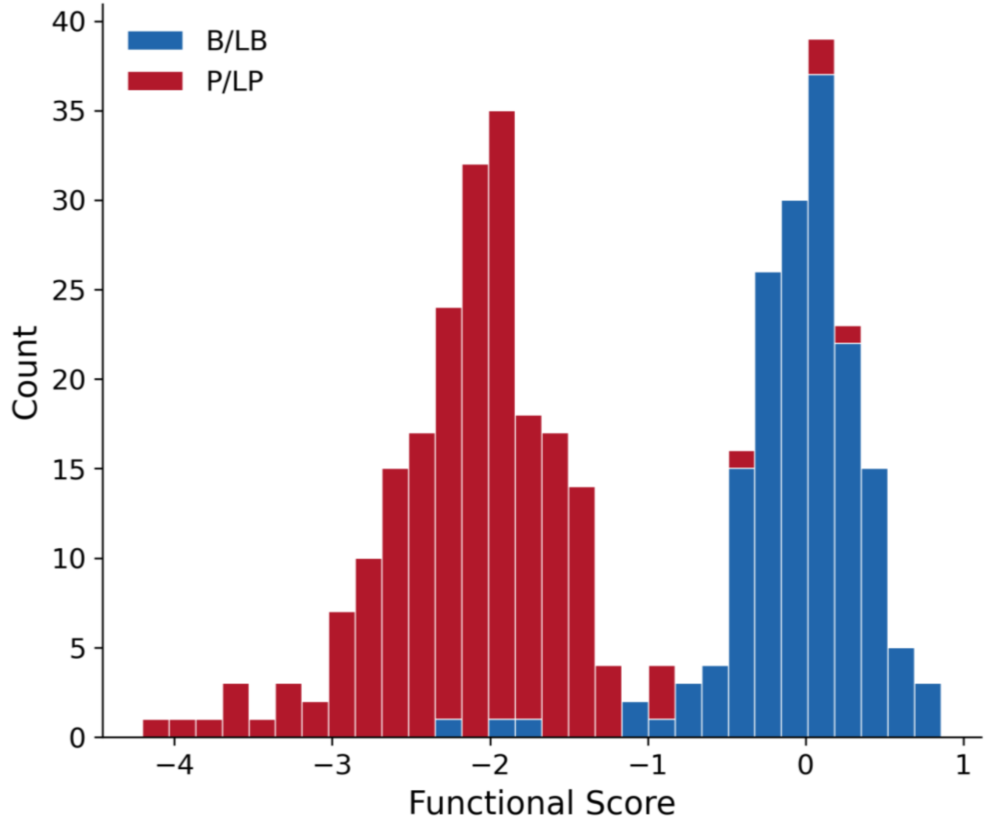

#### Clinical Performance:

Is disease causing separated from non-disease causing?

- Example: ClinVar Variants
- Other options:
  - Other databases
  - Literature
  - Clinical lab data

### WHAT EVIDENCE STRENGTH CAN BE APPLIED TO PS3/BS3?

### FUNCTIONAL DATA IS NOT FUNCTIONAL EVIDENCE!

Functional Data: Score and Functional Classification (Normal vs Abnormal)

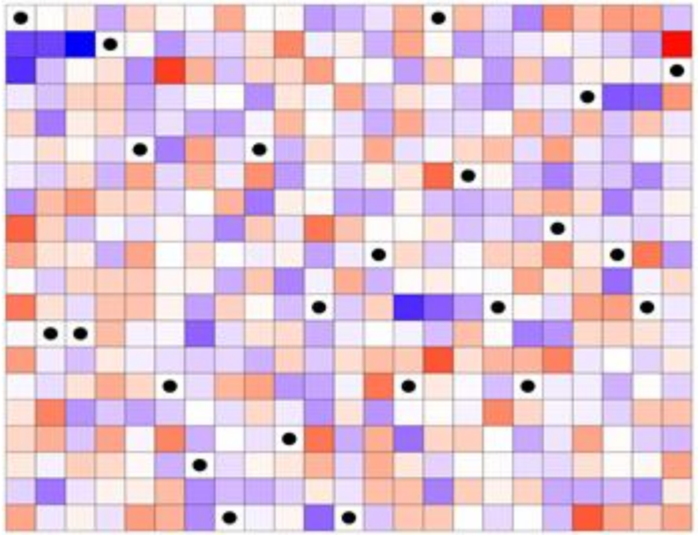

Functional Evidence: Change likelihood of disease association

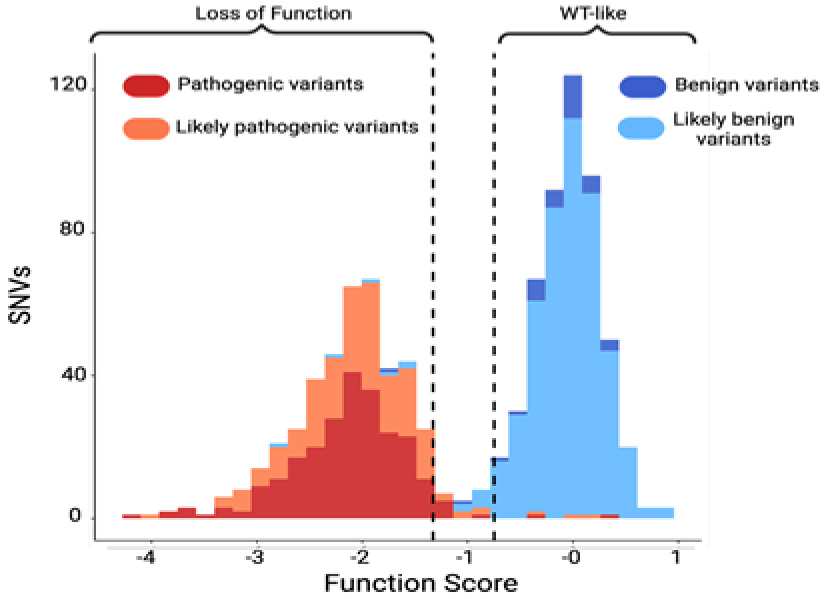

Score = -2 (functionally abnormal)
Score = 0.5 (functional normal)

Pathogenic Strong

Benign Strong

### TRANSLATING FUNCTIONAL DATA INTO EVIDENCE

Functional Scores

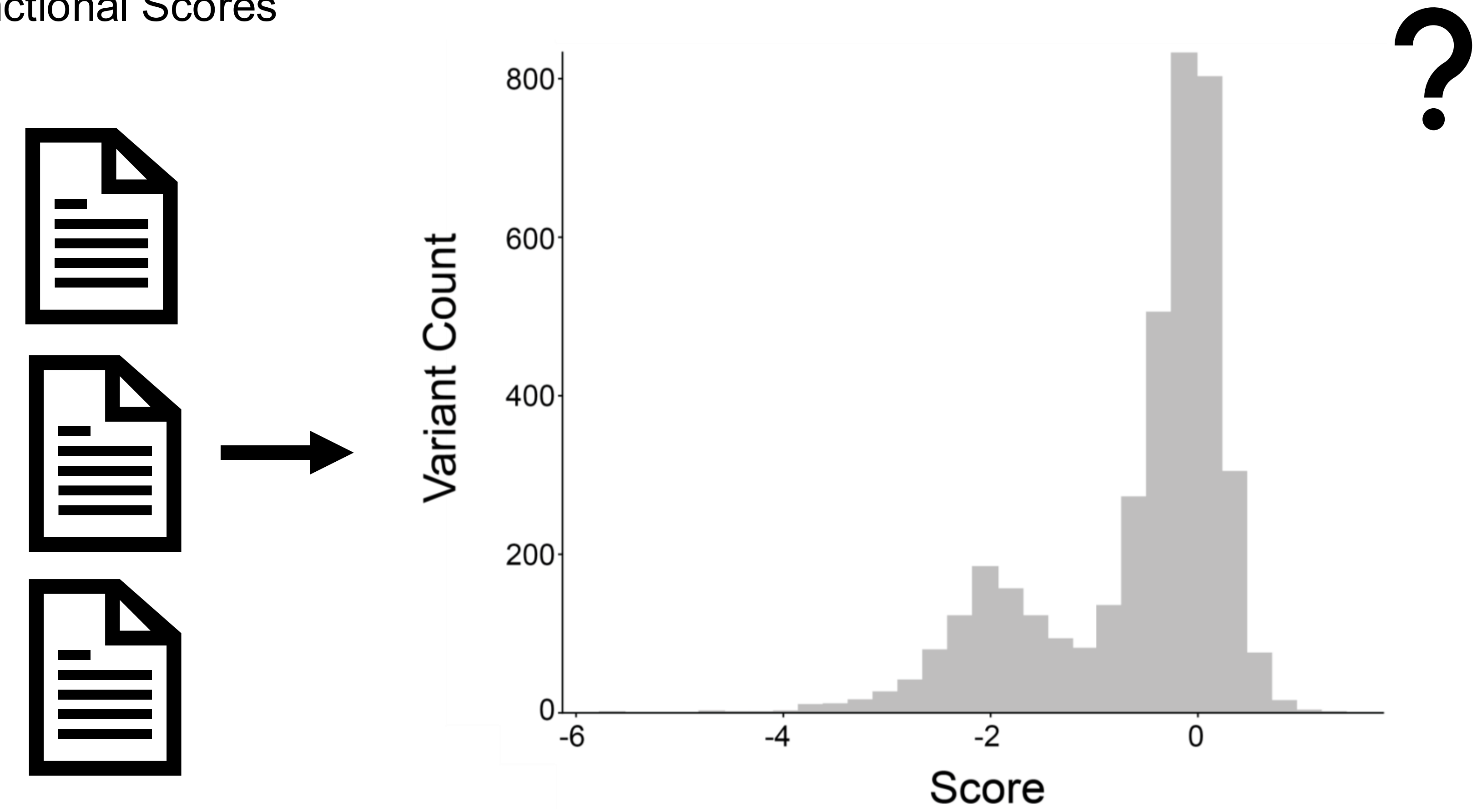

### TRANSLATING FUNCTIONAL DATA INTO EVIDENCE

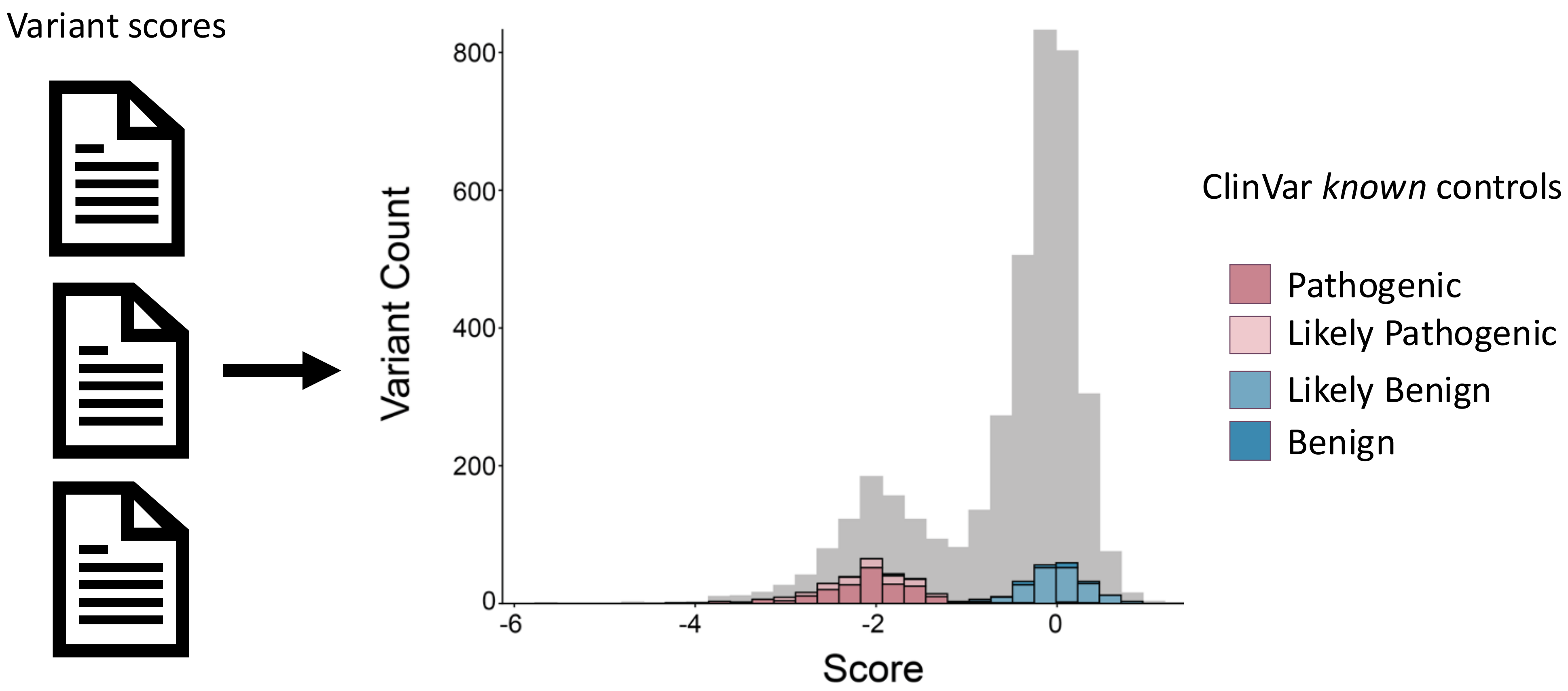

### TRANSLATING FUNCTIONAL DATA INTO EVIDENCE

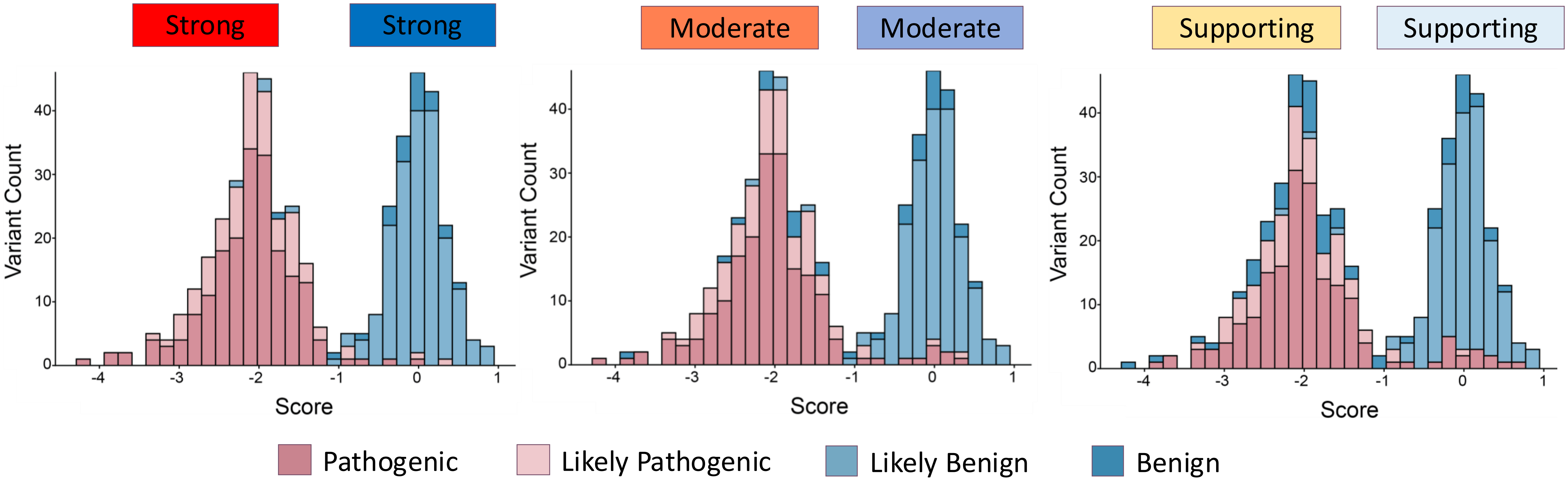

### CALCULATING EVIDENCE STRENGTH WITH LIKELIHOOD RATIOS

How much does this functional result change our confidence that a variant is pathogenic or benign?

$$\text{Prior Odds} \times \text{LR} = \text{Posterior Odds}$$

Initial Guess    New Information    Final Odds

### CALCULATING EVIDENCE STRENGTH WITH LIKELIHOOD RATIOS

Likelihood ratio: likelihood of an assay result (**normal** or **abnormal**) for variants known to be **clinically pathogenic** compared to the likelihood of the same assay result for variants known to be **clinically benign**.

$$LR+ = \frac{P(\text{functionally abnormal} \mid \text{pathogenic})}{P(\text{functionally abnormal} \mid \text{benign})}$$

$$LR- = \frac{P(\text{functionally normal} \mid \text{pathogenic})}{P(\text{functionally normal} \mid \text{benign})}$$

AKA OddsPath

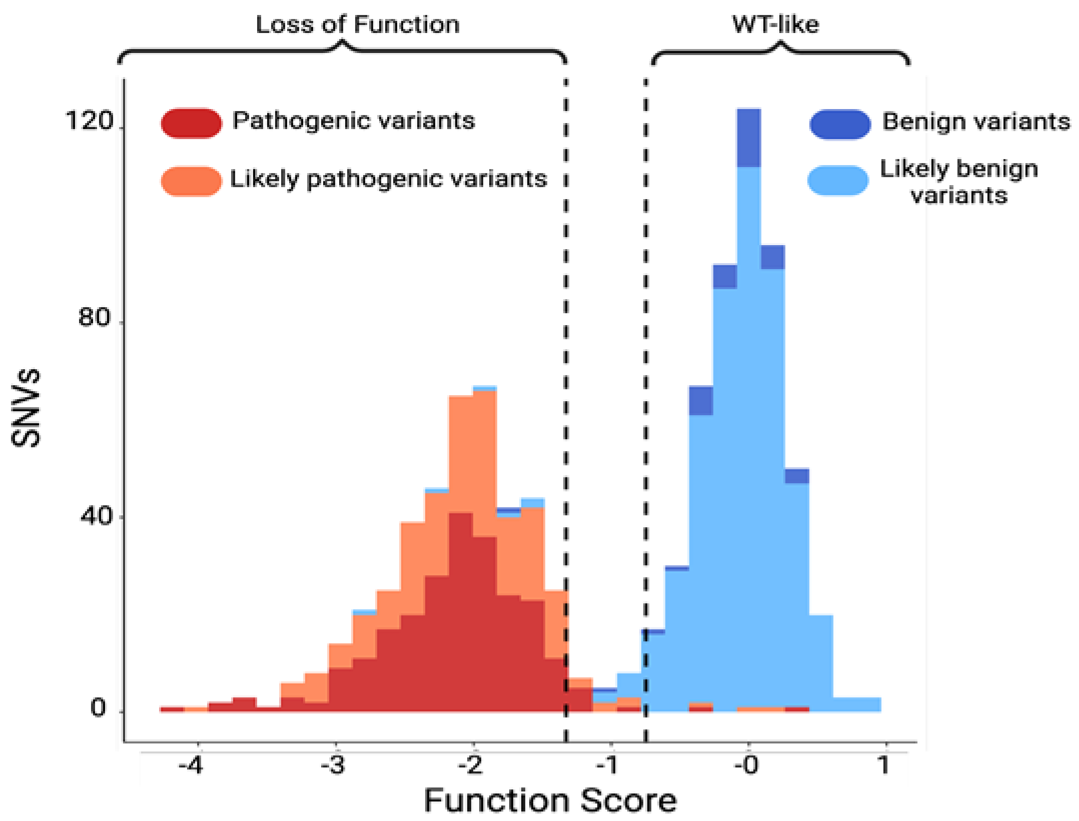

| | Benign | | Pathogenic | | |
|---|---|---|---|---|---|
| **Assay Result** | N | % | N | % | **LR** |
| **Functionally Normal** | 20 | 91 | 2 | 1 | 0.01 |
| **Indeterminate** | 1 | 5 | 5 | 3 | 0.65 |
| **Functionally Abnormal** | 1 | 5 | 162 | 96 | 19.2 |
| TOTAL = | 22 | | 169 | | |

$$LR+ = \frac{P(\text{functionally abnormal} | \textbf{pathogenic})}{P(\text{functionally abnormal} | \textbf{benign})}$$ = 96 / 5 = 19.2

$$LR- = \frac{P(\text{functionally normal} | \textbf{pathogenic})}{P(\text{functionally normal} | \textbf{benign})}$$ = 1 / 91 = 0.01

### CONVERTING LRS TO EVIDENCE STRENGTH

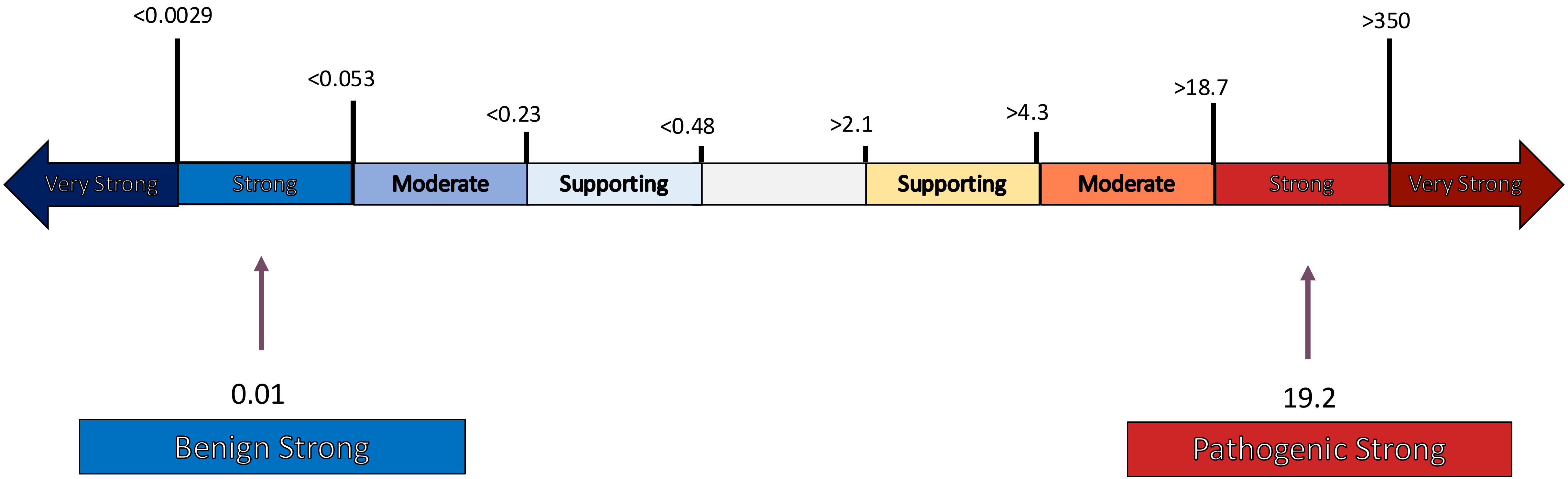

### CHANGES IN THE WORLD OF FUNCTIONAL DATA FROM 2020-2026

- Global MAVE community has generated much more functional data
- Large-scale MAVE production efforts from Sanger and NHGRI (IGVF)

- New ACMG/AMP/CAP/ClinGen v4 recommendations (set for 2026 release)
- ClinGen reorganization under SVC

- Data infrastructure readiness - minimum information standards, GA4GH variant representation, functional data models, and updates to MaveDB

### MAVE ASSAY TYPES AND TOOLS FOR CLINICAL PRACTICE

MAVEMD

### VUS ARE INCREASING EXPONENTIALLY

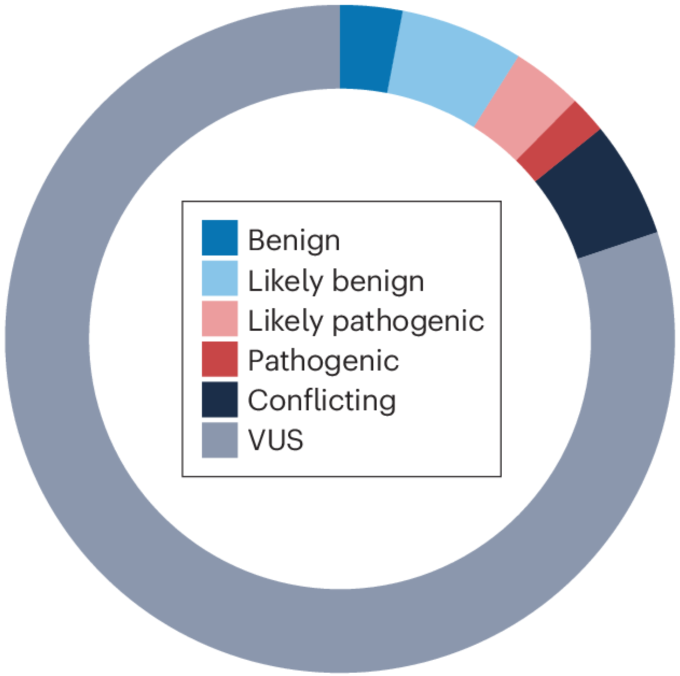

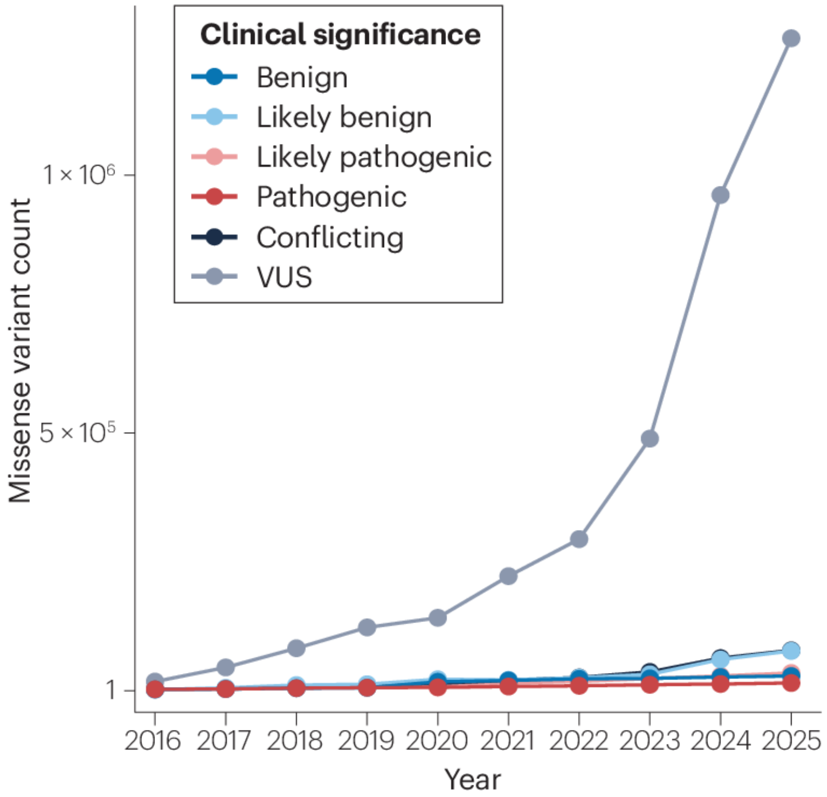

McEwen et al. 2025 (PMID: 40691352)

### VARIANTS MEASURED BY MAVES ARE ALSO GROWING EXPONENTIALLY

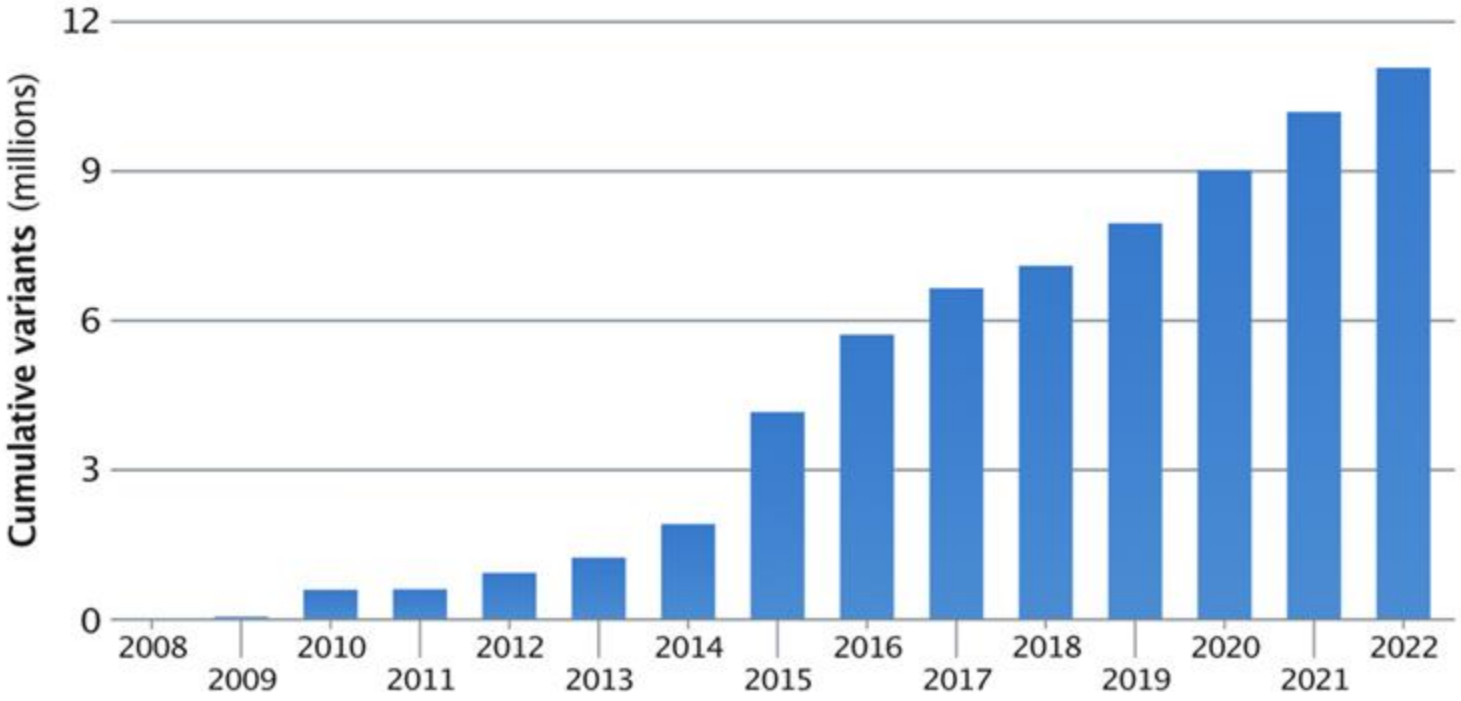

Fowler *et al.* 2023 (PMID: 37394429)

### Functional evidence derived from MAVEs drives VUS reclassification

~1500 variants in the literature have been reclassified with using MAVE evidence.

~50% were reclassified as benign, likely benign, likely pathogenic, or pathogenic.

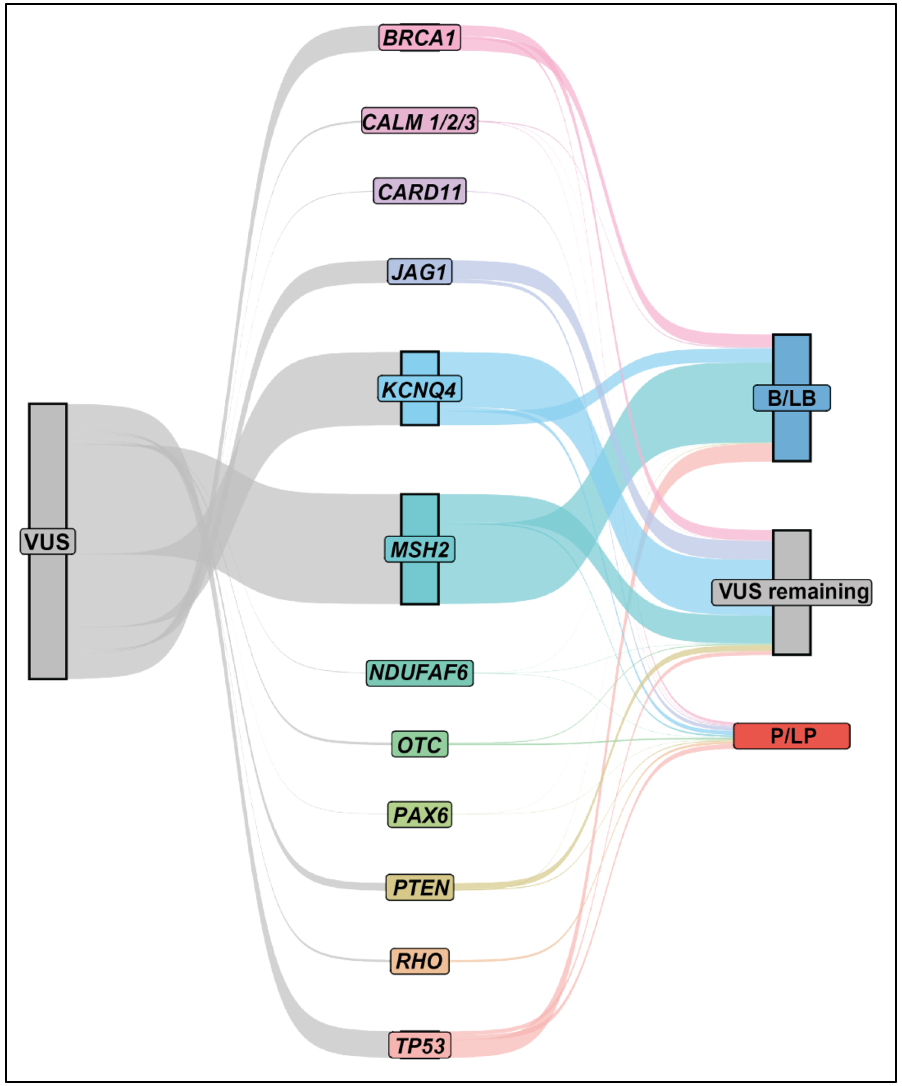

McEwen et al. 2025, *Nature Reviews Genetics*

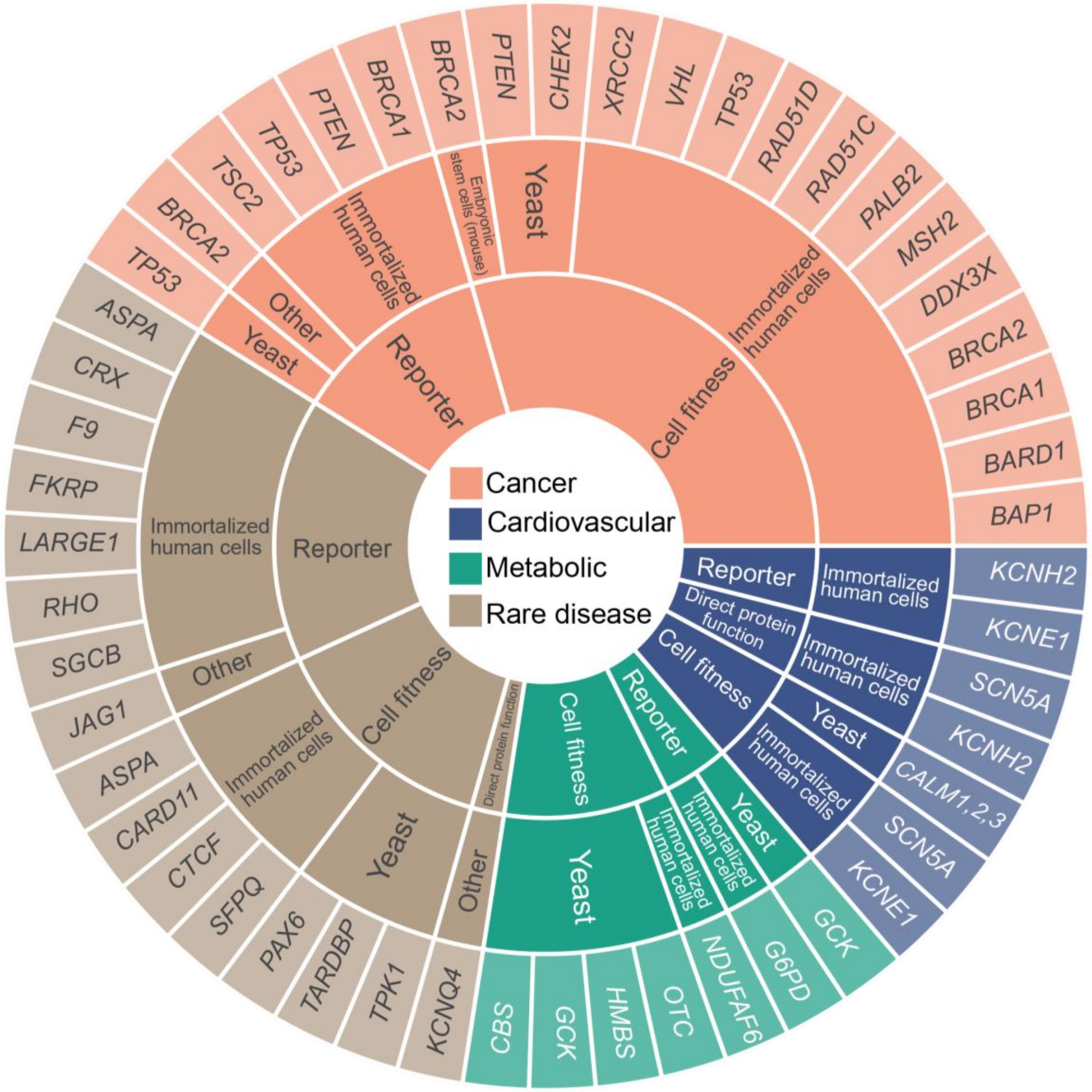

### MAVES OF MANY HUMAN DISEASE GENES

### DIVERSE MODEL SYSTEMS AND ASSAY TYPES

Tejura et al. 2026 (https://www.biorxiv.org/content/10.64898/2026.02.14.705848v1)

### **CELL FITNESS:** SATURATION GENOME EDITING (SGE)

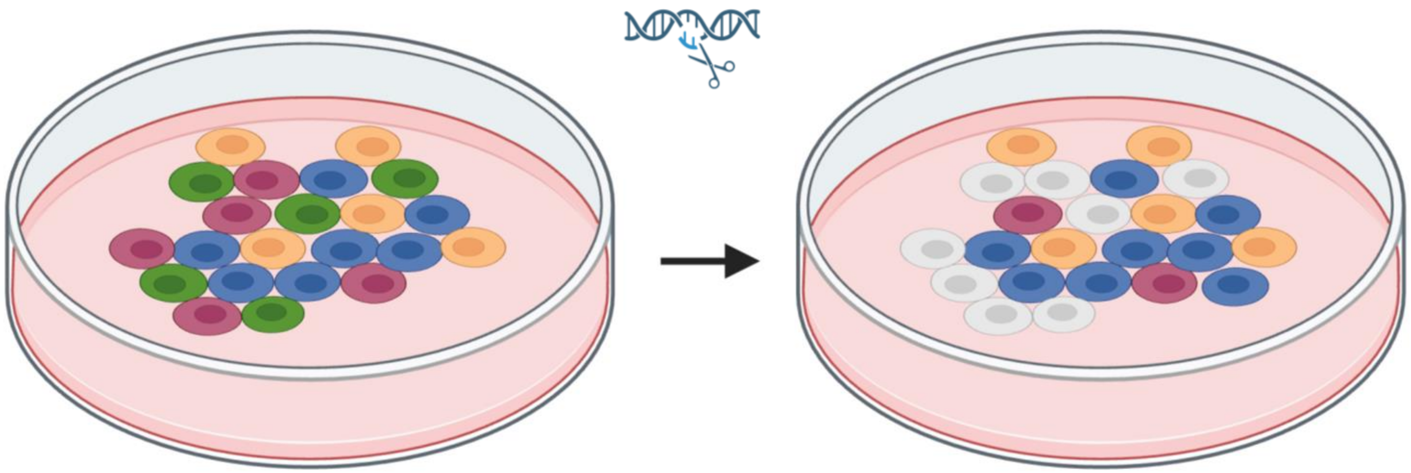

CRISPR/Cas9 used to make edits at endogenous locus. Measures the ability of a variant to allow growth or survival in cell line dependent on gene of interest.

Variant look-up table

| Variant | Score |
|---|---|
| Variant 1 | 0.9 |
| Variant 2 | 1 |
| Variant 3 | -2 |
| … | … |
| Variant n | 1.1 |

### VAMPseq: PROTEIN ABUNDANCE

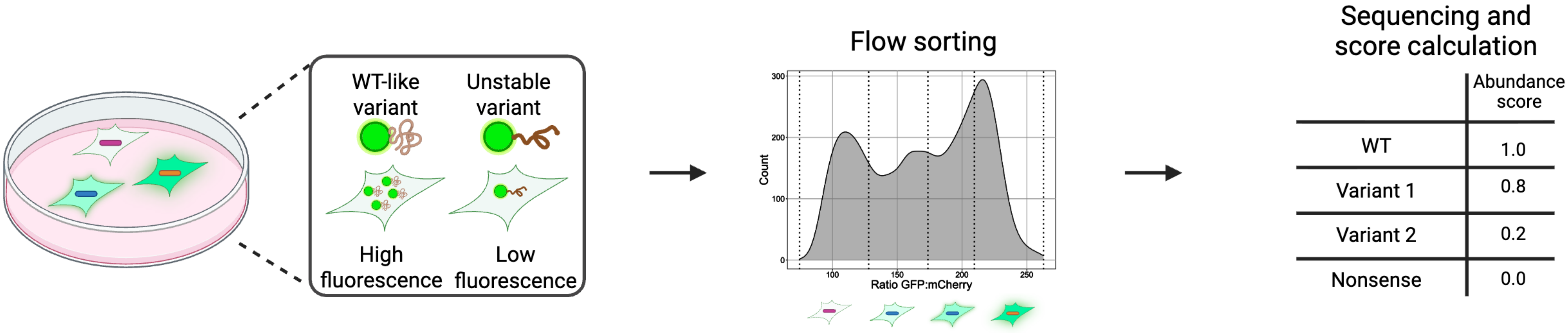

Fluorescence is a surrogate for protein abundance.

### DNA REPAIR

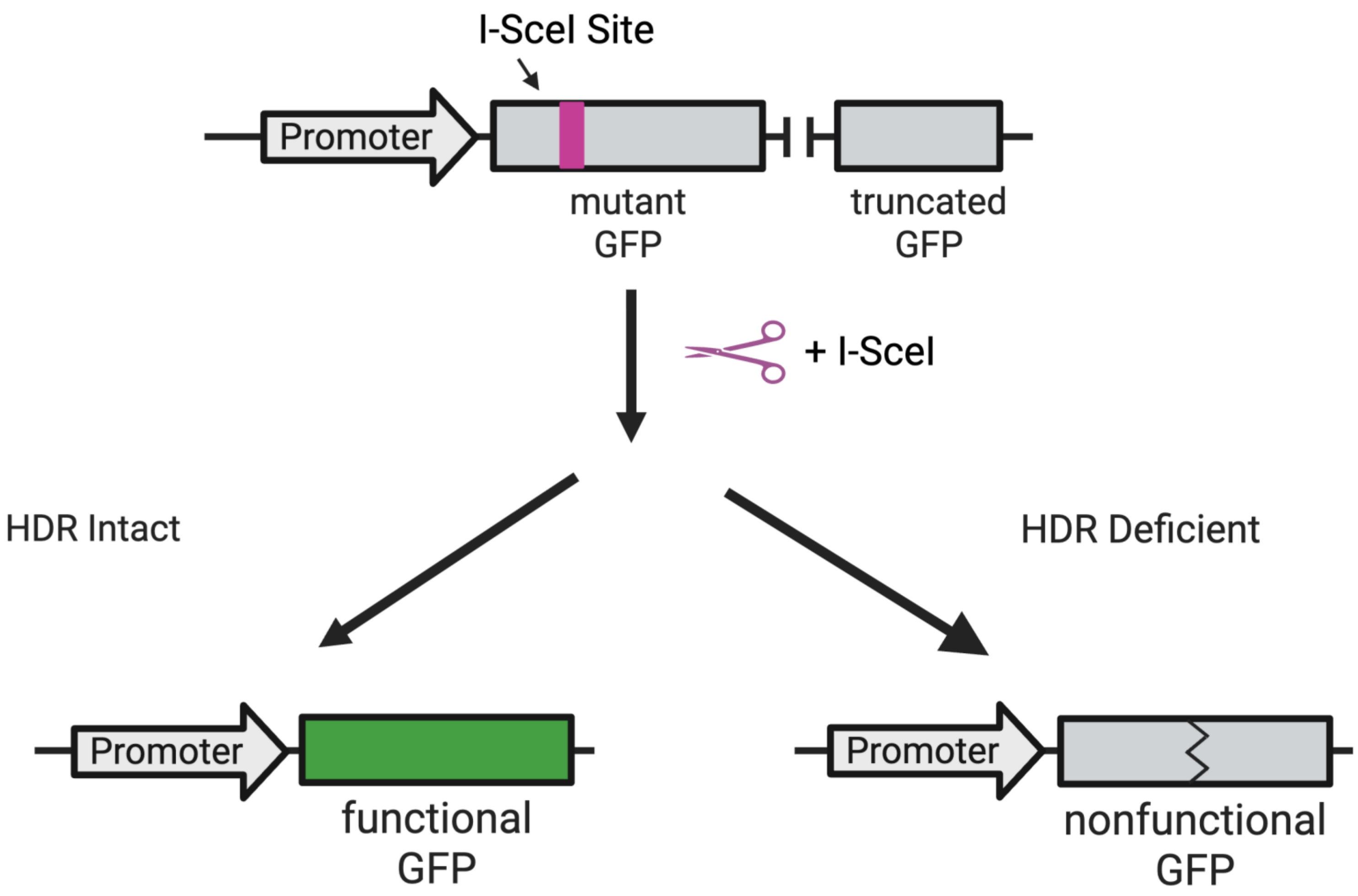

Fluorescence is a surrogate for functional HDR.

Detects one way to disrupt a protein. Provides information about mechanism.

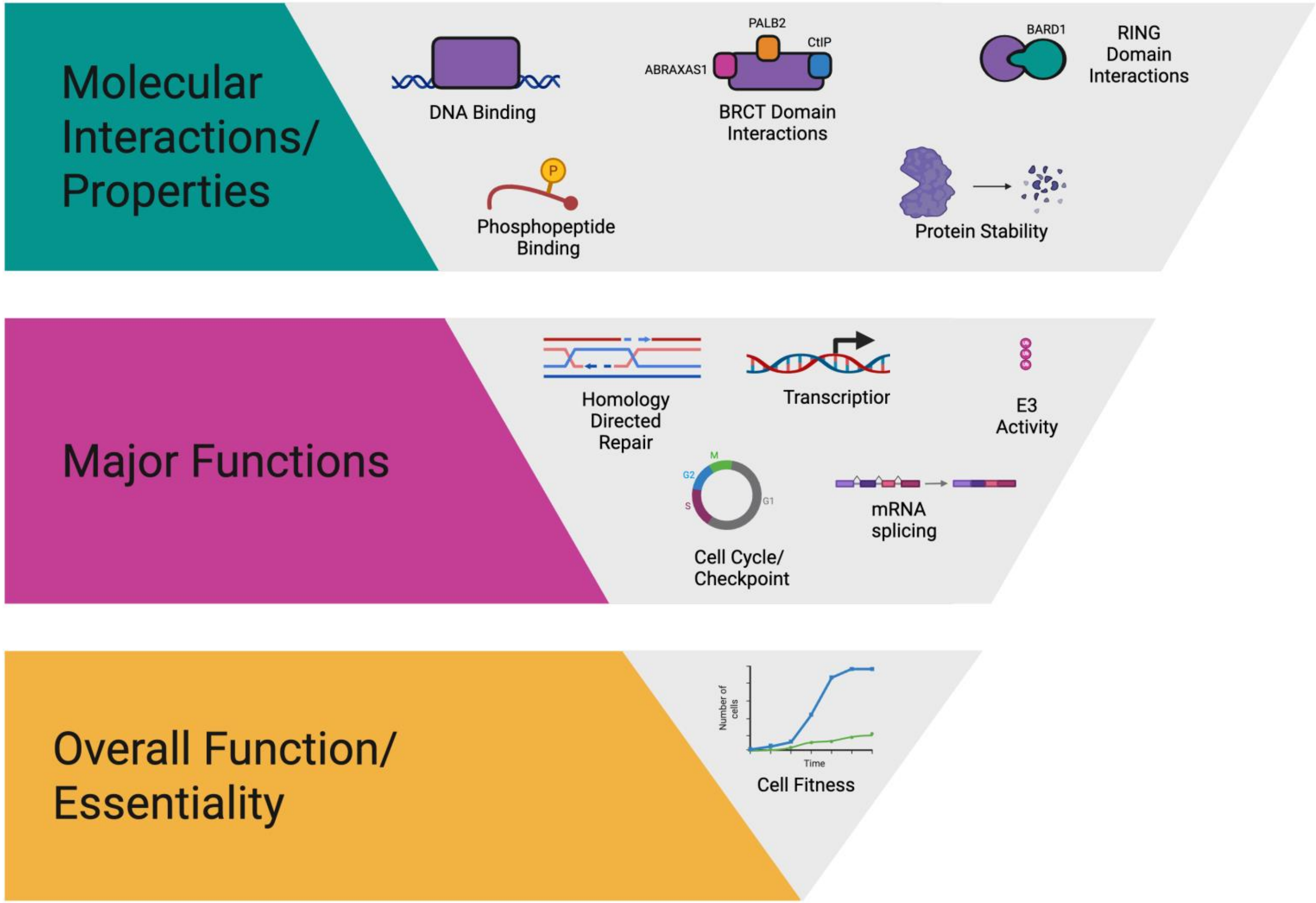

VampSeq

[illegible] Repair

SGE

Detects many ways to disrupt a protein. Cannot distinguish between mechanisms.

### WHERE CAN I QUICKLY FIND THIS INFORMATION?

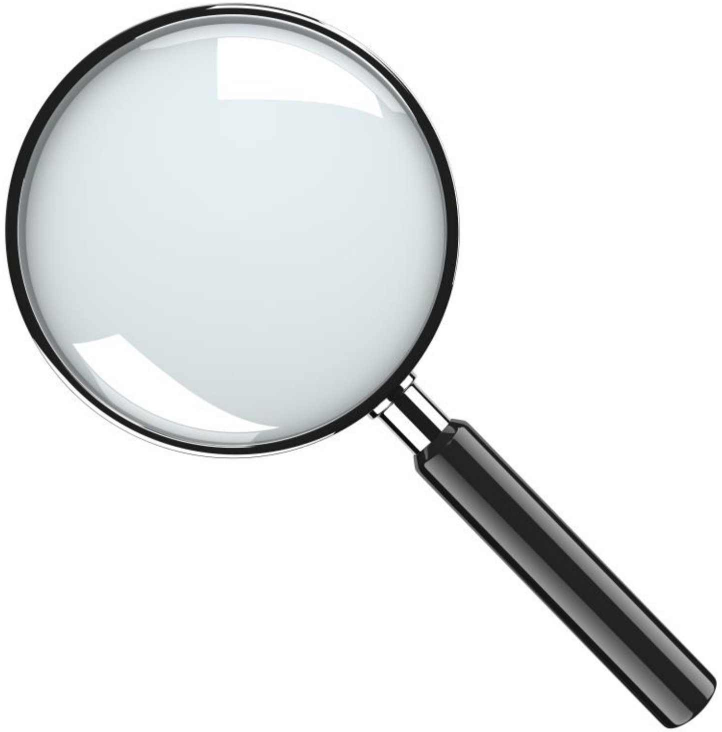

https://www.mavedb.org/

MAVEDB

Search

MaveMD Search About Support Sign in

MAVEMD

Looking for the new MaveMD clinical features? Click here.

#### About

MaveDB is a public repository for datasets from Multiplexed Assays of Variant Effect (MAVEs), such as those generated by deep mutational scanning (DMS) or massively parallel reporter assay (MPRA) experiments.

MaveDB is open-source, released under the AGPLv3 license.

MaveDB is hosted by the Fowler Lab in the Department of Genome Sciences at the University of Washington. It is supported and developed by the University of Washington, the Walter and Eliza Hall Institute of Medical Research, and the Brotman Baty Institute.

If you have questions, comments, or suggestions, check out the Help & Support page for information about how to get in touch.

#### Featured Searches

**Organisms**

- Homo sapiens
- Mus musculus
- Saccharomyces cerevisiae S288C

**Target genes**

- HSP90
- KCNQ4
- TEM-1 β-lactamase

#### Upload Your Data

Add an experiment

Add a score set

MaveMD (MAVEs for MeDicine) is an interface that integrates ClinVar and the ClinGen Allele Registry, displays clinical evidence calibrations, provides intuitive visualizations, and exports structured evidence compatible with ACMG/AMP variant classification guidelines. MaveMD currently contains 438,318 variant effect measurements mapped to the human genome from 74 MAVE datasets spanning 32 disease-associated genes.

Search MaveDB for human gene variants

HGVS | Enter a value | Search

Don't have a versioned reference sequence identifier? Click here to perform a fuzzy search instead: Fuzzy Search

Show search examples

| Gene | Score set | Publication | Calibrations w. evidence / Total |
|---|---|---|---|
| ASPA | Aspartoacylase (ASPA) cellular abundance | Grønbæk-Thygesen M et al. ASPA 2024 | 2 / 4 |
| | Aspartoacylase (ASPA) cellular toxicity | Grønbæk-Thygesen M et al. ASPA 2024 | 3 / 4 |
| BRCA1 | BRCA1 SGE Normalized Scores | Findlay et al. BRCA1 2018 | 5 / 6 |
| | Scores from growth assay of BRCA1 variants | Adamovich AI et al. BRCA1 2022 | 5 / 5 |
| | Scores from multiplexed functional assay of BRCA1 variants | Adamovich AI et al. BRCA1 2022 | 5 / 5 |
| BRCA2 | Scores from arrayed screen of BRCA2 homology directed repair function in VC-8 cells | Hu C et al. BRCA2 2024 | 4 / 5 |
| CALM1, CALM2, and CALM3 | Human Calmodulin DMS-TileSeq | Weile et al. CALM1 2017 | 1 / 2 |
| CARD11 | Scores from diploid SGE of CARD11 exons 3–5 in TMD8 cells | Meitlis et al. ENST00000396946.9 2020 | 0 / 2 |
| | Scores from diploid SGE of CARD11 exons 3–5 in TMD8 cells to select for GoF variants | Meitlis et al. ENST00000396946.9 2020 | 0 / 2 |

### MaveMD: VARIANT LEVEL SEARCHING

Variant Identifiers (HGVS string, ClinGen Allele ID, rsID)

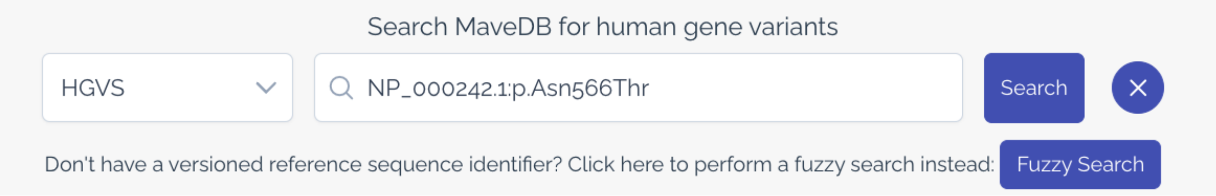

"Fuzzy Search"

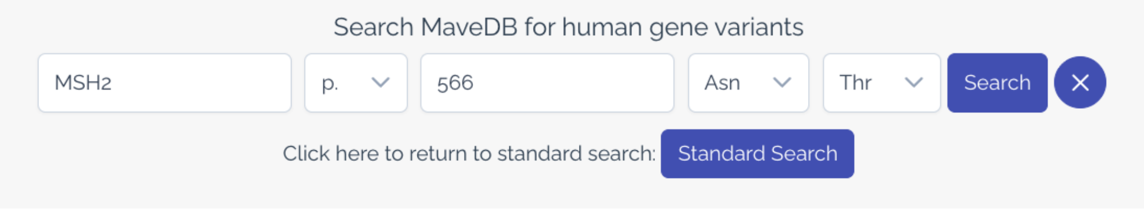

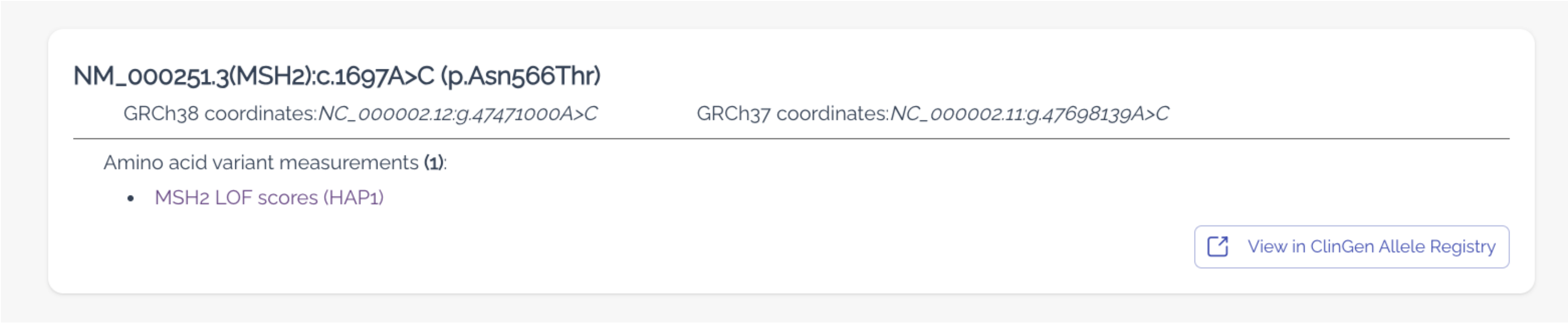

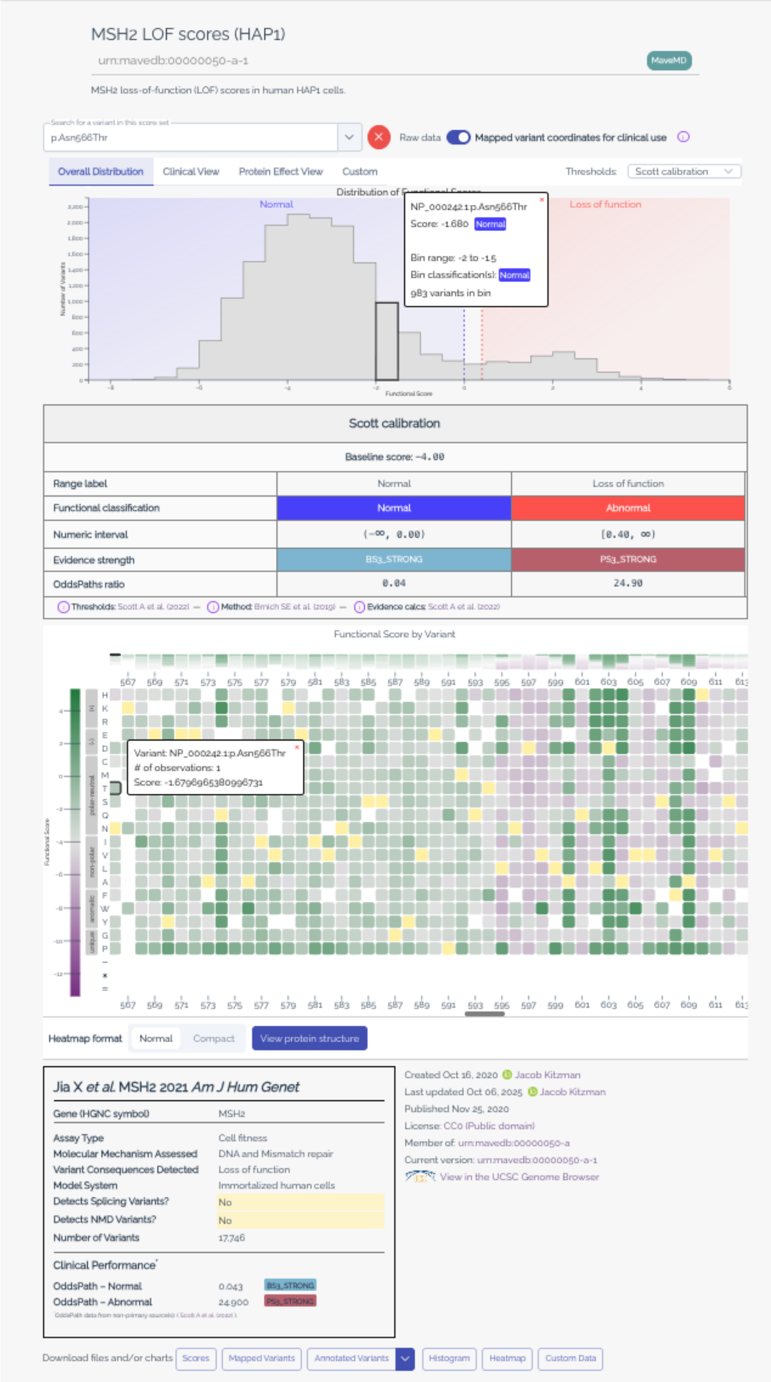

<u>Score Histogram</u>

Score Intervals

Variant score and functional classification

<u>Calibration Details</u>

OddsPath calculations

ACMG style evidence strength assignment

<u>Heat Map</u>

Functional scores by variant position
and amino acid change

<u>Assay Facts</u>

Quick reference for key assay details

### MaveMD: ANATOMY OF A VARIANT PAGE

### SCORE HISTOGRAM OPTIONS – DEFAULT VIEW

Overall Distribution | Clinical View | Protein Effect View | Custom    Thresholds: Scott calibration

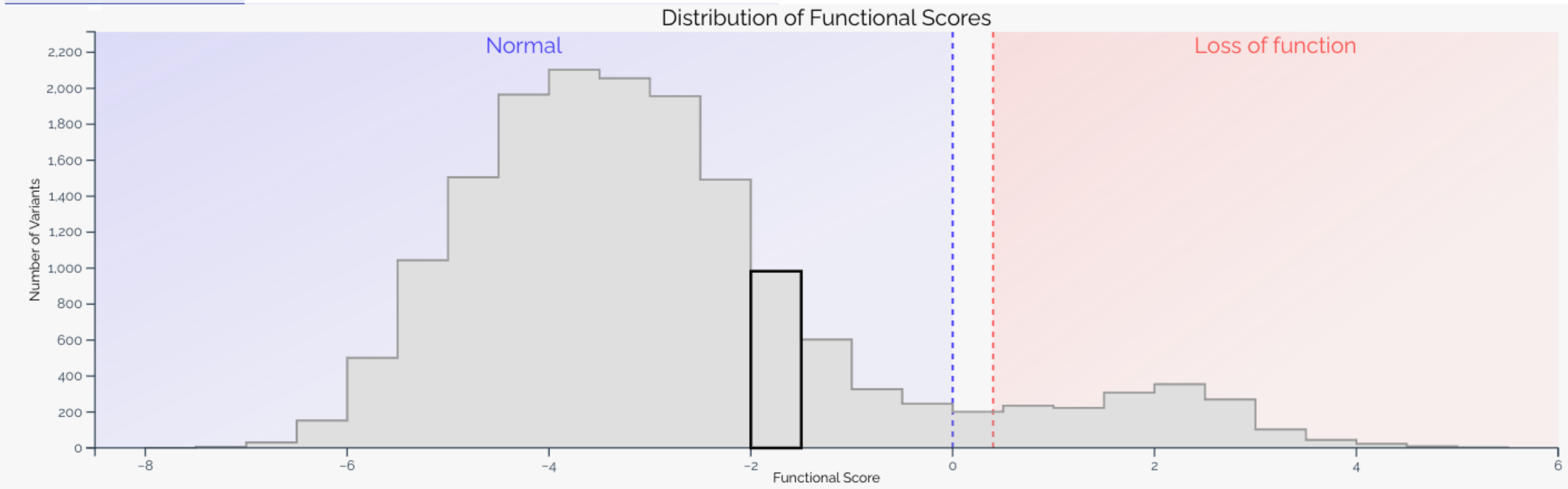

| Scott calibration | | |
|---|---|---|
| Baseline score: -4.00 | | |
| Range label | Normal | Loss of function |
| Functional classification | Normal | Abnormal |
| Numeric interval | (-∞, 0.00) | [0.40, ∞) |
| Evidence strength | BS3_STRONG | PS3_STRONG |
| OddsPaths ratio | 0.04 | 24.90 |

ⓘ Thresholds: Scott A et al. (2022) — ⓘ Method: Brnich SE et al. (2019) — ⓘ Evidence calcs: Scott A et al. (2022)

All variant distribution

Shaded by functional classification

### SCORE HISTOGRAM OPTIONS – CLINICAL VIEW

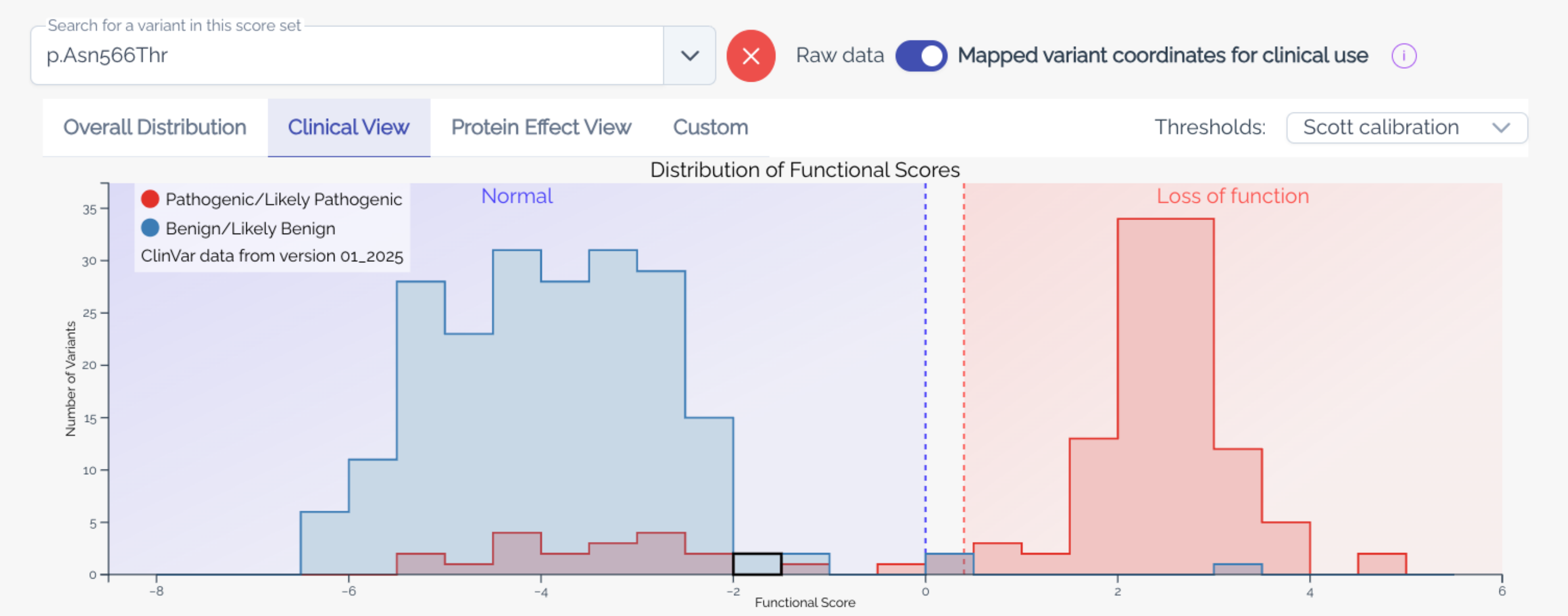

| Scott calibration | | |
|---|---|---|
| Baseline score: −4.00 | | |
| Range label | Normal | Loss of function |
| Functional classification | Normal | Abnormal |
| Numeric interval | (−∞, 0.00) | [0.40, ∞) |
| Evidence strength | BS3_STRONG | PS3_STRONG |
| OddsPaths ratio | 0.04 | 24.90 |

Thresholds: Scott A et al. (2022) — Method: Brnich SE et al. (2019) — Evidence calcs: Scott A et al. (2022)

Displays only variants with definitive (P/LP or B/LB) classifications

### THRESHOLDS AND CALIBRATIONS?

What score intervals define each functional class?
How much evidence does each functional class get?

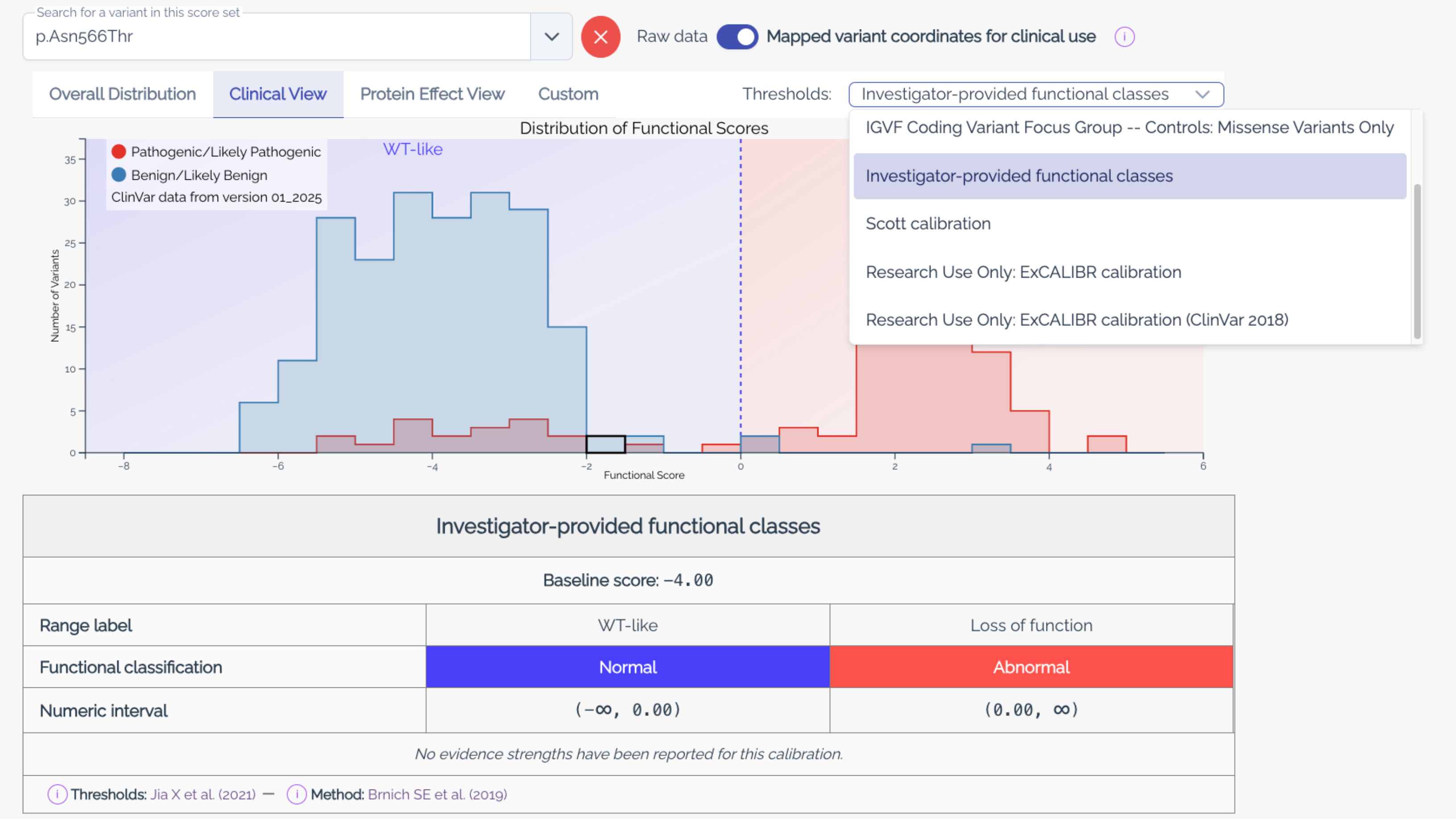

Multiple Options

- Original Publication
- Other studies

<u>Calibration details live here</u>

- Score Range and classification
- Source of thresholds
- Evidence strength assignment (if provided)

#### Original Publication

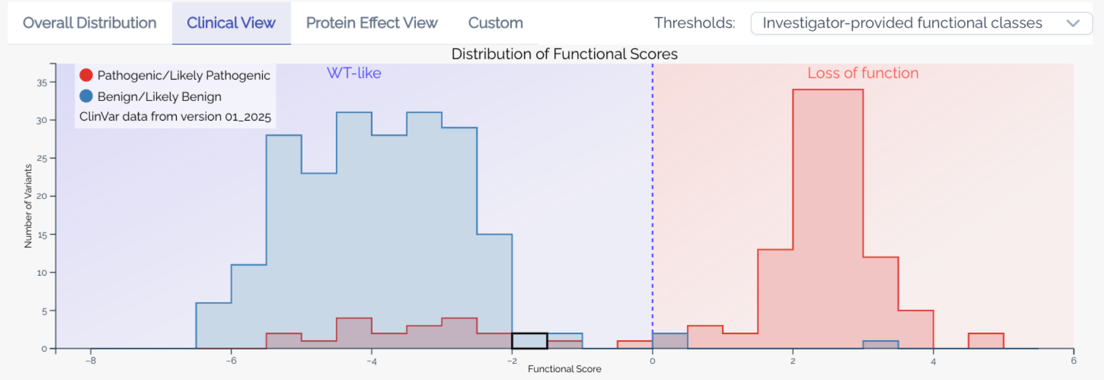

| Investigator-provided functional classes | | |
|---|---|---|
| Baseline score: −4.00 | | |
| Range label | WT-like | Loss of function |
| Functional classification | Normal | Abnormal |
| Numeric interval | (−∞, 0.00) | (0.00, ∞) |
| *No evidence strengths have been reported for this calibration.* | | |
| Thresholds: Jia X et al. (2021) — Method: Brnich SE et al. (2019) | | |

Evidence strength not calculated in original paper

#### Follow-up Publication

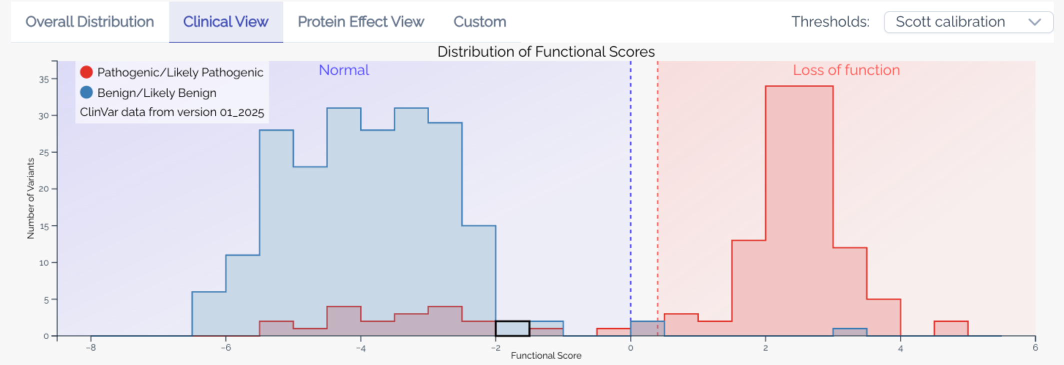

| Scott calibration | | |
|---|---|---|
| Baseline score: −4.00 | | |
| Range label | Normal | Loss of function |
| Functional classification | Normal | Abnormal |
| Numeric interval | (−∞, 0.00) | [0.40, ∞) |
| Evidence strength | BS3_STRONG | PS3_STRONG |
| OddsPaths ratio | 0.04 | 24.90 |
| Thresholds: Scott A et al. (2022) — Method: Brnich SE et al. (2019) — Evidence calcs: Scott A et al. (2022) | | |

New study used new thresholds and calculated evidence strength via Brnich OddsPath method

### ASSAY FACTS

#### Jia X *et al.* MSH2 2021 *Am J Hum Genet*

| | | |
|---|---|---|
| Gene (HGNC symbol) | MSH2 | |
| Assay Type | Cell fitness | |
| Molecular Mechanism Assessed | DNA and Mismatch repair | |
| Variant Consequences Detected | Loss of function | |
| Model System | Immortalized human cells | |
| Detects Splicing Variants? | No | |
| Detects NMD Variants? | No | |
| Number of Variants | 17,746 | |
| **Clinical Performance*** | | |
| OddsPath – Normal | 0.043 | BS3_STRONG |
| OddsPath – Abnormal | 24.900 | PS3_STRONG |

*OddsPath data from non-primary source(s): ( Scott A et al. (2022) ).

Basic metadata needed to get a quick understanding of what the assay does and what it measures

### ASSAY FACTS

#### Jia X *et al.* MSH2 2021 *Am J Hum Genet*

| | | |
|---|---|---|
| Gene (HGNC symbol) | MSH2 | |
| Assay Type | Cell fitness | |
| Molecular Mechanism Assessed | DNA and Mismatch repair | |
| Variant Consequences Detected | Loss of function | |
| Model System | Immortalized human cells | |
| Detects Splicing Variants? | No | |
| Detects NMD Variants? | No | |
| Number of Variants | 17,746 | |
| **Clinical Performance*** | | |
| OddsPath – Normal | 0.043 | BS3_STRONG |
| OddsPath – Abnormal | 24.900 | PS3_STRONG |

*OddsPath data from non-primary source(s): ( Scott A et al. (2022) ).

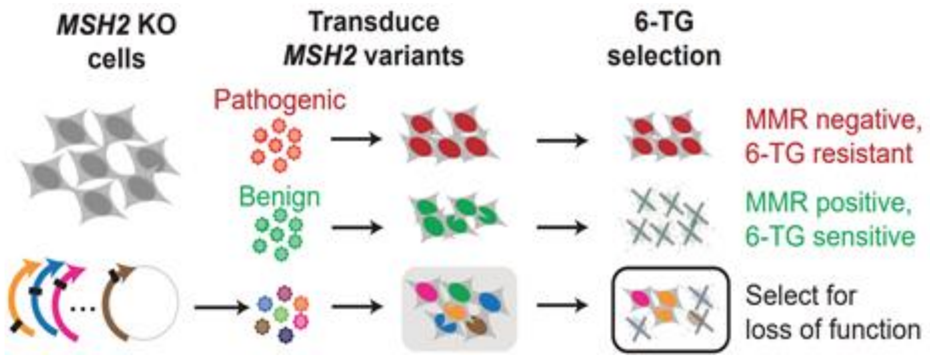

### ASSAY FACTS

##### Jia X *et al.* MSH2 2021 *Am J Hum Genet*

| | |
|---|---|
| Gene (HGNC symbol) | MSH2 |
| Assay Type | Cell fitness |
| Molecular Mechanism Assessed | DNA and Mismatch repair |
| Variant Consequences Detected | Loss of function |
| Model System | Immortalized human cells |
| Detects Splicing Variants? | No |
| Detects NMD Variants? | No |
| Number of Variants | 17,746 |

**Clinical Performance***

| | | |
|---|---|---|
| OddsPath – Normal | 0.043 | BS3_STRONG |
| OddsPath – Abnormal | 24.900 | PS3_STRONG |

*OddsPath data from non-primary source(s): ( Scott A et al. (2022) ).

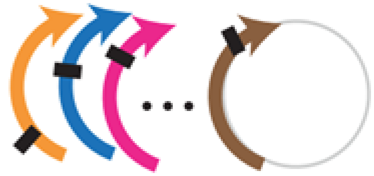

cDNA: no introns

1. can't detect splicing
2. Nonsense variants can't undergo NMD

### ASSAY FACTS

##### Jia X *et al.* MSH2 2021 *Am J Hum Genet*

| | | |
|---|---|---|
| Gene (HGNC symbol) | MSH2 | |
| Assay Type | Cell fitness | |
| Molecular Mechanism Assessed | DNA and Mismatch repair | |
| Variant Consequences Detected | Loss of function | |
| Model System | Immortalized human cells | |
| Detects Splicing Variants? | No | |
| Detects NMD Variants? | No | |
| Number of Variants | 17,746 | |
| **Clinical Performance*** | | |
| OddsPath – Normal | 0.043 | BS3_STRONG |
| OddsPath – Abnormal | 24.900 | PS3_STRONG |

*OddsPath data from non-primary source(s): ( Scott A et al. (2022) ).

Evidence Strength: defaults to author provided

### GUIDED ACTIVITY – USING FUNCTIONAL DATA FOR CLINICAL VARIANT INTERPRETATION

### CLINICAL CASE FROM THE UDN

Pediatric individual with neurodevelopmental features and ocular findings:

- Global developmental delay w/ speech/language difficulty
- Hypotonia
- Growth abnormalities (microcephaly, short stature)
- Limb abnormalities (brachydactyly, sandal gap, hallux varus)
- Head/neck abnormalities (short neck, excess nuchal fold, brachycephaly, midface hypoplasia)
- Ears abnormalities (B/L high-frequency SNHL; low-set ears; overfolded upper helices)
- Eye abnormalities (B/L microphthalmia, cataracts, iris dysgenesis, retinal detachments)

(IRB# 15HG0130)

### REPEAT EXOME SEQUENCING

- BAP1(NM_004656.4):c.687C>A (p.Asn229Lys)
  - De novo
  - Absent from population databases

### GENE-DISEASE ASSOCIATION: TUMOR PREDISPOSITION SYNDROME (TPDS)

? What do you think about a cancer predisposition gene showing up in exome for a child with NDD?

#### Gene-Disease Validity

| Gene | Disease | MOI | Expert Panel | Classification | Report & Date |
|---|---|---|---|---|---|
| BAP1 | BAP1-related tumor predisposition syndrome MONDO:0013692 | AD | Hereditary Cancer GCEP | Definitive | 03/21/2019 |

### RECENTLY DESCRIBED BAP1-ASSOCIATED NEURODEVELOPMENTAL DISORDER: KURY-ISIDOR SYNDROME (KURIS) (MIM:619762)

- Hypotonia
- Seizures
- Abnormal behavior (ASD, ADHD, hypersensitivity)
- Dysmorphic facial features
- Skeletal malformations (hands, feet, or spine)
- Growth failure
- Other organ abnormalities (eyes, heart, GU)

9 *de novo* het **missense** variants (11 individuals)

Kury et al 2022 (PMID: 35051358)

### VARIANT CONSEQUENCE VS PHENOTYPE

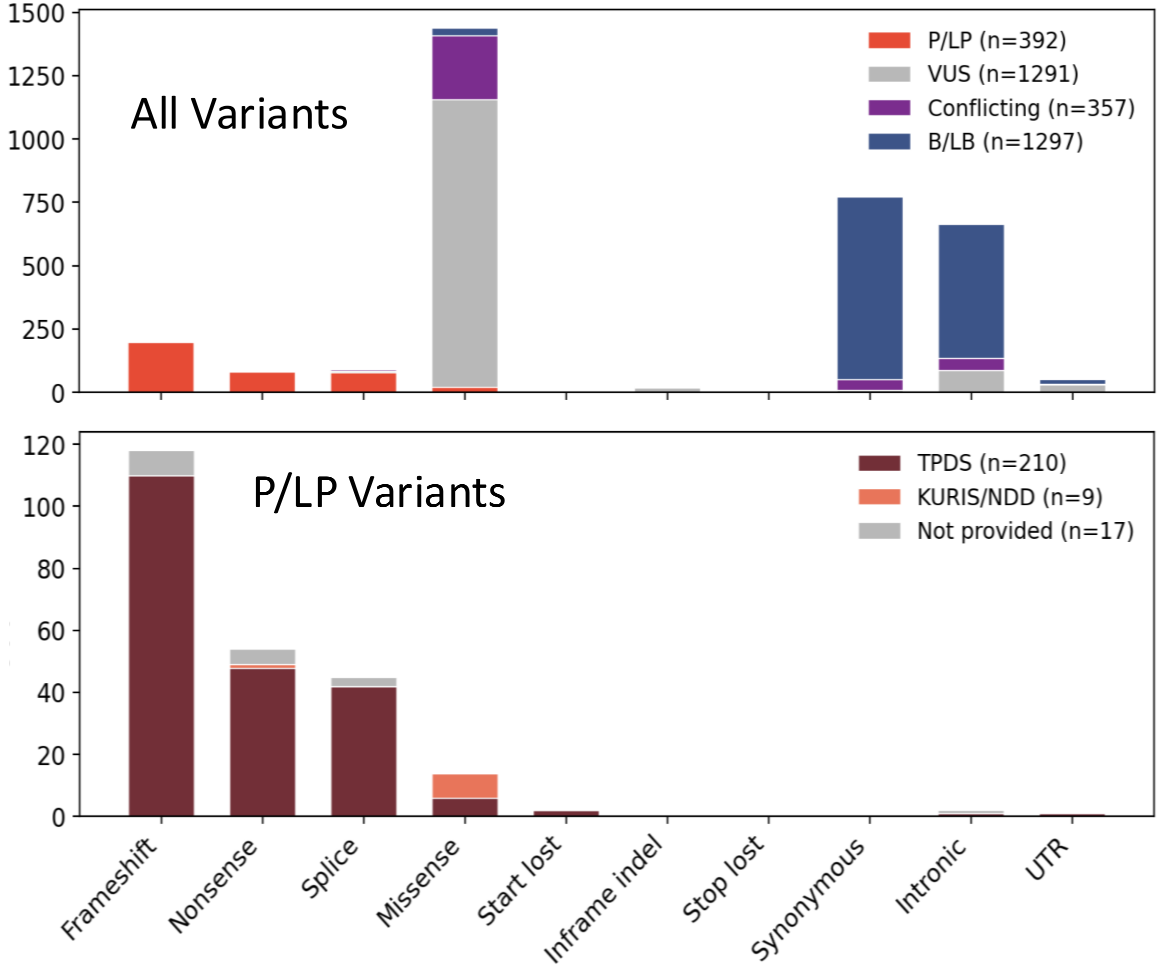

| Variant Source | Molecular Consequence |
|---|---|
| P/LP All ClinVar | Truncating |
| TPDS | Truncating |
| KURIS/NDD | Missense |

What kind of L/P variants dominate the ClinVar dataset?

Truncating cancer variants

ClinVar Feb 2026

### BAP1 SATURATION GENOME EDITING

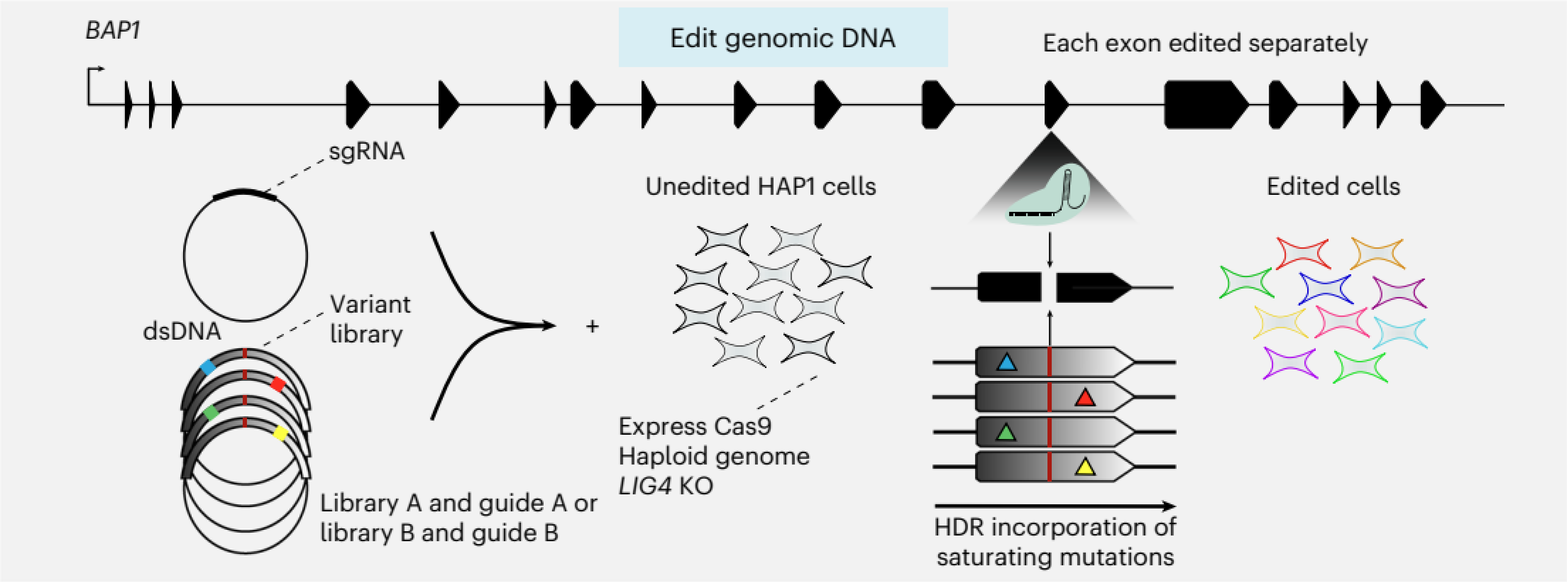

Waters et al 2024 (PMID: **38969833)**

### BAP1 SATURATION GENOME EDITING

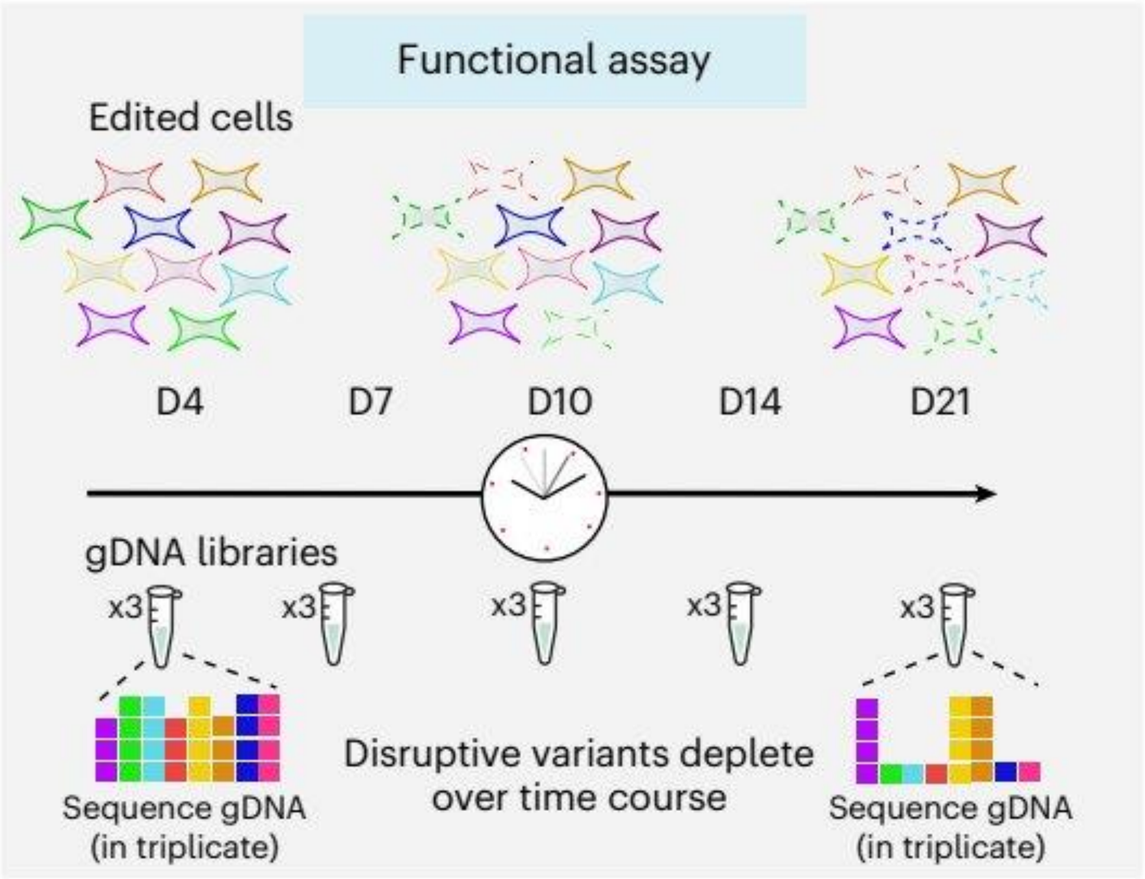

- Assayed ~**18,000** distinct variants
- Deleterious variants are depleted

Waters et al 2024 (PMID: **38969833)**

Normal Function = 0

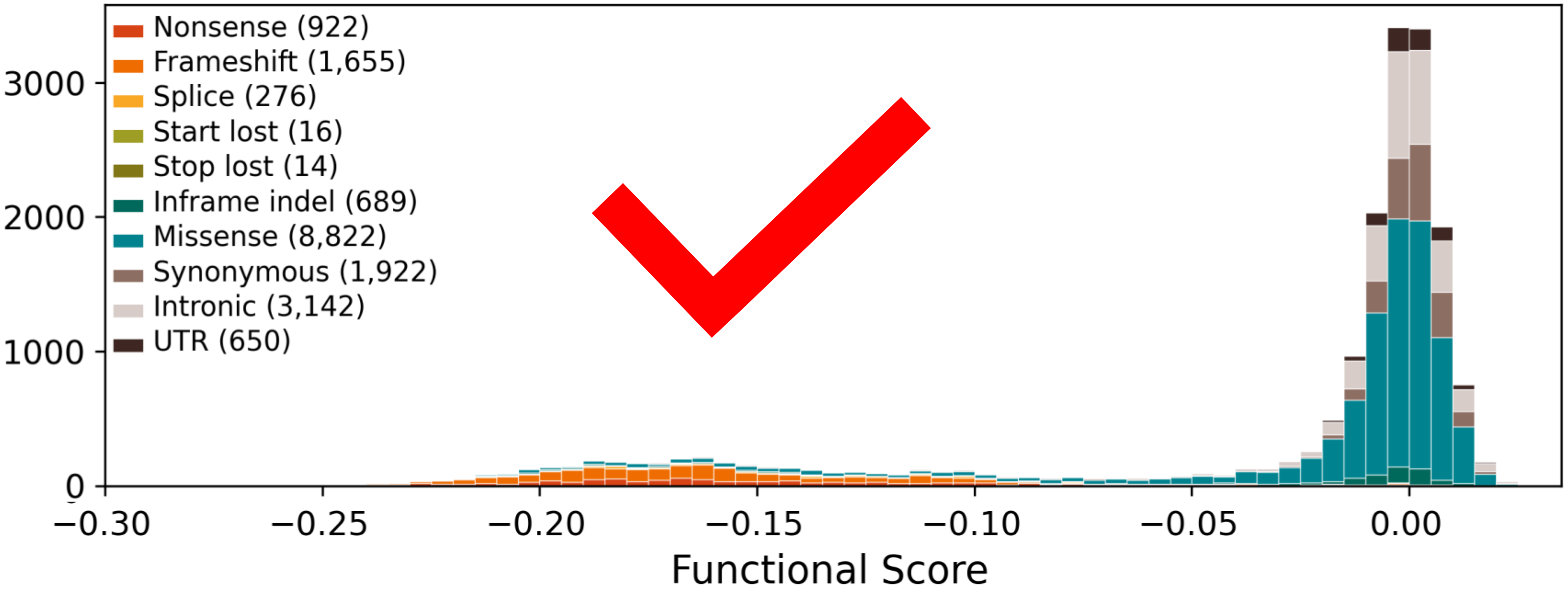

~~Analytical Performance:~~

Separation between truncating and synonymous?

Waters et al 2024 (PMID: **38969833)**

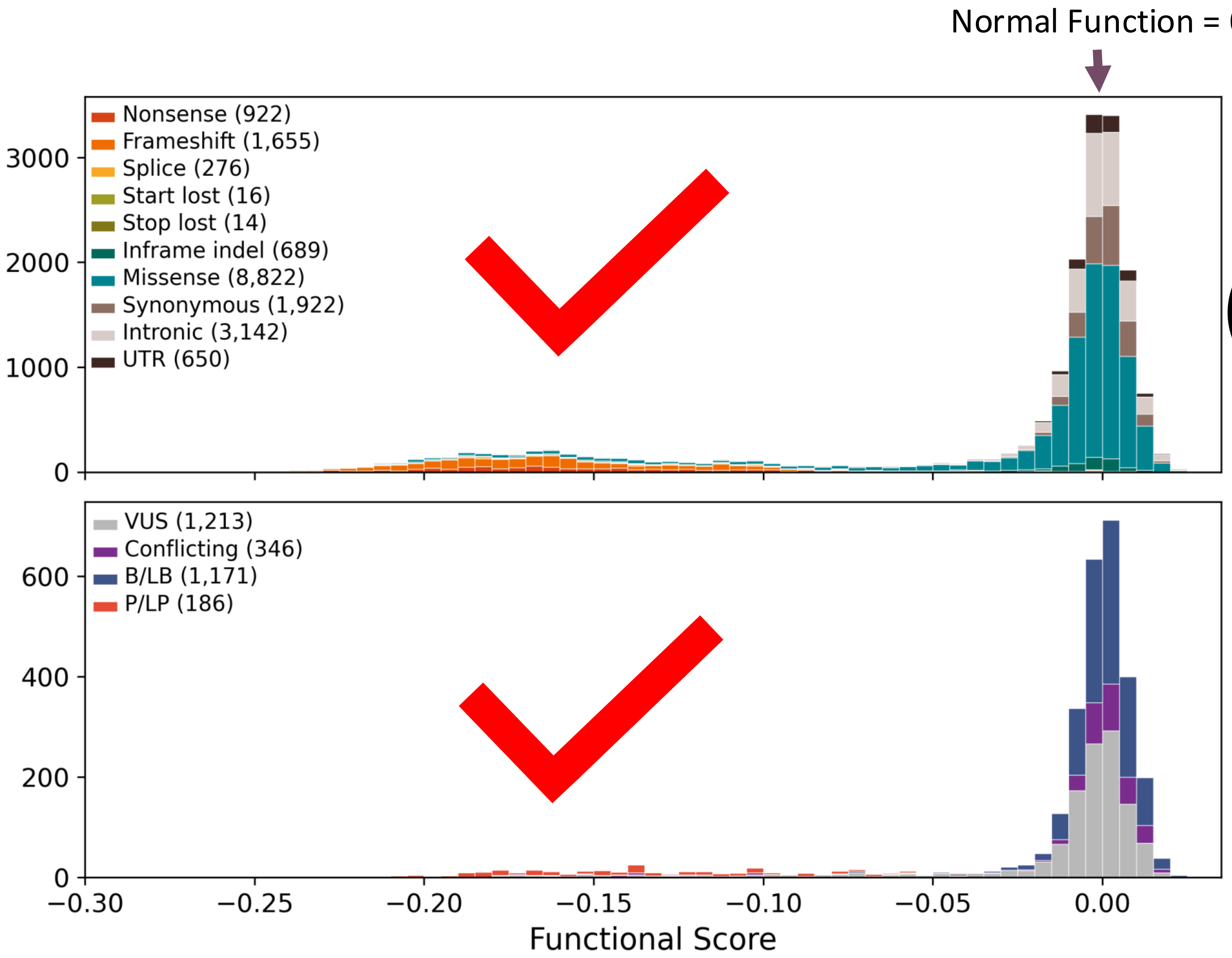

Analytical Performance:

Separation between truncating and synonymous?

Clinical Performance:

Separation between benign and pathogenic?

Waters et al 2024 (PMID: **38969833)**

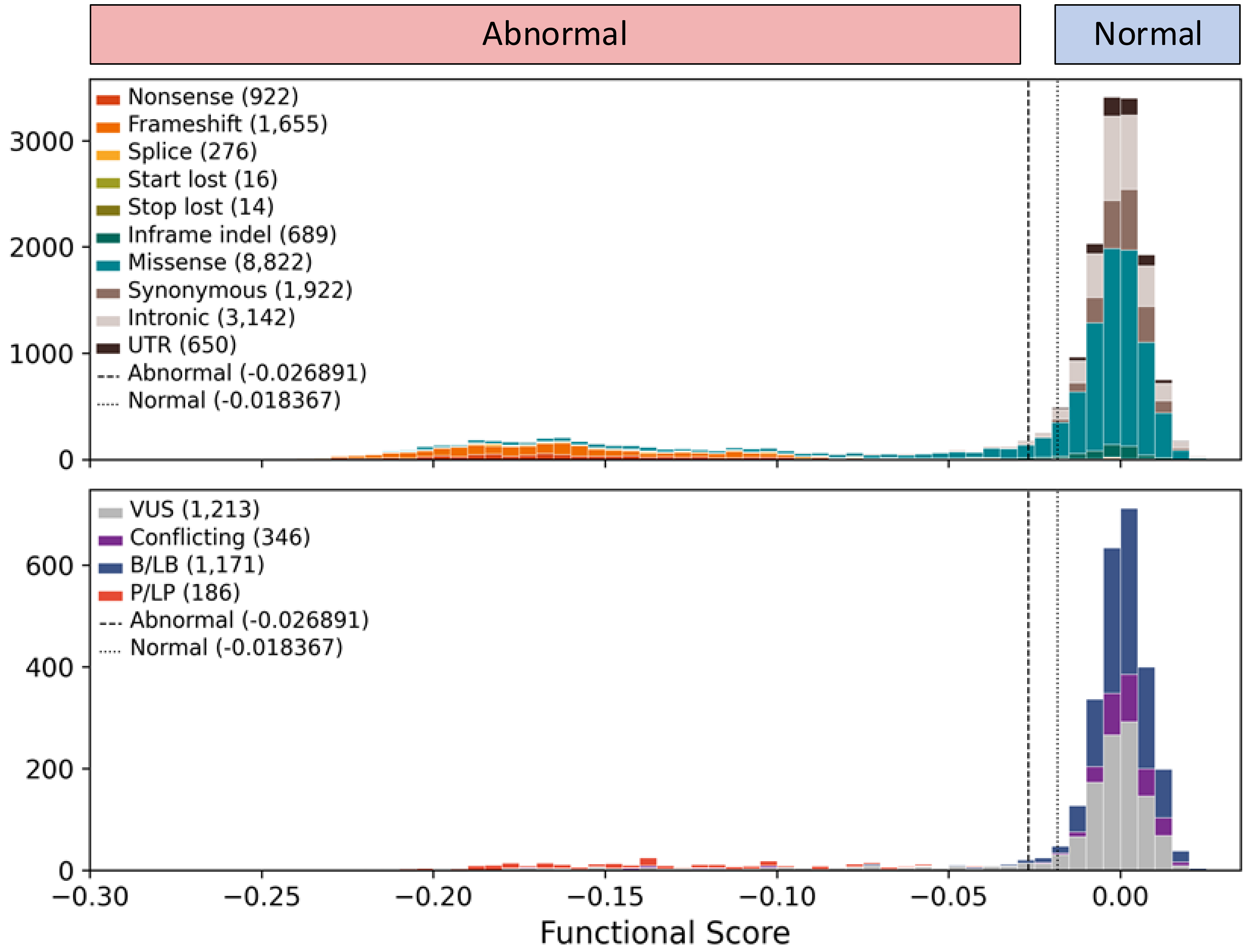

### Separating normal from abnormal

Many methods quantitative methods

Example:
Gaussian Mixture Model

Waters et al 2024 (PMID: **38969833)**

### Waters AJ *et al.* BAP1 2024 *Nat Genet*

| | |
|---|---|
| Gene (HGNC symbol) | BAP1 |
| Assay Type | Cell fitness |
| Molecular Mechanism Assessed | Not specified |
| Variant Consequences Detected | Loss of function |
| Model System | Immortalized human cells |
| Detects Splicing Variants? | Yes |
| Detects NMD Variants? | Yes |
| Number of Variants | 18,108 |

#### Clinical Performance

| | | |
|---|---|---|
| OddsPath – Normal | 0.002 | BS3_STRONG |
| OddsPath – Abnormal | 27.600 | PS3_STRONG |

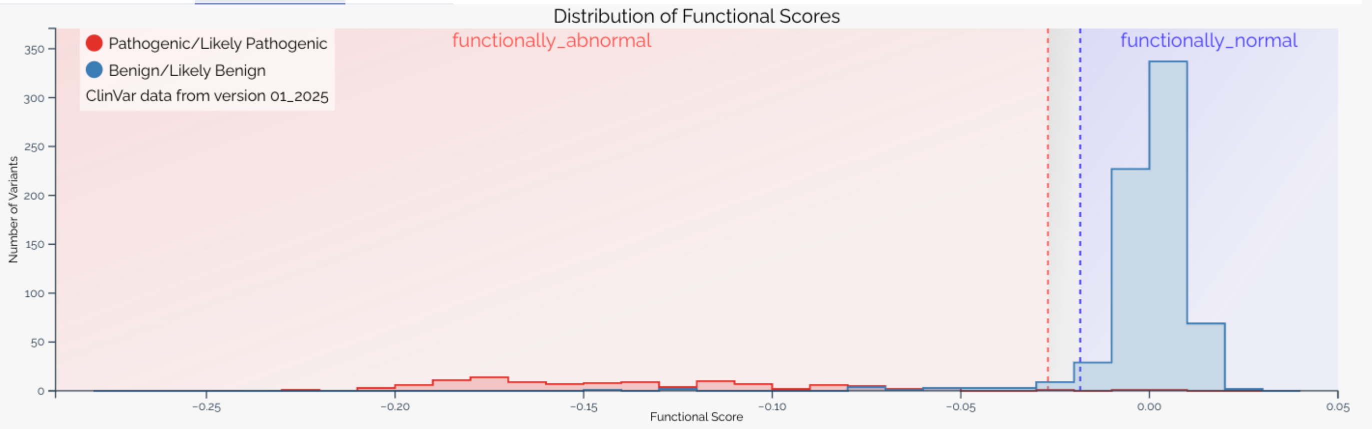

Research Use Only: Gaussian mixture model with ranges

Baseline score: 0.00

| Range label | functionally_abnormal | indeterminate | functionally_normal |
|---|---|---|---|
| Functional classification | Abnormal | Not Specified | Normal |
| Numeric interval | (−∞, −0.03) | [−0.03, −0.02] | (−0.02, ∞) |
| Evidence strength | PS3_STRONG | — | BS3_STRONG |
| OddsPaths ratio | 42.14 | — | 0.01 |

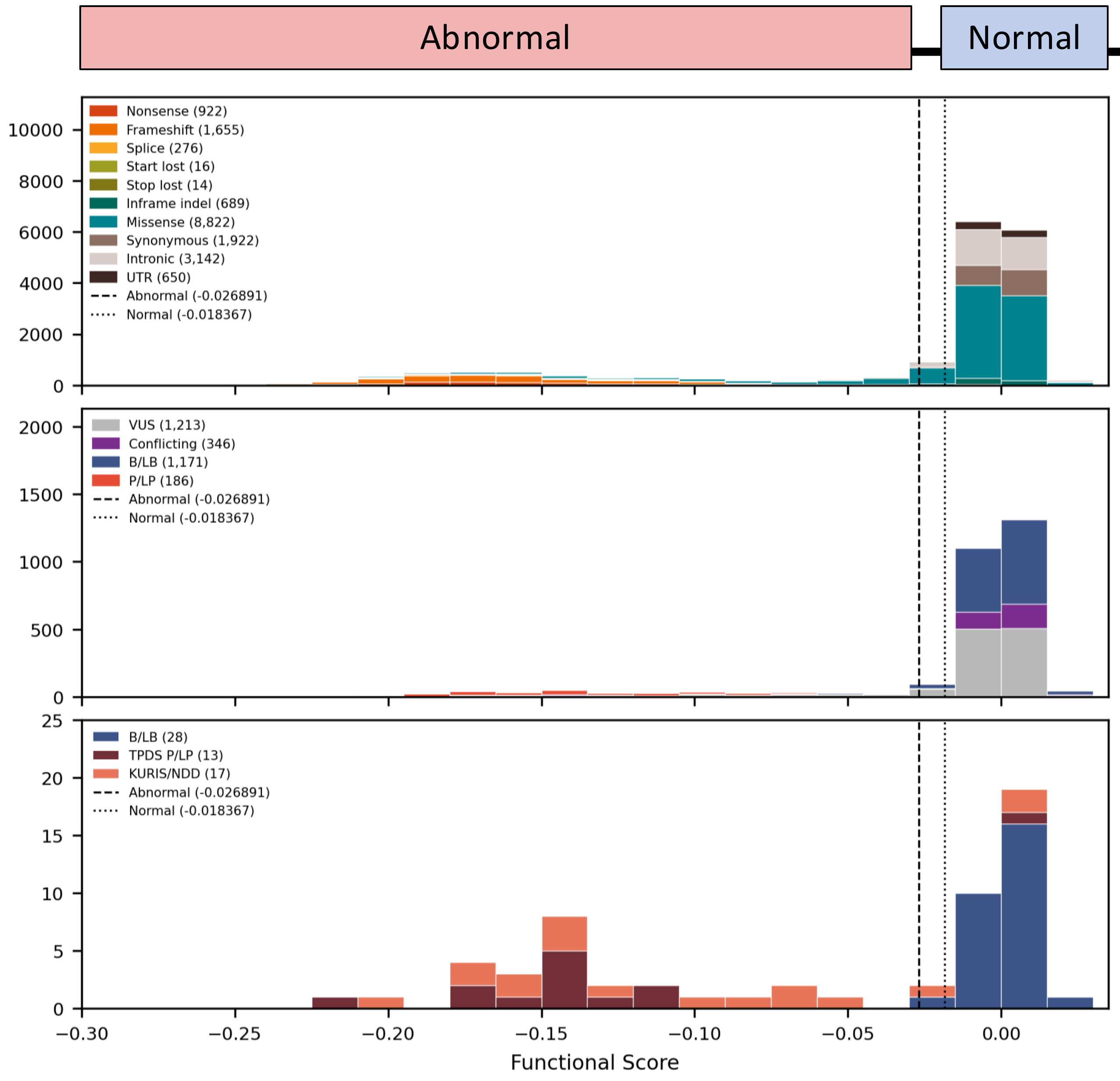

### BAP1 IS ASSOCIATED WITH TWO DISTINCT ENTITIES

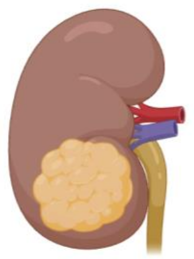

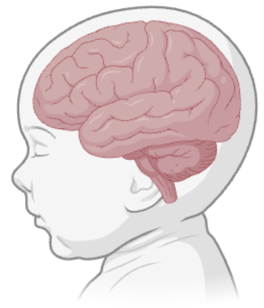

Cancer vs Neurodevelopmental Disorder

Assay thresholds primarily driven by truncating variants (cancer)

Does the SGE assay detect both phenotypes?

### CALCULATING EVIDENCE STRENGTH

How to calculate, re-calculate or calibrate assay evidence yourself

Why?

- Maybe the paper does not provide evidence strength
- You want to validate the evidence for a specific phenotype
- You prefer to use a more ‘rigorous’, more recent or just a different control/reference set to assess evidence strength

How

Tavtigian et Al. 2018 PMID: 29300386

Brnich et Al. 2019 PMID: 31892348

### LIKELIHOOD RATIO (LR)

- LR estimates the association (or strength of evidence) of a particular test result with a clinical outcome (pathogenicity)

- Can use LR to adjust our assessment of pathogenicity

$$LR+ = \frac{P(\text{functionally abnormal} \mid \text{pathogenic})}{P(\text{functionally abnormal} \mid \text{benign})}$$

$$LR- = \frac{P(\text{functionally normal} \mid \text{pathogenic})}{P(\text{functionally normal} \mid \text{benign})}$$

To read about LR, see Deeks et Al. 2004 PMID: 15258077

### BAP1 FUNCTIONAL ASSAY RESULTS

Assay results presented as 'functional categories'

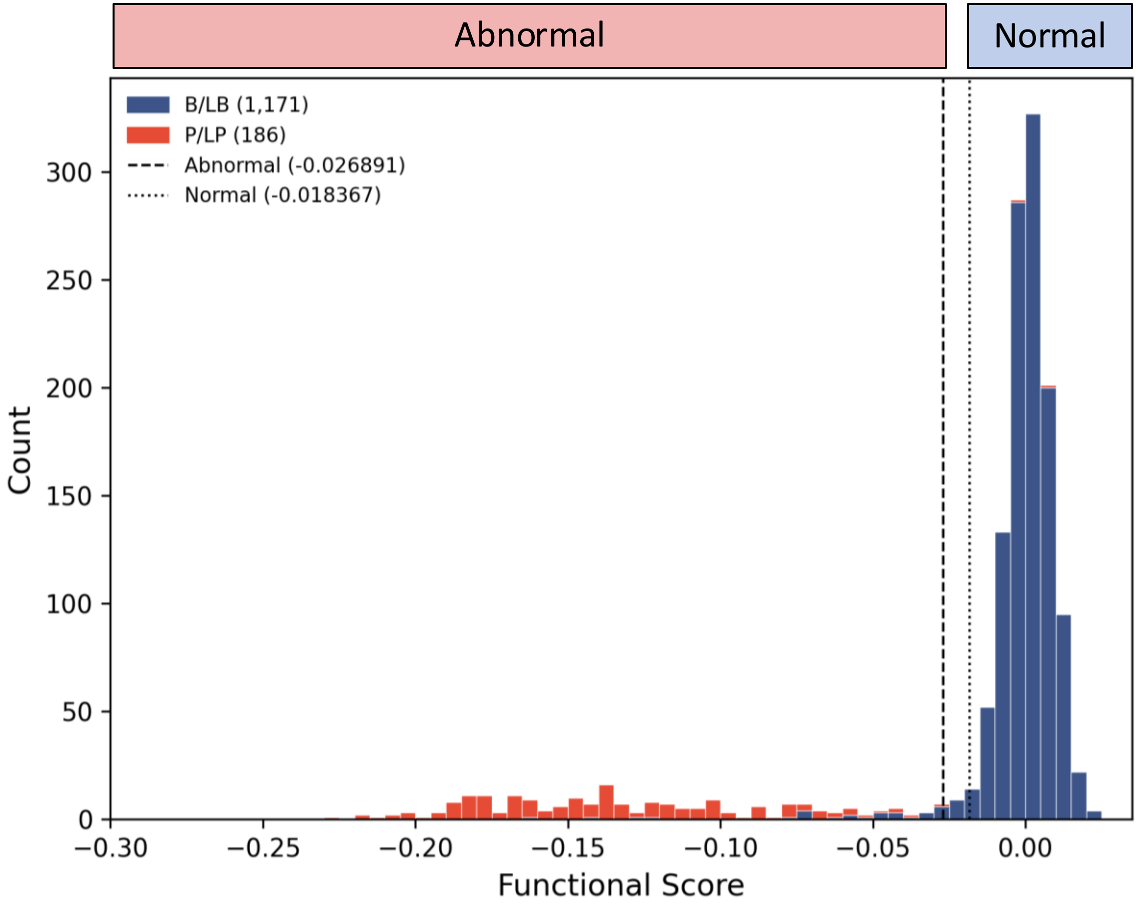

Assay = Saturation genome editing (SGE) of BAP1 in HAP1 cells

Assay result categories designated via Gaussian Mixture Model

Waters et Al. 2024 PMID: 38969833

### HOW PREDICTIVE ARE THE TEST CATEGORIES FOR CLINICAL OUTCOME?

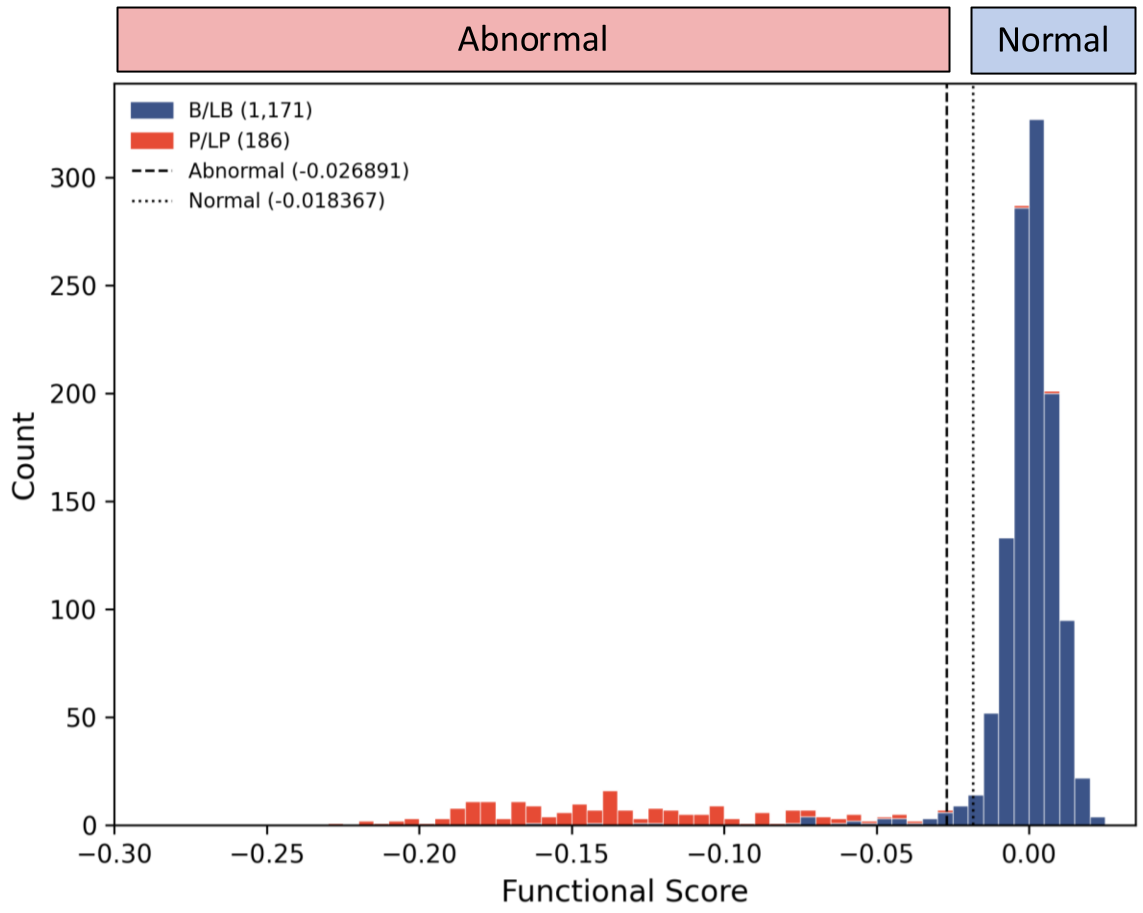

How does the assay perform according to the clinical reference?

Clinical outcome = pathogenicity

### CALCULATING ASSOCIATION OF TEST RESULT WITH CLINICAL OUTCOME? (LR)

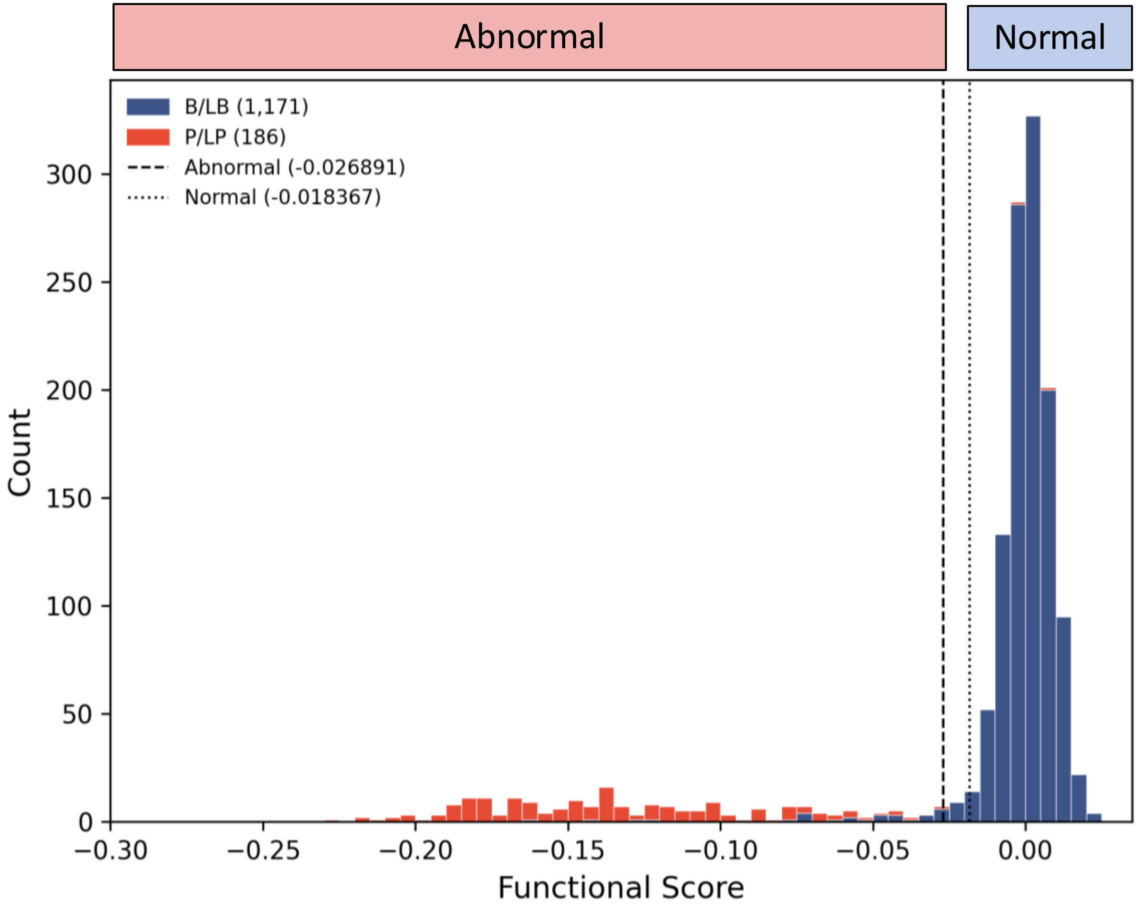

$$LR+ = \frac{P(\text{functionally abnormal}|\text{pathogenic})}{P(\text{functionally abnormal}|\text{benign})}$$

### CALCULATING ASSOCIATION OF TEST RESULT WITH CLINICAL OUTCOME? (LR)

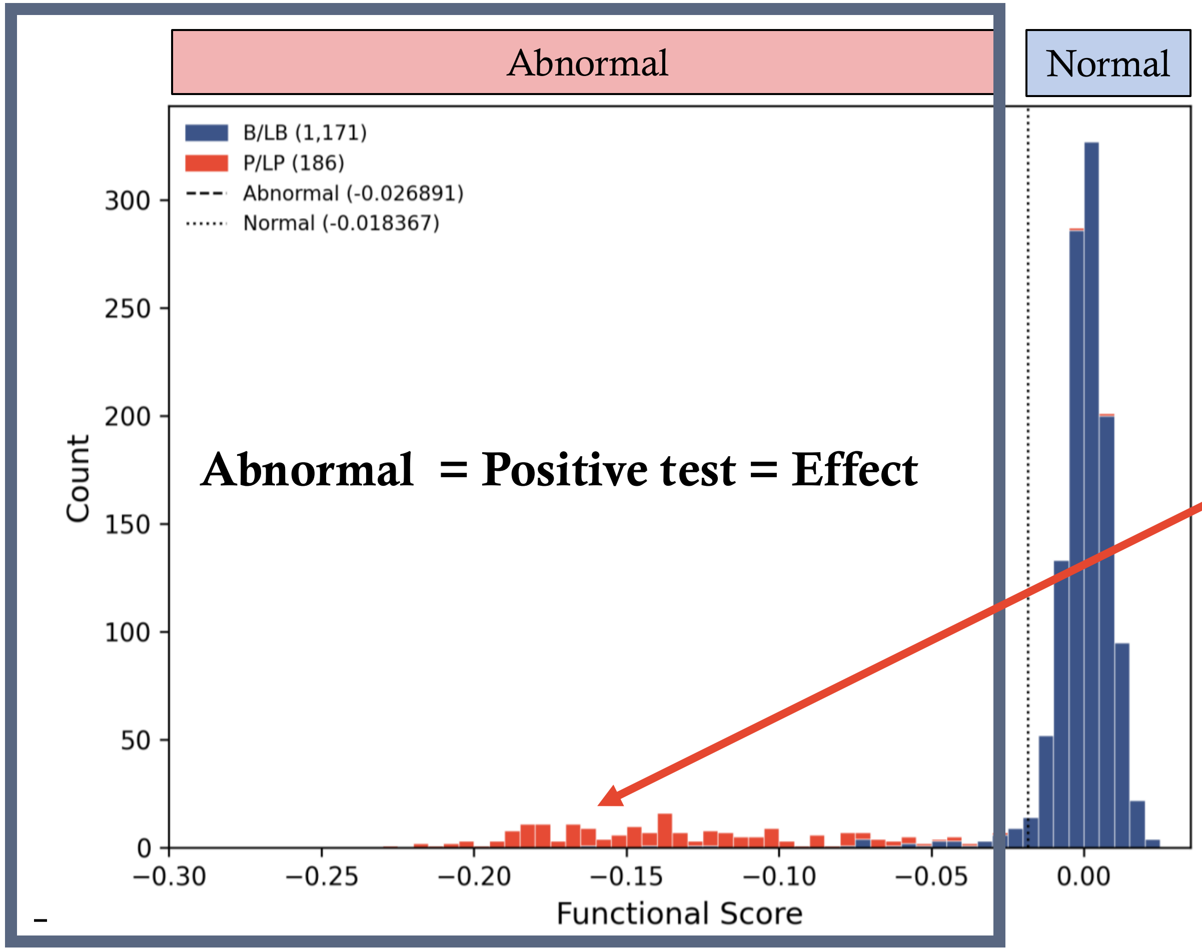

184/186 pathogenic variants in clinical reference set = abnormal

$$LR+ = \frac{P(\text{functionally abnormal}|\text{pathogenic})}{P(\text{functionally abnormal}|\text{benign})}$$

### CALCULATING ASSOCIATION OF TEST RESULT WITH CLINICAL OUTCOME? (LR)

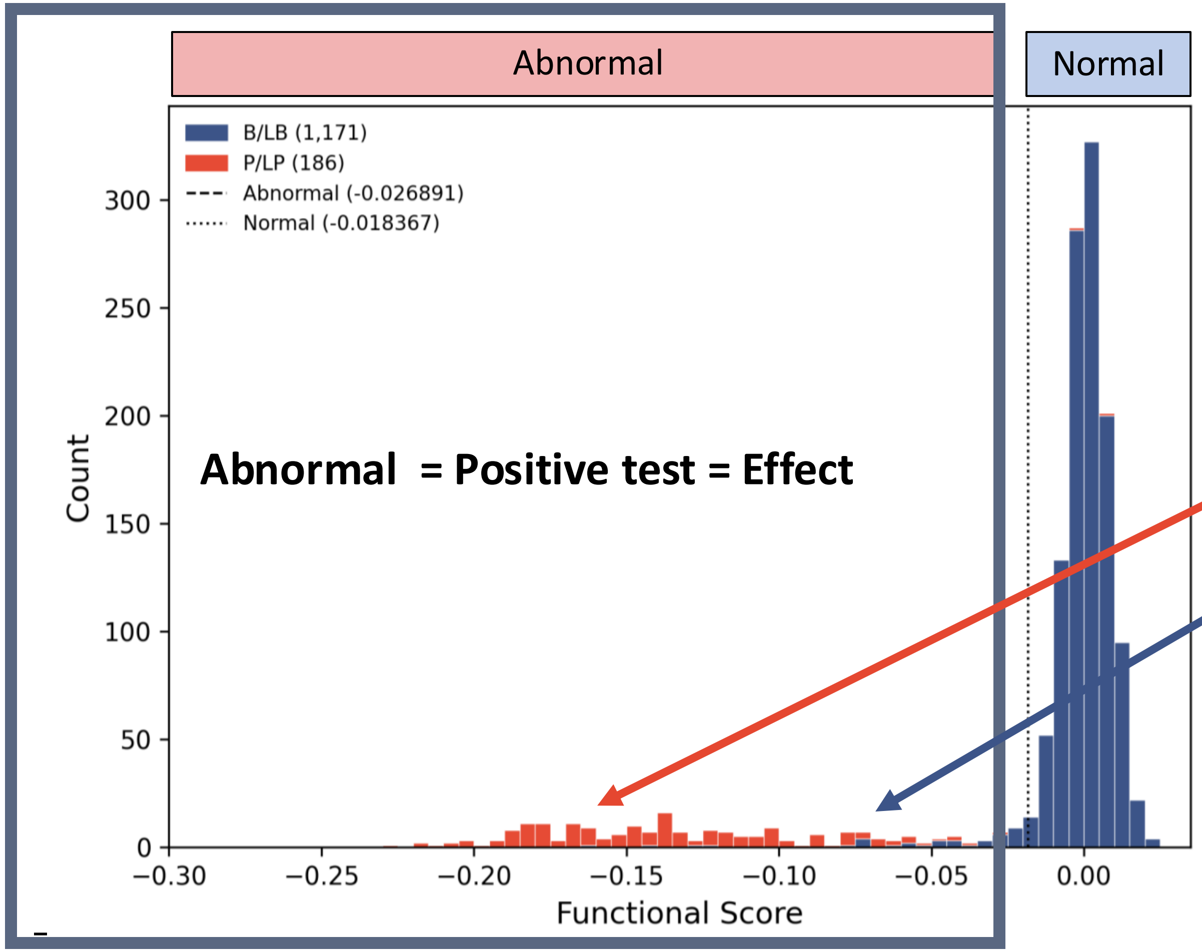

184/186 pathogenic variants in clinical reference set = abnormal

$$LR+ = \frac{P(\text{functionally abnormal}|\textbf{pathogenic})}{P(\text{functionally abnormal}|\textbf{benign})}$$

27/1171 benign variants in clinical reference set = abnormal

### CALCULATING LIKELIHOOD RATIO (LR)

#### LR positive

| Functional Category assigned | Benign reference set | Proportion benign | Pathogenic reference set | Proportion pathogenic | LR | Low CI | High CI |
|---|---|---|---|---|---|---|---|
| No-impact (normal) | 1132 | 0.97 | 2 | 0.01 | | | |
| intermediate | 12 | 0.01 | 0 | 0.00 | | | |
| Impact (abnormal) | 27 | 0.02 | 184 | 0.99 | | | |
| TOTAL= | 1171 | | 186 | | | | |

$$LR+ = \frac{P(\text{functionally abnormal}|\text{pathogenic})}{P(\text{functionally abnormal}|\text{benign})}$$

© 2026 American College of Medical Genetics and Genomics. All rights reserved

### CALCULATING LIKELIHOOD RATIO (LR)

LR positive

| Functional Category assigned | Benign reference set | Proportion benign | Pathogenic reference set | Proportion pathogenic | LR | Low CI | High CI |
|---|---|---|---|---|---|---|---|
| No-impact (normal) | 1132 | 0.97 | 2 | 0.01 | | | |
| intermediate | 12 | 0.01 | 0 | 0.00 | | | |
| Impact (abnormal) | 27 | 0.02 | 184 | 0.99 | 42.90 | 29.54 | 62.31 |
| TOTAL= | 1171 | | 186 | | | | |

0.9892/0.02306 = LR positive 42.90

**TRY the LR CALCULATOR**

https://zenodo.org/records/13324450

### WHAT ABOUT BENIGN ASSOCIATION? (LR-)

Essentially look at the inverse…

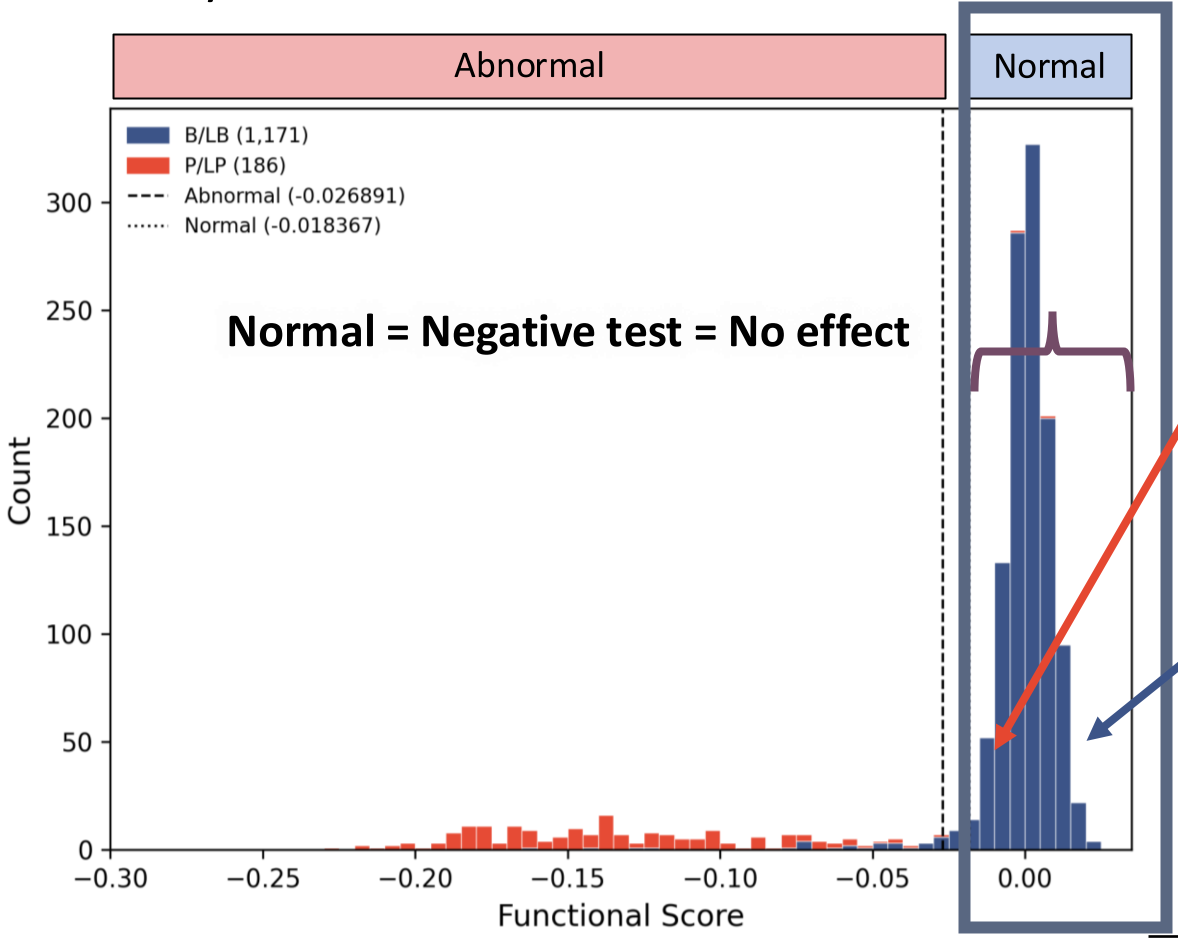

2/186 pathogenic variants in clinical reference set = normal

$$LR- = \frac{P(\text{functionally normal} \mid \text{pathogenic})}{P(\text{functionally normal} \mid \text{benign})}$$

1132/1171 benign variants in clinical reference set = normal

### CALCULATING LIKELIHOOD RATIO (LR)

LR negative

| Functional Category assigned | Benign reference set | Proportion benign | Pathogenic reference set | Proportion pathogenic | LR | Low CI | High CI |
|---|---|---|---|---|---|---|---|
| No-impact (normal) | 1132 | 0.97 | 2 | 0.01 | 0.01 | 0.00 | 0.04 |
| intermediate | 12 | 0.01 | 0 | 0.00 | - | - | - |
| Impact (abnormal) | 27 | 0.02 | 184 | 0.99 | 42.90 | 29.54 | 62.31 |
| TOTAL= | 1171 | | 186 | | | | |

0.010753/0.966695 = LR negative is 0.0111

### BUT WITH THE KURIS PHENOTYPE

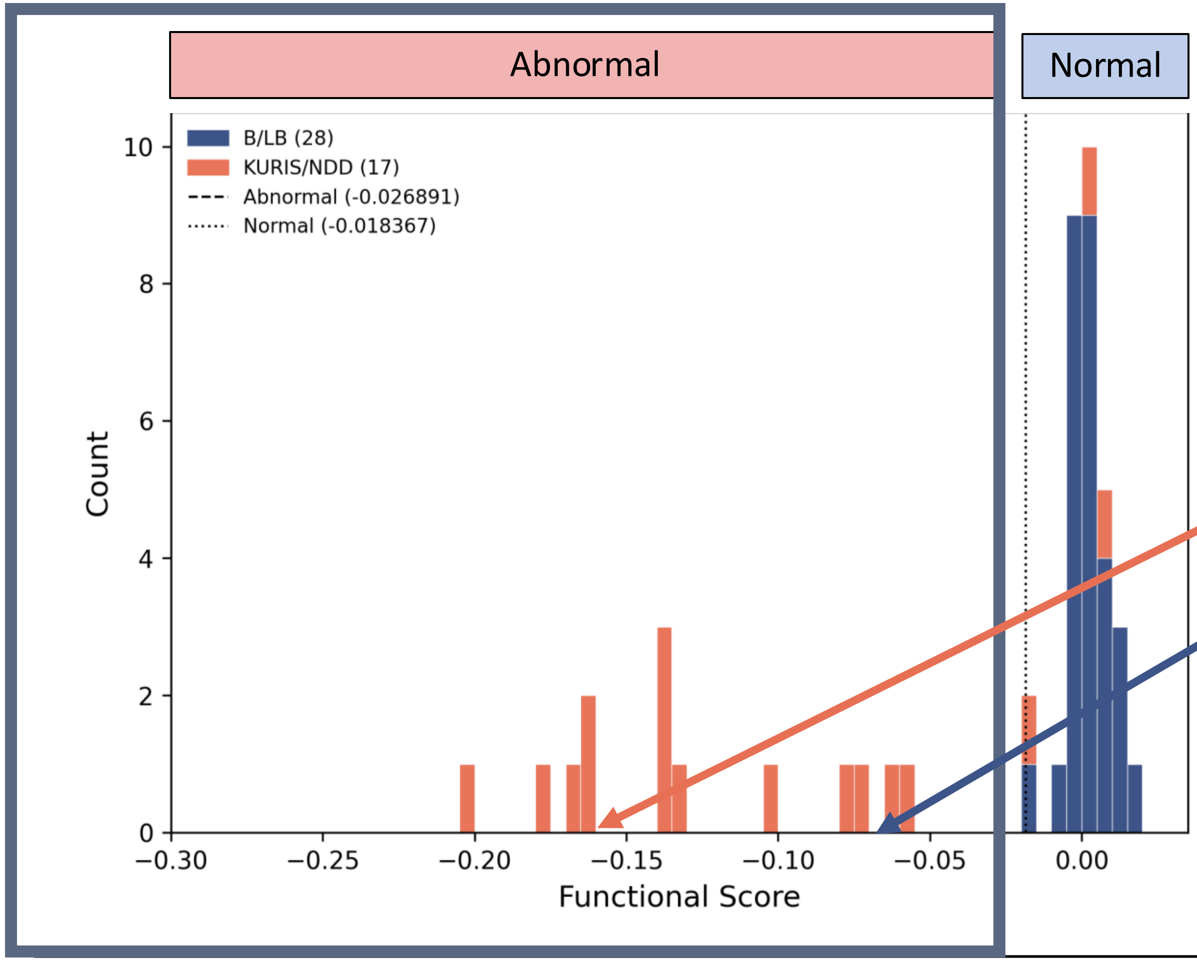

14/17 KURIS pathogenic variants in clinical reference set = abnormal

$$LR+ = \frac{P(\text{functionally abnormal}|\textbf{pathogenic})}{P(\text{functionally abnormal}|\textbf{benign})}$$

0/28 MISSENSE benign variants in clinical reference set = abnormal

### CALCULATING LIKELIHOOD RATIO (LR)

LR positive

| Functional Category assigned | Benign reference set | Proportion benign | Pathogenic reference set | Proportion pathogenic | LR | Low CI | High CI |
|---|---|---|---|---|---|---|---|
| No-impact (normal) | 28 | 1.00 | 3 | 0.18 | 0.18 | 0.06 | 0.49 |
| intermediate | 0 | 0.00 | 0 | 0.00 | - | - | - |
| Impact (abnormal) | 0 | 0.00 | 14 | 0.82 | 46.72 | 2.97 | 736.15 |
| TOTAL= | 28 | | 17 | | | | |

0.82/0.00* = LR positive 46.72

* Haldane-Anscombe method (+0.5) was used to resolve zeroes
(Haldane, JBS 1940, Anscombe, J 2956)

### BUT HOW DO I USE THE INFORMATION?

Determine which evidence strength the assay result supports

Tavtigian et Al 2018 aligned the ACMG/AMP variant classification model to a Bayesian framework

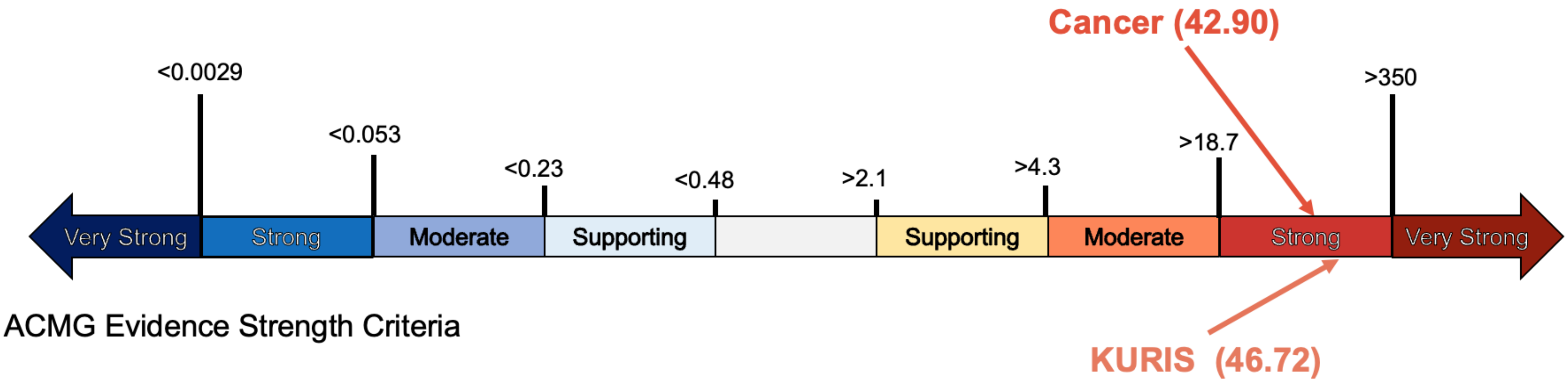

Tavtigian et Al. 2018 PMID: 29300386

### CONCLUSIONS

Likelihood ratio as a measure of evidence strength

- Reference set is everything (sets context for interpretation)
- Assay can give evidence of impact not mechanism. Both mechanisms lead to the same type of impact. Have to prove it!

### BACK TO THE CLINICAL CASE...

### BAP1: c.687C>A p.Asn229Lys

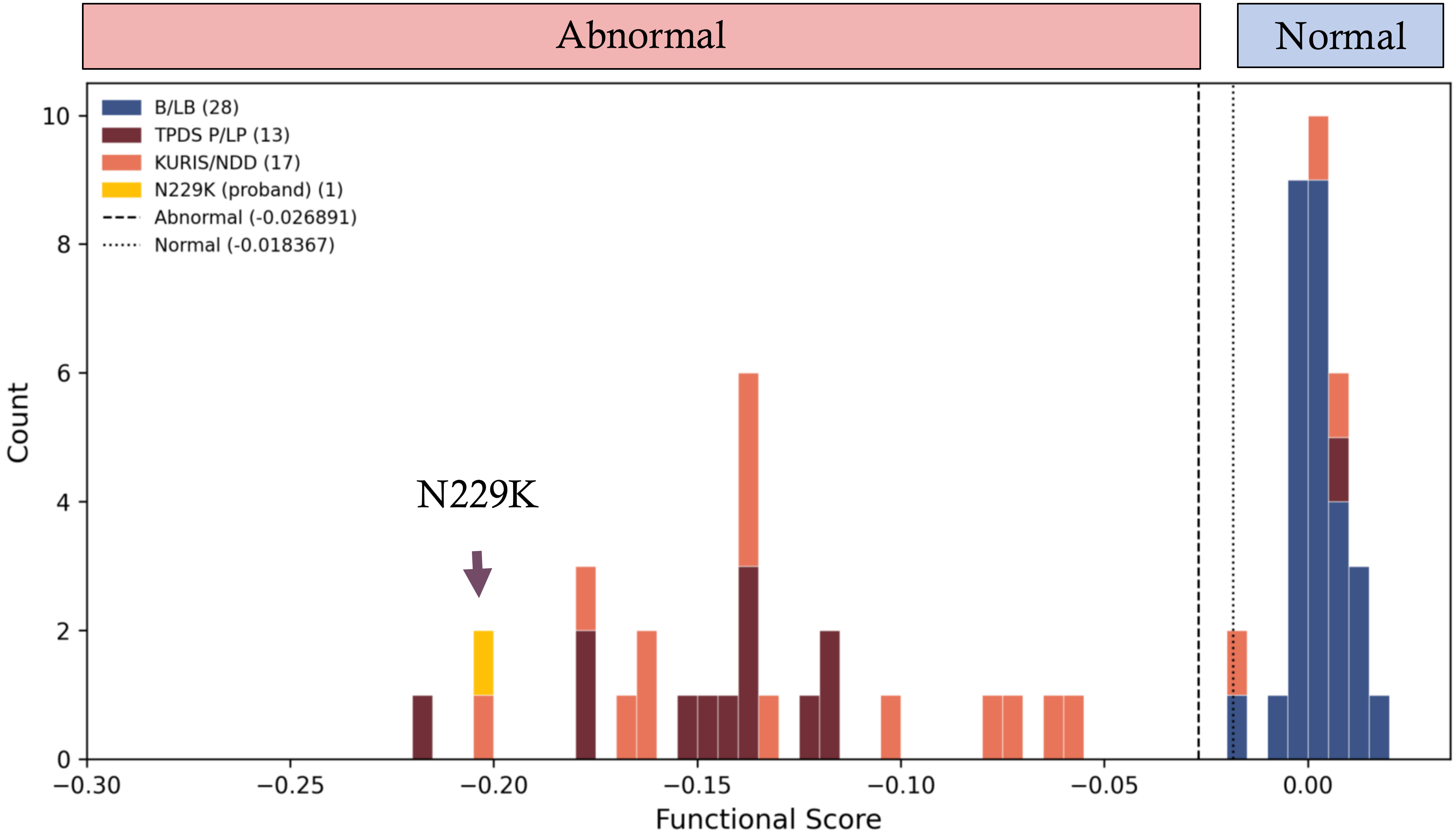

Functional score = -0.20

**Abnormal**

PS3_Strong

### BAP1:c.687C>A (p.Asn229Lys) CLASSIFICATION:
### LIKELY PATHOGENIC
#### The end of a long diagnostic odessey

| | |
|---|---|
| **PS2_Moderate** | Confirmed *de novo* in proband w/ consistent but non-specific phenotype |
| **PS4_Supporting** | Observed in >2 additional individuals with NDD |
| **PM2_Supporting** | Absent from large population databases |
| PS3_Strong | Functionally abnormal in a well validated assay |

### RESOURCES

- LR calculator - The simple excel likelihood ratio calculator developed by the ENIGMA VCEP
  - https://zenodo.org/records/13324450
  - Free LR calculators such as https://www.medcalc.org/en/calc/likelihoodratios.php or follow the method in Deeks et al. 2004.

- MaveDB - a public repository for datasets from Multiplexed Assays of Variant Effect (MAVEs)

- MaveMD - https://www.mavedb.org/mavemd

- ClinVar - https://www.ncbi.nlm.nih.gov/clinvar/

- AVE channel - https://www.varianteffect.org/
